# A rapid review of the effectiveness of interventions to enhance equitable or overall access to mental health services by ethnic minority groups

**DOI:** 10.1101/2024.05.16.24307468

**Authors:** Deborah Edwards, Judit Csontos, Elizabeth Gillen, Sioned Gwyn, Juliet Hounsome, Meg Kiseleva, Mala Mann, Abubakar Sha’aban, Rhiannon Tudor Edwards, Jacob Davies, Ruth Lewis, Alison Cooper, Adrian Edwards

## Abstract

It is estimated that one in four people will experience poor mental health throughout their lifetime. However, ethnic minority groups, refugees and asylum seekers experience more barriers accessing mental health services and have poorer mental health outcomes than those from non-ethnic minority groups. Evidence suggests that interventions that improve access and engagement with mental health services may help reduce disparities affecting ethnic minority groups. This review aims to assess the effectiveness of interventions that enhance equitable or overall access to mental health services by ethnic minority groups.

The review included evidence available up until 19^th^ December 2023.

Psycho-educational interventions that focused on providing culturally appropriate information, showed mixed results for help seeking behaviour, improvements in depressions stigma. Multi-component interventions within healthcare settings had mixed results. Some studies showed positive outcomes; such as increased help seeking intentions and improved attendance rates, while others did not show significant differences in outcomes.

Interventions that included integrating specialist mental health services within primary care resulted in variable outcomes. The findings of interventions incorporating language support into mental health services were also variable. The effectiveness of interventions to enhance the cultural competency of mental health services varied across studies.

---

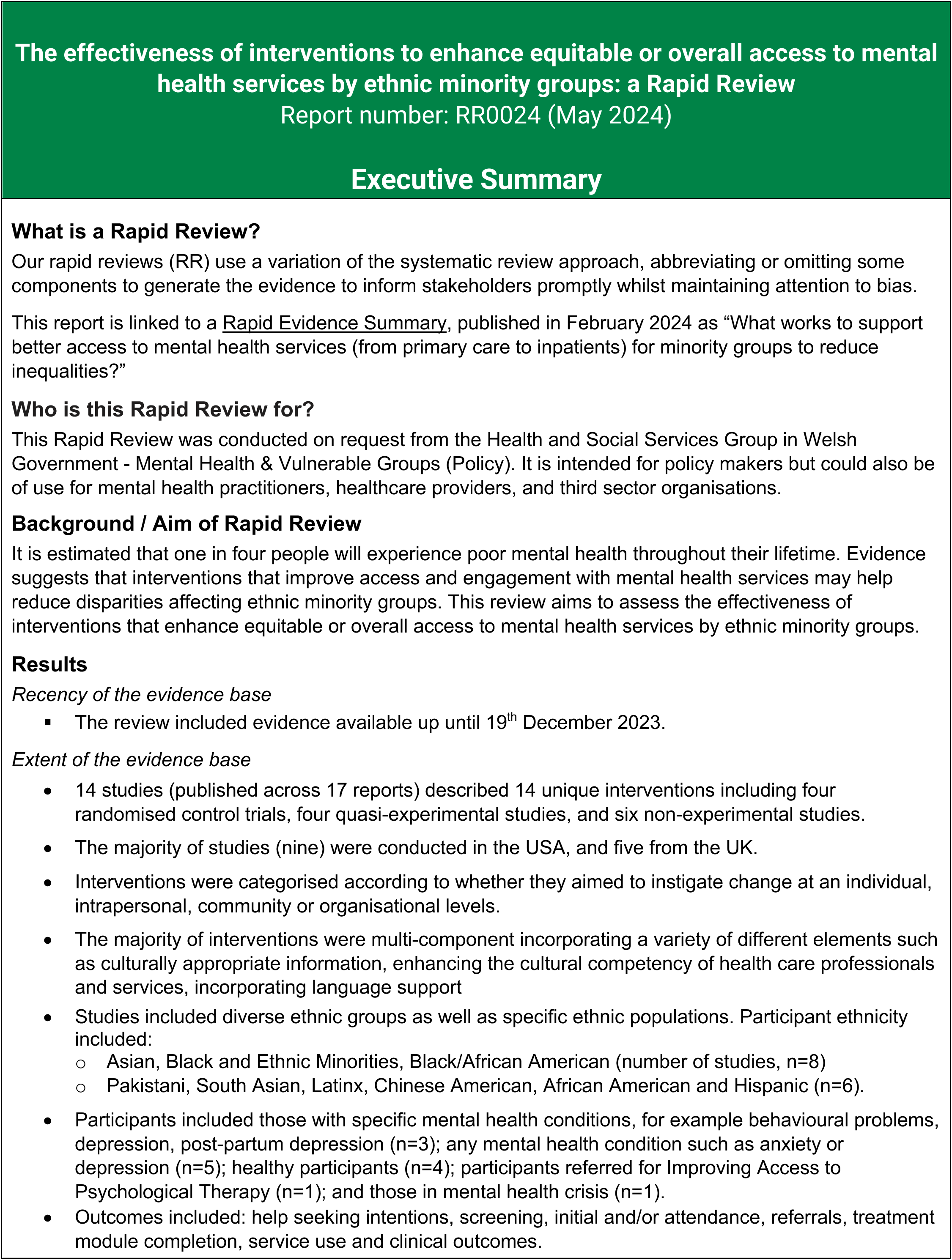
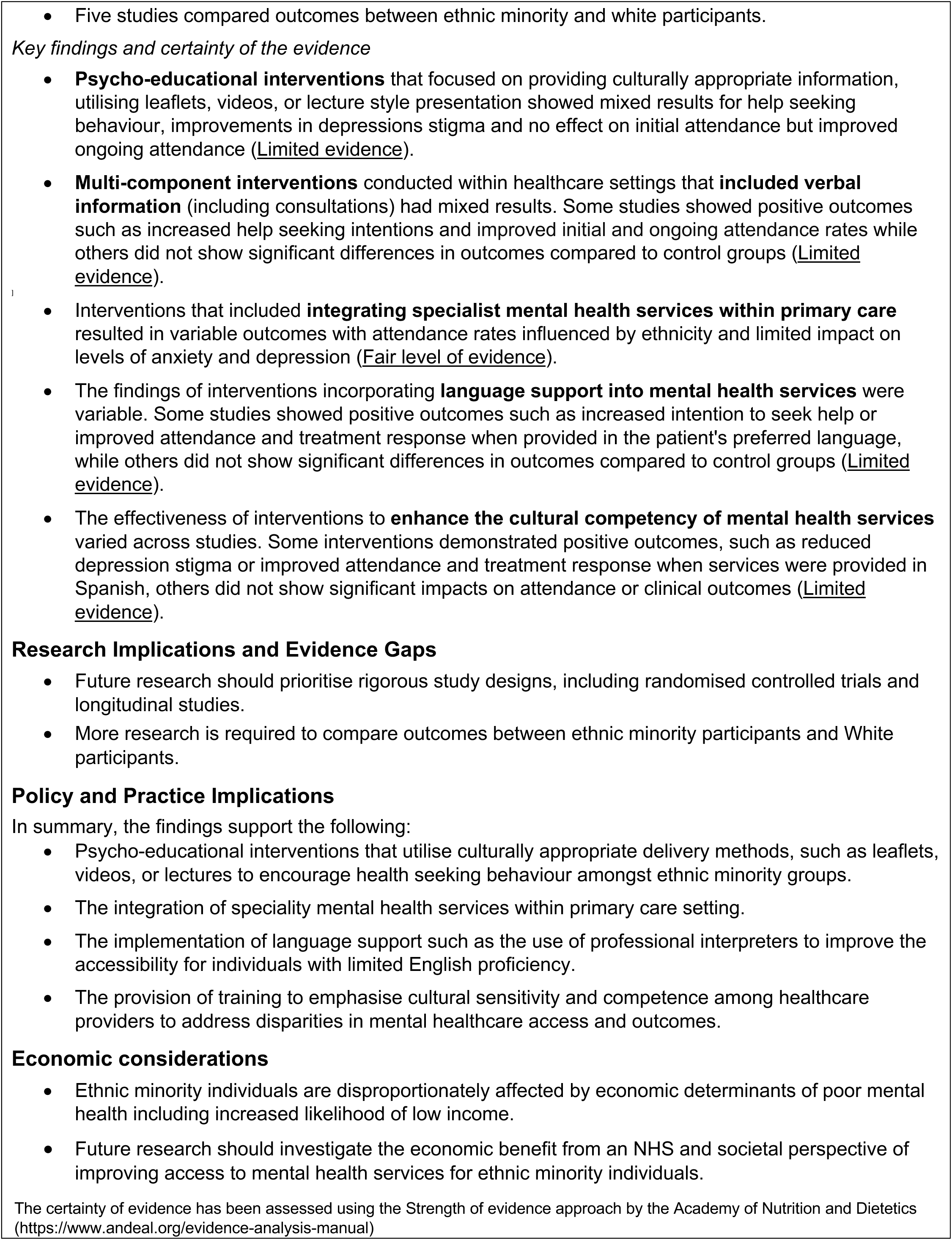

## 1. BACKGROUND

### 1.1 Who is this review for?

This rapid review was conducted as part of the Health and Care Research Wales Evidence Centre Work Programme. The above question was suggested by Health and Social Services Group Welsh Government - Mental Health & Vulnerable Groups (Policy). This rapid review is intended for policy makers but could also be of use for mental health practitioners, healthcare providers, and third sector organisations.

### 1.2 Background and purpose of this review

It is estimated that one in four people will experience poor mental health^1^ throughout their lifetime (Centre for Mental Health 2020). Evidence suggests that ethnic minority groups^1^ experience inequalities in access, experience and outcomes of mental health care (Lowther-Payne et al. 2023, Bansal et al. 2022). At the same time, these groups are at higher risk of mental health conditions^1^ and this risk is often associated with being disproportionally impacted by detrimental social factors, such as racism and poverty (Bignall et al. 2019).

Wales is home for diverse ethnic minority groups, including approximately 89,000 people who identify as Asian, Asian British or Asian Welsh, 28,000 Black, Black British, Black Welsh, Caribbean or African individuals, 49,000 mixed or multiple ethnic people, 26,000 members of other ethnic groups (ONS 2022), and 3,630 Gypsy and Irish Traveller residents (ONS 2023). Together for Mental Health, the Welsh Government’s mental health strategy has highlighted the need to consider equality since 2012 (Welsh Government 2012). However, the equality impact assessment of this strategy found that since the publication of Together for Mental Health, stigma and discrimination were still more prevalent for people with protected characteristics, including ethnic minority groups (Welsh Government 2014). In addition, the COVID-19 pandemic highlighted systemic issues, and disproportionately affected ethnic minority groups, prompting the Welsh Government to focus more on reducing health inequalities in their Together for Mental Health updated Delivery Plan (Welsh Government 2020). A recently published research review of the Together for Mental Health Delivery Plan acknowledged that ethnic minority groups in Wales had poorer mental health outcomes than the wider public while identifying potential barriers to access and service provision (Lock et al. 2023). Improvement in the cultural competency of mental health services and providers in Wales was identified as necessary to help the engagement of ethnic minority groups (Lock et al. 2023). Additionally, issues with multi-lingual service provision in common international languages was also mentioned in the report, which could negatively impact on help seeking for members of ethnic minority groups who can discuss their condition better in a language other than English (Lock et al. 2023). Welsh Government is currently consulting on a new Mental Health and Wellbeing Strategy for Wales. The draft strategy sets out ten overarching principles – one of which is equity of access, experience and outcomes without discrimination: ensuring services and support are accessible and appropriate for all. This means understanding the barriers people face and putting necessary systems in place so that when people get support, there is equity in terms of experiences and outcomes. To achieve this, support and services will need to be culturally and age appropriate and meet the needs of Welsh speakers, ethnic minority people, LGBTQ+ communities and people with sensory loss.

Services will also need to meet the needs of under-served groups such as people with co-occurring substance misuse, people who are care experienced, neurodivergent people and people who are experiencing poverty and people who are experiencing homelessness.

Evidence from the wider international literature also suggests that disparities^1^ in ethnic minority groups’ engagement with mental health services exist along the entire care pathway, with them being less likely to initiate mental health care and more likely to end treatment early (Aggarwal et al. 2016, Interian et al. 2013). These disparities are linked to various individual, organisational, and systemic barriers (Aggarwal et al. 2016). Individual-level barriers may include insufficient information to make treatment decisions, communication difficulties and linguistic issues, lack of trust in service providers, psychological distress, fear of stigma, and cultural beliefs resulting in feeling shame about seeking mental health support. Organisational-level barriers refer to unequal access to services and lack of cultural competence in service providers. The systemic level includes poor funding of mental health services and inaccessibility of information about available services as well as broader issues such as lack of access to transportation or childcare necessary to attend mental health services (Aggarwal et al. 2016).

To improve ethnic minority groups’ access to mental health services in Wales, a Mental Health Ethnic Minorities Task and Finish Group – jointly chaired by Welsh Government and the Wales Alliance for Mental Health was set up as part of the Anti-Racist Wales Action Plan (Welsh Government 2022). Moreover, suggestions were made that the new mental health strategy should be developed with the involvement of community organisations, the third sector and the NHS to make sure that the needs and experiences of ethnic minority groups are considered (Welsh Government 2022). To support these plans, it is also crucial to know what evidence is available on interventions that could support equitable^2^ or overall access to mental health services by ethnic minority groups. It is thought that effective interventions to improve access and engagement with services may reduce disparities (Interian et al. 2013). Preliminary work for this review looking at the wider evidence base has been published as a separate <u>Rapid Evidence Summary</u>.

## 2. RESULTS

This section details the extent of the evidence base and findings of the included research reporting on effectiveness of interventions to enhance equitable or overall access to mental health services by ethnic minority groups.

The overview of the evidence base (section 2.1) provides a description of the characteristics of the available evidence, including participants (age, gender, ethnic minority groups, mental health conditions), study design, country of origin of the research, interventions, settings and outcome/s of interest. A summary of the findings of the included studies is presented in section 2.2, which is structured around the type of intervention, type of outcomes measures (help seeking intentions, screening, initial attendance, ongoing attendance, referrals, service use and clinical outcomes), and target population characteristics.

A summary of the interventions included in the review is provided in Table 1, and a summary of the findings and the effectiveness of each intervention is provided in Table 2. Table 3 provides a summary of the strength of the evidence and assigns a Grade to indicate whether this was Good, Fair or Limited.

**Table 1:** Summary of interventions evaluated by included studies and type of comparison made.

| Authors/ year (country)<br>Study design<br>Name and type of intervention/control | Mode of delivery and duration/intensity | Data analysis |
| --- | --- | --- |
| **Interventions focusing on change at the individual level** |  |  |
| **Alvidrez et al. 2005 (USA)**<br>**Study design**<br>Cross-sectional post-test only quasi-experimental<br>**Intervention**<br>Psycho-educational to promote psychotherapy entry and attendance for older African Americans<br>**Comparison**<br>Historical comparison group<br>Referred for psychotherapy in the 12 months prior to the intervention | **Mode of delivery**<br>Delivered by researchers<br>Face to face psycho-education sessions<br>**Duration**<br>Intervention: 15-minute scripted psycho-education session before the day of their first scheduled psychotherapy appointment covering six topics<br>Study duration: 14 months | **Data analysis**<br>Treatment entry and attendance, the impact of psycho educator ethnicity (African American or other) on treatment entry and attendance were compared between the pre-implementation and post-implementation period |
| **Martin et al. 2022 (USA)**<br>**Study design**<br>RCT<br>**Intervention**<br>Psycho-educational intervention to increase help seeking intentions around addressing difficulties coping with depressive symptoms for adolescents from ethnic minority groups<br>**Control**<br>Control video | **Mode of delivery**<br>Delivered by researchers<br>Face to face - short video interviews with a 16-year-old Black female actor<br>Video 1 ("Depressed"): The videos address difficulties coping with depressive symptoms, thoughts that life is not worth living, false assumptions about treatment, and how and when she decided to seek help<br>Video 2 ("Depressed adjusted"): is adjusted to aspects unique to being a Black adolescent girl (e.g., experienced or internalised racism)<br>Video 3 (control): describes hobbies and lacks the description of depressive symptoms or any other mental health-related content<br>**Duration**<br>Intervention: 117 seconds long and were administered once<br>Study duration: not reported | **Data analysis**<br>Depression stigma and help seeking intention were compared between baseline and post intervention, and across the groups receiving different psycho-educational videos (1, 2 and control)<br>Depression stigma for Black participants were also analysed relative to non-Black participants |
| **Mukadam et al. 2018 (UK)**<br>**Study design**<br>Cluster RCT<br>**Intervention**<br>Patient education DVD and bilingual leaflet about dementia and getting help for memory symptoms for South Asian adults without known dementia aged >50 years<br>**Control**<br>No DVD/leaflet information sent | **Mode of delivery**<br>Delivered by researchers<br>Leaflet and DVD sent out in the post with a letter on headed paper from the practice<br>**Duration**<br>Intervention: not reported<br>Study duration: 1 year | **Data analysis**<br>Help seeking intention (based on the APEND questionnaire) was compared between intervention (patient education DVD and leaflet) and control (no DVD and leaflet) |
| **Teng and Friedman 2009 (USA)**<br>**Study design**<br>Longitudinal pre-post non-experimental<br>**Intervention**<br>Psycho-educational intervention to increase awareness of mental health issues and available resources among elderly Chinese Americans | **Mode of delivery**<br>Delivered by the co-author of the research report and no further details<br>Provided through a faith based organisation<br>Face to face - didactic presentation delivered in English/Mandarin<br>**Duration**<br>Intervention: 1 hour<br>Study duration: not reported | **Data analysis**<br>Help seeking intentions for psychiatric symptoms were compared immediately before and after the intervention |
| **Interventions focusing on change at the organisational level** |  |  |
| **Arean et al. 2008 (USA)**<br>**Study design**<br>RCT<br>**Intervention**<br>Integrating mental health and substance abuse services into primary care settings for older adults (> 65 years) from ethnic minority groups<br>**Control**<br>Referral to community services | **Mode of delivery**<br>Delivered by practitioners within the primary care setting<br>Face to face evaluation and subsequent referral to primary care clinic or community services<br>**Duration**<br>Intervention: not reported<br>Study duration: not reported | **Data analysis**<br>Percentage of having at least one mental health visit, average number of visits, estimated number of days to first visit, anxiety and depression scores were compared between intervention (integrated care) and control (speciality mental health care) for different ethnicities<br>Outcomes for different ethnicities were also analysed relative to white participants |
| **Evans et al 2014 (UK)**<br>**Study design**<br>Cross-sectional post-test only quasi-experimental | **Mode of delivery:**<br>Delivered by practitioners within the primary care setting | **Data analysis**<br>Referral rates were descriptively compared between GP services with and without a link worker |
| **Intervention**<br>Placing mental health link workers in GP surgeries to increase Black and Minority Ethnic referrals to the IAPT programme<br>**Control**<br>GP surgeries without link workers | Face to face explanation of psychological therapy and raising awareness of common mental disorders in Black and Minority Ethnic groups<br>**Duration:**<br>Intervention: not reported<br>Study duration=Approx. 17 months |  |
| **Gorman et al 2021 (USA)**<br>**Study design**<br>Cross-sectional post-test only quasi-experimental<br>**Intervention**<br>Routine depression screening by medical assistants across ethnic minority groups<br>**Comparison:**<br>Routine depression screening by physicians in the 12 months prior to the intervention (historical comparison group) | **Mode of delivery**<br>Delivered by practitioners within the primary care setting<br>Face to face - medical assistants verbally administered the PHQ-2 and recorded the patients' responses in the electronic health record<br>**Duration/Intensity:**<br>Intervention: not reported<br>Study duration: 2 years | **Data analysis**<br>Depression screening rates were compared between routine screening by medical assistant and routine screening by physician<br>Outcomes for different ethnicities were also analysed relative to white participants |
| **Interventions focusing on change at both interpersonal and organisational levels** |  |  |
| **Ortega and Rosenheck 2002 (USA)**<br>**Study design**<br>Longitudinal interrupted time series non-experimental<br>**Intervention:**<br>Client-clinician ethnic and language matching for White and Hispanic homeless clients with severe mental illness in ongoing community treatment | **Mode of delivery:**<br>Delivered by community practitioners working in mental health<br>Face to face assertive community treatment<br>**Duration**<br>Intervention: not reported<br>Study duration: 3 years | **Data analysis:**<br>Analysis of covariance of 12-month change to determine statistically significant interactions of case managers' race or ethnicity and clients' race or ethnicity with regard to entry for into treatment<br>Outcomes for different ethnicities were also analysed relative to white participants |
| **Yeung et al. 2004 (USA)**<br>**Study design:**<br>Repeat cross-sectional pre-post non-experimental<br>**Intervention**<br>Psychiatry and primary healthcare service integration for Chinese American adults<br>**Comparison** | **Mode of delivery**<br>Delivered by practitioners within the primary care setting<br>Face to face training for PCPs<br>A primary care nurse acted as the "bridge" or care manager.<br>A liaison psychiatrist provided services within the primary care clinic<br>**Duration** | **Data analysis**<br>Referral and attendance rates were compared between the pre-implementation and post-implementation period |
| Historical service data | Intervention: Training of PCPs on established treatment guidelines (two, 1-hour seminars)<br>Training of PCPs in cultural sensitivity (1-hour seminar)<br>Appointments with liaison psychiatrist scheduled within two weeks<br>Study duration: Records assessed for the period of the project (Jan–Dec 2000) and the year prior to it (Jan–Dec 1999) |  |
| **Interventions focusing on change at both community and organisational levels** |  |  |
| **Hackett et al. 2009 (UK)**<br>**Study design**<br>Repeat cross-sectional pre-post non-experimental<br>**Intervention**<br>Enhancing Pathways Into Care project: information provision, community engagement; link worker and more appropriate and responsive services for those from the Pakistani community in mental health crisis<br>**Comparison**<br>Historical service data | **Mode of delivery**<br>Delivered by community practitioners working in mental health<br>Face to face - Crisis Resolution Home Treatment joined with a community organisation called the Pakistani Muslim Centre to work in partnership<br>**Duration**<br>Intervention: not reported<br>Study duration: 2002-2006 clinical database review / 2005-2006 case note review | **Data analysis**<br>Referrals (to crisis resolution home or inpatient ward) and service use (admission to hospital or home treatment, length of stay) were descriptively compared between the pre-implementation and post-implementation period |
| **Robidoux et al. 2023 (USA)**<br>**Study design**<br>Repeat cross-sectional pre-post non-experimental<br>**Intervention**<br>Postpartum depression screening and referral to behaviour services (by community health workers) for Latinx immigrant mothers<br>**Comparison**<br>Historical service data | **Mode of delivery**<br>Delivered by community practitioners working in mental health<br>Face to face<br>**Duration**<br>Intervention: not reported<br>Study duration: 1 year | **Data analysis**<br>Screening and referral rates were compared between the pre-implementation and post-implementation period |
| **Interventions focusing on change at both individual and organisational levels** |  |  |
| **Seabrooke and Milne 2009 (UK)**<br>**Study design**<br>Repeat cross-sectional pre-post non-experimental | **Mode of delivery**<br>Delivered by practitioners within the primary care setting<br>Letters and information leaflets mailed to patients<br>Patient information leaflets were left in waiting area. | **Data analysis**<br>Initial attendance at GP and referral rates to specialist services were descriptively compared between the pre-implementation and post-implementation period |
| **Intervention**<br>Patient education and health care professional training to facilitate access to dementia screening and diagnosis in for older Asians<br>**Comparison**<br>Historical service data | Specialist dementia training for practice nurse<br>**Duration**<br>Intervention: not reported<br>Study duration: Post implementation - Sept 2006 – Oct 2007<br>Post implementation - 3 months prior to implementation of the project in 05/06 |  |
| **Interventions focusing on change across individual, interpersonal and organisation levels** |  |  |
| **Michelson and Day 2014 (UK)**<br>**Study design**<br>Cross-sectional post-test only quasi-experimental<br>**Intervention**<br>Pre-intake intervention designed to enhance initial engagement at CAMHS targeted at primary parental caregivers of all children and adolescents from Black and Minority Ethnic groups referred and accepted into CAMHS<br>**Comparison**<br>Historical comparison group<br>Families who received standard clinic procedure (opt-out letter six months prior to the intervention) | **Mode of delivery**<br>Delivered by community practitioners working in mental health<br>The intervention was a telephone conversation during which a key worker attempted to systematically identify and address possible barriers to initial attendance at CAMHS<br>Practitioners working in schools could deliver the intervention face-to-face<br>**Duration/Intensity**<br>Intervention: 20–30 min telephone conversations<br>Study duration: 6 months. | **Data analysis**<br>Initial and ongoing attendance (at least one appointment of the first three scheduled appointments) outcomes (attended, did not attend, discharged, cancelled) were compared between the intervention and historical comparison group<br>DNA rates were compared between Black and Minority Ethnic and White participants |
| **Sanchez et al. 2022 (USA)**<br>**Study design**<br>RCT<br>**Intervention**<br>Person-centred cultural assessment (with primary caregivers of children aged 2-7 from ethnic minority groups with behaviour problems)<br>**Control**<br>Assessment as usual | **Mode of delivery**<br>Delivered by community practitioners working in mental health<br>Face to face - Clinical Assessment and Cultural Formulation Interview<br>**Duration**<br>Intervention: 2-3, one hour sessions<br>Study duration: 3 years | **Data analysis**<br>Initial and ongoing attendance, and clinical outcomes (treatment response, first treatment module completion) were compared between cultural assessment and assessment as usual group<br>The impact of language (Spanish or English) on outcomes was also analysed |

Key: IAPT: Improving Access to Psychological Therapy; MDT: multidisciplinary team; MH: mental health; PCPs: primary care practitioners; PHQ-2: the Patient Health Questionnaire Version 2; RCT randomised controlled trial

**Table 2:**
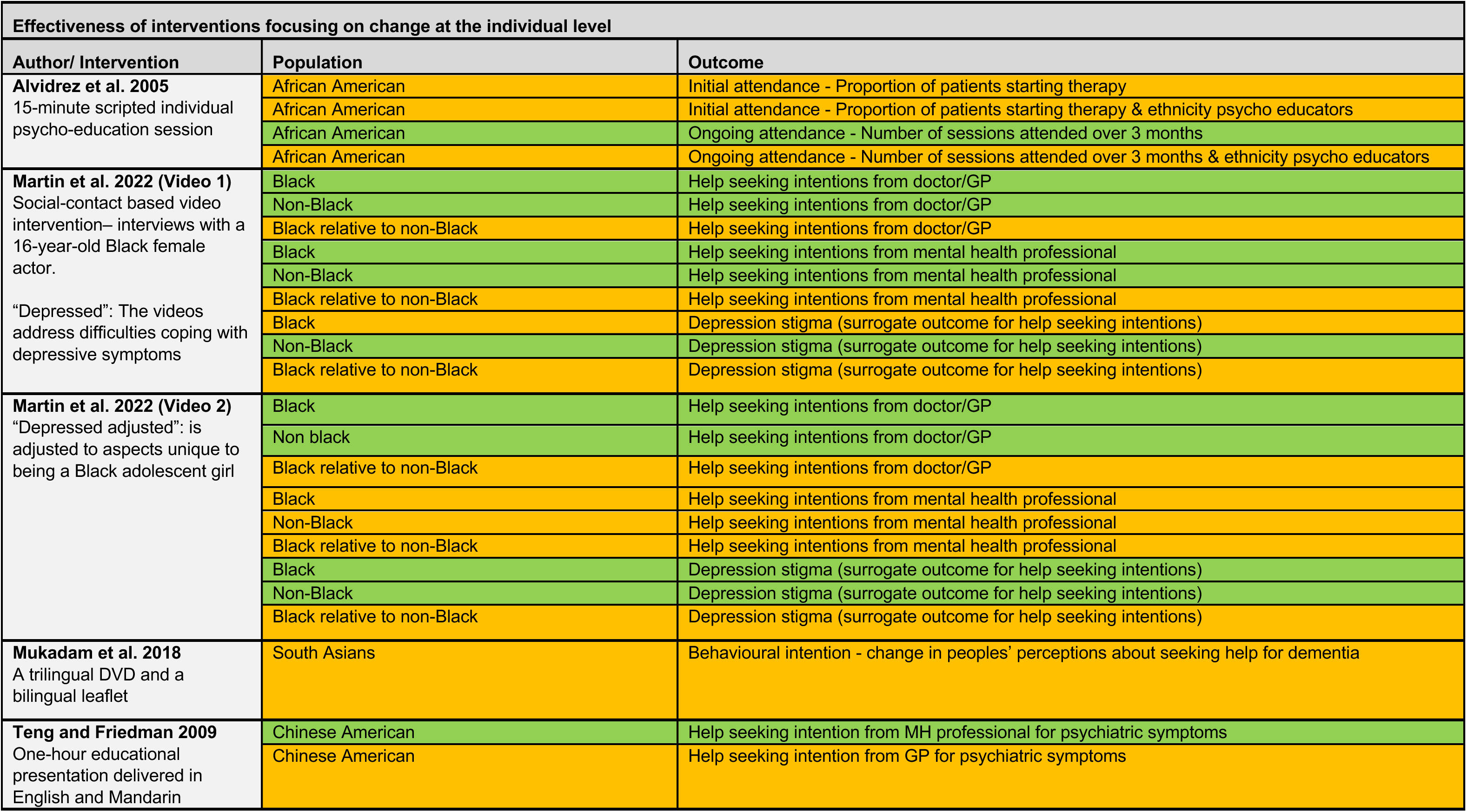
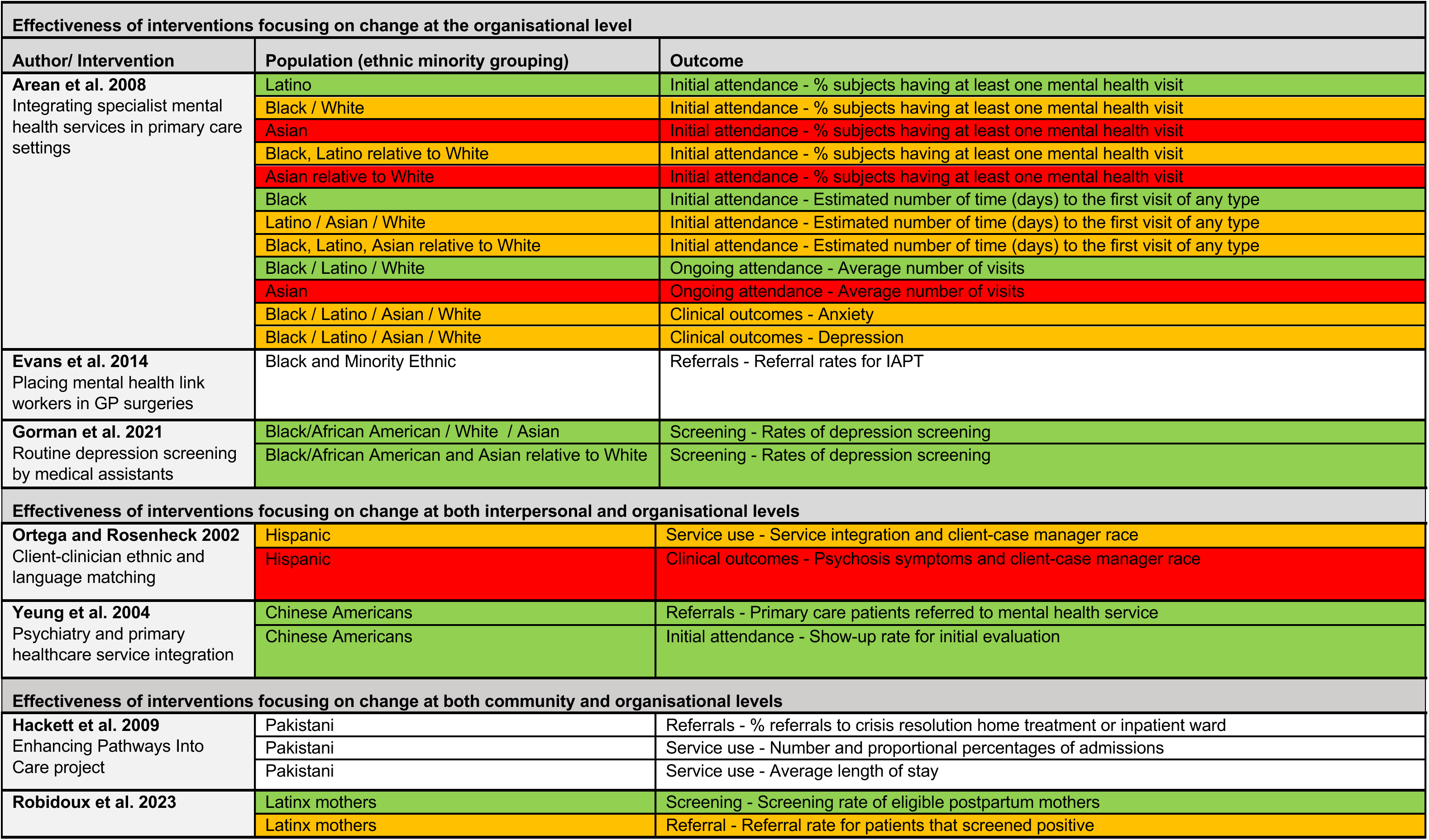
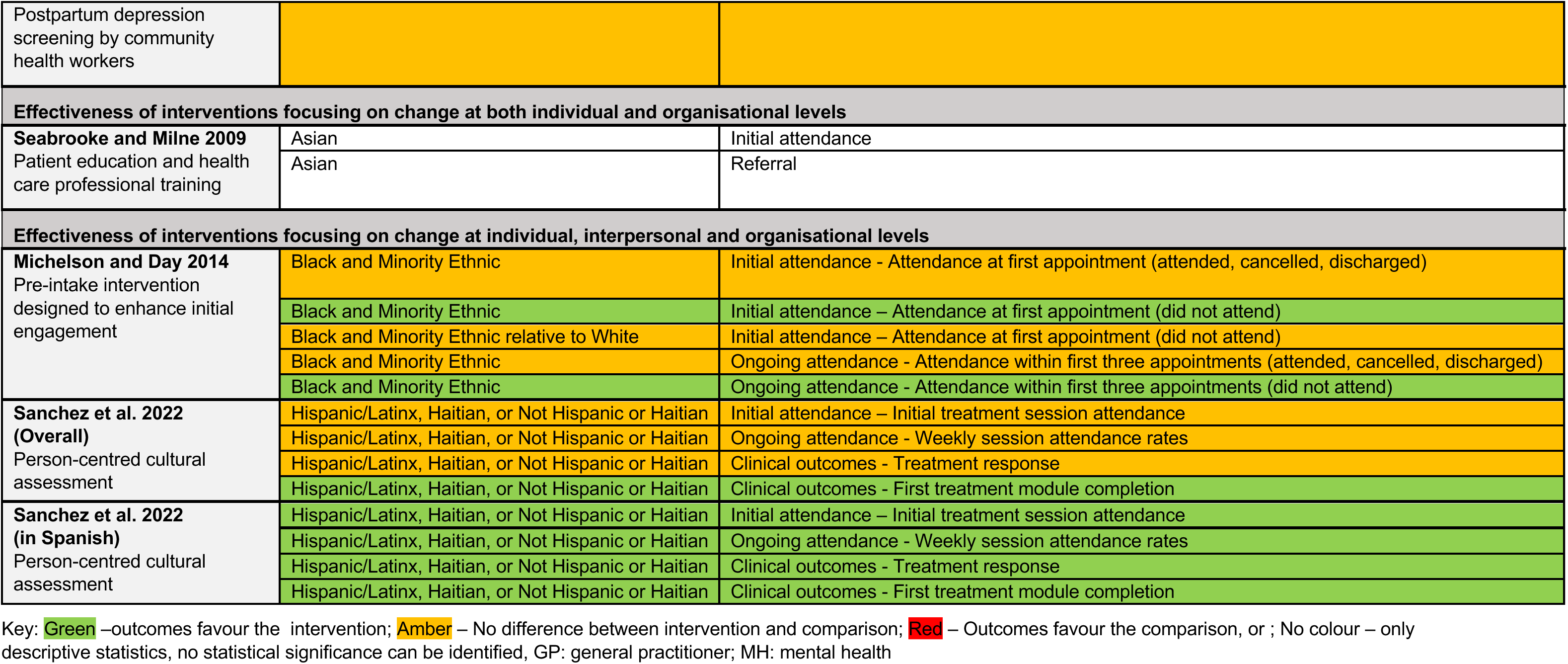
Summary of effectiveness by interventions, ethnic minority groups, and outcomes.

**Table 3:** Summary of strength of the evidence. The strength of the evidence supporting (overall findings based on each outcome) was established and graded based on the quality, quantity and consistency of the available evidence, the likely clinical impact and generalisability of the findings.

| Outcome | Quality | Consistency | Quantity | Clinical Impact | Generalisability | GRADE<sup>b</sup> |
| --- | --- | --- | --- | --- | --- | --- |
| **Interventions focusing on change at the individual level** |  |  |  |  |  |  |
| Ongoing appointment attendance<br>Alvidrez et al. 2005 | One study<br>medium quality | N/A | Limitations<br>(one study small sample size) | Limitations<br>(one study no effect size) | USA<br>Older adults<br>African American participants | Grade III |
| Ethnicity of psycho educator on initial or ongoing appointment attendance<br>Alvidrez et al. 2005 | One study<br>medium quality | N/A | Limitations<br>(one study small sample size) | Limitations<br>(one study no effect size) | USA<br>Older adults<br>African American participants | Grade III |
| Initial appointment attendance<br>Alvidrez et al. 2005 | One study<br>medium quality | N/A | Limitations<br>(one study small sample size) | Limitations<br>(one study no effect size) | USA<br>Older adults<br>African American participants | Grade III |
| Help seeking intention doctor/GP<br>Teng and Friedman 2009<br>Martin et al. 2022 | One study<br>medium quality<br>One study low quality | Inconsistency<br>(one study increase)<br>(one study no difference) | Limitations<br>(one study small sample size)<br>(one study adequate sample size) | Limitations<br>(two studies no effect sizes) | USA<br>Adolescents aged 14-18 years<br>Black participants<br>USA<br>Older adults<br>Chinese American participants | Grade III |
| Help seeking intention MH professionals<br>Teng and Friedman 2009<br>Martin et al. 2022 | One study<br>medium quality<br>One study low quality | Inconsistency<br>(one study increase)<br>(one study no difference) | Limitations<br>(one study small sample size)<br>(one study adequate sample size) | Limitations<br>(two studies no effect sizes) | USA<br>Adolescents aged 14-18 years<br>Black participants<br>USA<br>Older adults<br>Chinese American participants | Grade III |
| Help seeking intentions regarding dementia<br>Mukadam et al. 2018 | One study<br>medium quality | N/A | Limitations<br>(one study small sample size) | Limitations<br>(one study no effect size) | UK<br>Adults >50 years<br>South Asian participants | Grade III |
| **Interventions focusing on change at the organisational level** |  |  |  |  |  |  |
| Number days until first visit<br>Arean et al. 2008 | One study high quality | N/A | No limitations | No limitations | USA<br>Older adults (>65 years)<br>Ethnic minority groups | Grade II |
| Initial appointment attendance<br>Arean et al. 2008 | One study high quality | N/A | No limitations | No limitations | USA<br>Older adults (>65 years)<br>Ethnic minority groups | Grade II |
| Ongoing appointment attendance<br>Arean et al. 2008 | One study high quality | N/A | No limitations | No limitations | USA<br>Older adults (>65 years)<br>Ethnic minority groups | Grade II |
| Depression<br>Arean et al. 2008 | One study high quality | N/A | No limitations | No limitations | USA<br>Older adults (>65 years)<br>Ethnic minority groups | Grade II |
| Anxiety<br>Arean et al. 2008 | One study high quality | N/A | No limitations | No limitations | USA<br>Older adults (>65 years)<br>Ethnic minority groups | Grade II |
| Referrals<br>Evans et al. 2014 | One study medium quality | N/A | Limitations<br>(one study adequate sample size) | Limitations<br>(one study no effect size) | UK<br>Adults<br>Black and Minority Ethnic | Grade III |
| Screening<br>Gorman et al 2021 | One study high quality | N/A | No limitations | No limitations | USA<br>Adults<br>Ethnic minority groups | Grade II |
| **Interventions focusing on change on change at both interpersonal and organisational levels** |  |  |  |  |  |  |
| Service use<br>Ortega and Rosenbeck 2002 | One study medium quality | N/A | Limitations<br>(one study adequate sample size) | Limitations<br>(one study no effect size) | USA<br>Adults (Homeless clients)<br>Hispanic | Grade III |
| Clinical outcomes<br>Ortega and Rosenheck 2002 | One study medium quality | N/A | Limitations<br>(one study adequate sample size) | Limitations<br>(one study no effect size) | USA<br>Adults (Homeless clients)<br>Hispanic | Grade III |
| Psychosis symptoms<br>Ortega and Rosenheck 2002 | One study medium quality | N/A | Limitations<br>(one study adequate sample size) | Limitations<br>(one study no effect size) | USA<br>Adults (Homeless clients)<br>Hispanic | Grade III |
| Referral<br>Yeung et al. 2004 | One study medium quality | N/A | Limitations<br>(one study small sample size) | Limitations<br>(one study no effect size) | USA<br>Adults<br>Chinese Americans | Grade III |
| Initial appointment attendance<br>Yeung et al. 2004 | One study medium quality | N/A | Limitations<br>(one study small sample size) | Limitations<br>(one study no effect size) | USA<br>Adults<br>Chinese Americans | Grade III |
| **Interventions focusing on change on change at both community and organisational levels** |  |  |  |  |  |  |
| Referral<br>Hackett et al. 2009<br>Robidoux et al. 2009 | Two studies low quality | Inconsistency<br>(one study increase, no statistical test)<br>(one study no difference) | Limitations<br>(one study small sample size)<br>(one study adequate sample size) | Limitations<br>(two studies no effect sizes) | UK<br>Adults<br>Pakistani participants<br>USA<br>Mothers | Grade III |
|  |  |  |  |  | Latinx |  |
| Service use<br>Hackett et al. 2009 | One study low quality | N/A | Limitations<br>(one study small sample size) | Limitations<br>(one study no effect sizes) | UK<br>Adults<br>Pakistani participants | Grade III |
| Length of stay<br>Hackett et al. 2009 | One study low quality | N/A | Limitations<br>(one study small sample size) | Limitations<br>(one study no effect sizes) | UK<br>Adults<br>Pakistani participants | Grade III |
| Screening<br>Robidoux et al. 2023 | One study low quality | N/A | No limitations | Limitations<br>(one study no effect sizes) | USA<br>Mothers<br>Latinx | Grade III |
| **Interventions focusing on change on change at both individual and organisational levels** |  |  |  |  |  |  |
| Initial appointment attendance<br>Seabrooke and Milne 2009 | One study low quality | N/A | Limitations<br>(one study small sample size) | Limitations<br>(one study no effect sizes) | UK<br>Older Adults<br>Asians | Grade III |
| Referral<br>Seabrooke and Milne 2009 | One study low quality | N/A | Limitations<br>(one study small sample size) | Limitations<br>(one study no effect sizes) | UK<br>Older Adults<br>Asians | Grade III |
| **Interventions focusing on change on change at individual, interpersonal and organisation levels** |  |  |  |  |  |  |
| Initial attendance<br>Michelson and Day 2014<br>Sanchez et al. 2022 | Two studies medium quality | Inconsistency<br>(One study no difference)<br>(One study mixed results) | Limitations<br>(two studies small sample sizes) | Limitations<br>(One study no effect size)<br>(One study effect sizes calculated) | UK<br>Primary parental caregivers of all children and adolescents<br>Black and Minority Ethnic<br>USA<br>Primary caregivers of children aged 2-7 years<br>Ethnic minority groups<br>(Hispanic/Latinx, Haitian, or Not Hispanic or Haitian) | Grade III |
| Ongoing attendance<br>Michelson and Day<br>Sanchez et al. 2022 | Two studies medium quality | Inconsistency<br>(One study no difference)<br>(One study mixed results) | Limitations<br>(two studies small sample sizes) | Limitations<br>(One study no effect size)<br>(One study effect sizes calculated) | UK<br>Primary parental caregivers of all children and adolescents<br>Black and Minority Ethnic<br>USA<br>Primary caregivers of children aged 2-7 years<br>Ethnic minority groups<br>(Hispanic/Latinx, Haitian, or Not Hispanic or Haitian) | Grade III |
| Treatment completion<br>Sanchez et al. 2022 | One study<br>medium<br>quality | N/A | Limitations<br>(one study small<br>sample sizes) | No limitations | USA<br>Primary caregivers of children<br>aged 2-7 years<br>Ethnic minority groups<br>(Hispanic/Latinx, Haitian, or Not<br>Hispanic or Haitian) | Grade III |
| Treatment response<br>Sanchez et al. 2022 | One study<br>medium<br>quality | Inconsistency<br>(One study mixed results) | Limitations<br>(one study small<br>sample sizes) | No limitations | USA<br>Primary caregivers of children<br>aged 2-7 years<br>Ethnic minority groups<br>(Hispanic/Latinx, Haitian, or Not<br>Hispanic or Haitian) | Grade III |

<sup>a</sup> The interaction of client and case manager race was only significant for psychosis symptoms where Hispanic clients of Hispanic case managers had less reduction in psychotic symptoms than other ethnic and racial pairings ( $p < 0.05$ ).
<sup>b</sup> The strength of the overall evidence supporting each conclusion (associated with each outcome) were assessed using the approach used by the Academy of Nutrition and Dietetics (Academy of Nutrition and Dietetics 2022) (see Section 5.8) and Table 8.

The methods used for this rapid review are described in Section 5. This includes the eligibility criteria, which can be found in section 5.1 (Table 4); and the individual elements incorporated in the assessment of the strength of the evidence, which can be found in Section 5.7 (Table 5).

**Table 4:** Eligibility criteria.

|  | Inclusion criteria | Exclusion criteria |
| --- | --- | --- |
| **Participants** | People from ethnic minority groups* of all ages experiencing mental health conditions**<br>Including dementia / Alzheimer's disease and perinatal mental health*** | - International students<br>- Refugees<br>- Immigrants<br>- Asylum seekers<br>- Indigenous populations<br>- All White British ethnicities other than Gypsy or Irish Travellers |
| **Settings** | Primary or secondary care |  |
| **Intervention / exposure** | Interventions aimed at improving equitable or overall access to mental health services (including early access)<br>For example:<br>- Provision of mental health services and therapies<br>- Pathways/routes into mental health support services<br>- Onward referral to specialist mental health services | Interventions for:<br>- Treating mental health conditions<br>- Promoting good mental health<br>- Preventing poor mental health<br>- Improving medication adherence<br>- Engagement in research<br>- Participating, initiating or completing treatment<br>- Cultural adaptations to treatment interventions |
| **Comparison** | Any |  |
| **Outcomes** | Any outcome measure relating to the equitable or overall access of mental health services by ethnic minority groups. For example:<br>- Uptake of the service by ethnic minority groups<br>- Referral rates<br>- Primary care use as a pathway to care by ethnic minorities | - Clinical effectiveness outcomes of mental health treatment<br>- Effectiveness of mental health promotion and prevention interventions<br>- Descriptions of barriers and facilitators |
| **Study design** | Quantitative empirical studies of an intervention with any type of control or comparison group, for example:<br>- randomised control trials<br>- quasi-experimental studies | - Quantitative empirical studies of an intervention without a control group<br>- Qualitative studies<br>- Secondary research |
| **Countries** | No limits |  |
| **Language of publication** | English |  |
| **Publication date** | No limits |  |
| **Publication type** | Published and grey literature | Letters, commentary, editorials, conference abstracts, protocols, theses |

\*Ethnic minority groups of interest include Asian, Asian Welsh or Asian British (i.e., Bangladeshi, Chinese, Indian, Pakistani or any other Asian background), Black, Black Welsh or Black British (i.e., African, Caribbean or any other Black background), Arab, Gypsy, Travellers, Roma, mixed or multiple ethnic groups, or any other minority ethnic groups, excluding White British ethnicities not previously specified.
\*\*For the purposes of searching, this included both common mental disorders (for example, anxiety and depression) and severe mental illnesses (for example bipolar disorder, schizophrenia and schizoaffective disorders, personality disorders, major depressive disorder, obsessive-compulsive disorder, psychosis and psychotic disorders, post-traumatic stress disorder, eating disorders, suicidal behaviour).
\*\*\*While Dementia, Alzheimer's disease and perinatal mental health are not mental health conditions, these are included as they are of interest to policy development.

**Table 5:**
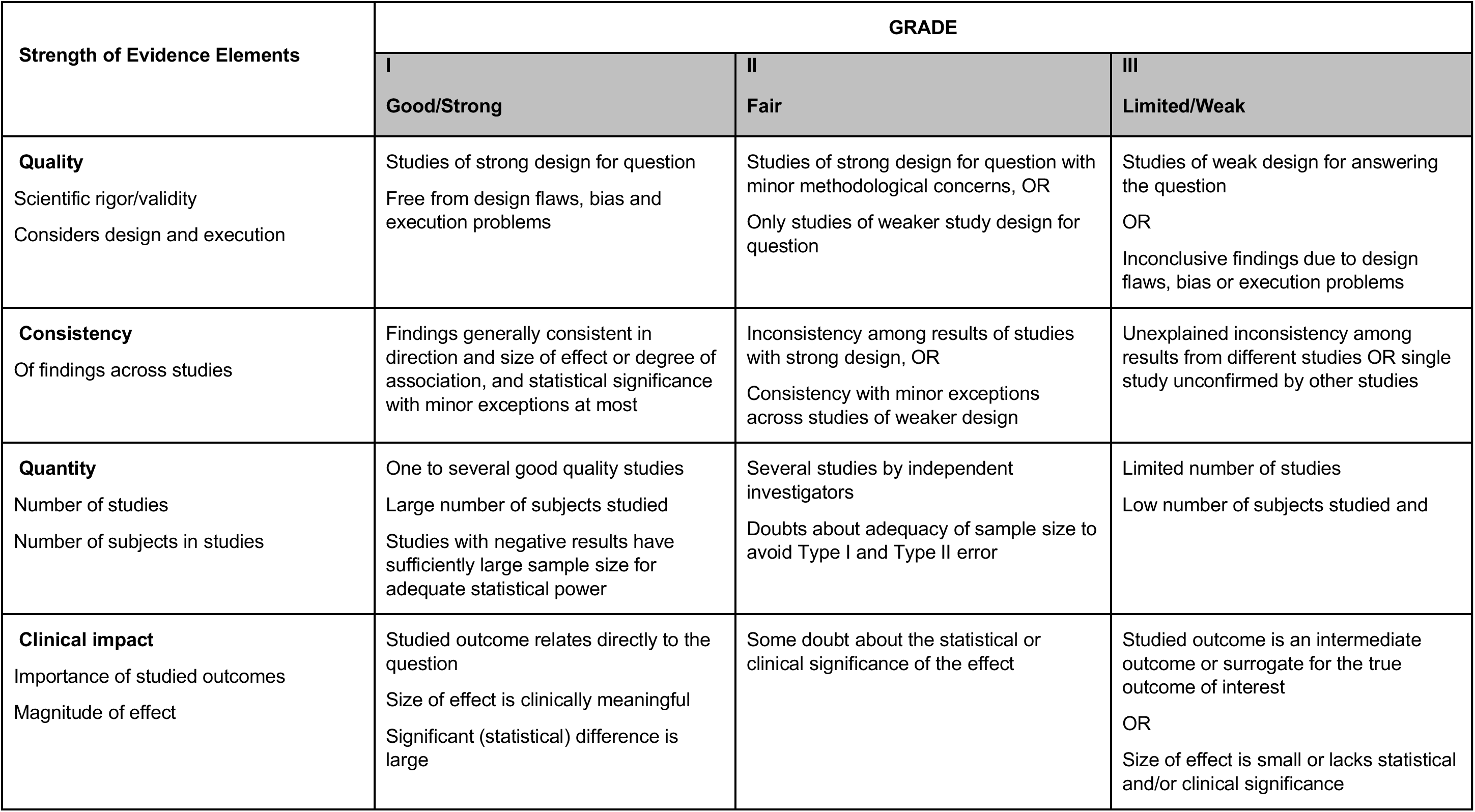
Strength of evidence elements used to assign an overall GRADE level.

### 2.1 Overview of the Evidence Base

The searches identified 3,451 records and after deduplication there were 1,721 records. Following title and abstract screening there were 66 records of which the full papers were retrieved and assessed for inclusion. Fifteen records were found to be relevant and citation searching of these records identified two additional records. Fourteen studies, consisting of seventeen reports^3,4^ were included in this review.

#### Interventions and settings

The 14 studies describe 14 unique interventions delivered by either the research team (n=3), mental health practitioners (n=10) and the co-author of the research report (n=1). To organise the various interventions in a meaningful way an adapted version of the social ecological model (CDC 2022) was used as a framework to identify the levels (individual, intrapersonal, community and organisational) at which interventions aimed to instigate change.

**A summary of the included interventions is provided in Table 1**. A more detailed summary of the interventions evaluated by each included study is also provided in Table 6. (Characteristics of included studies).

**Table 6:**
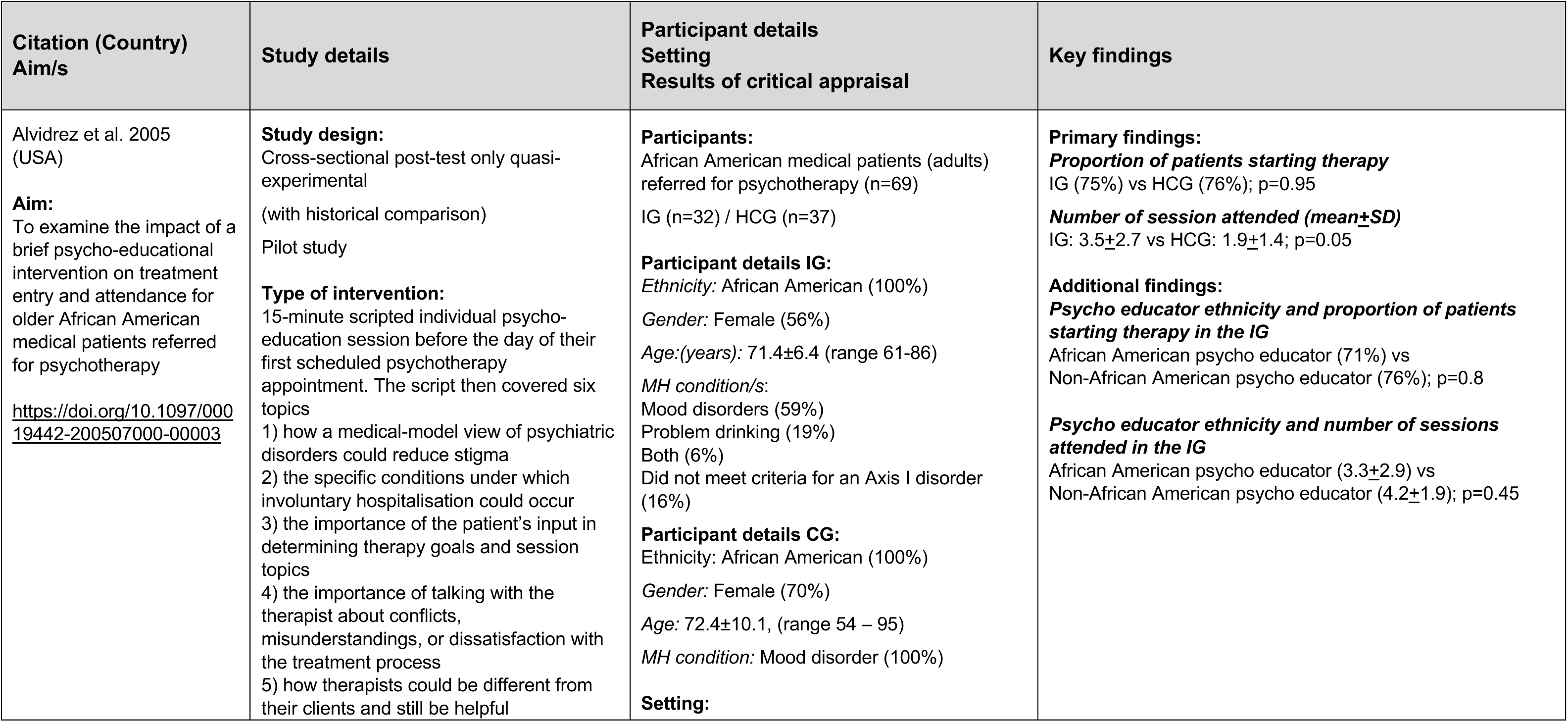
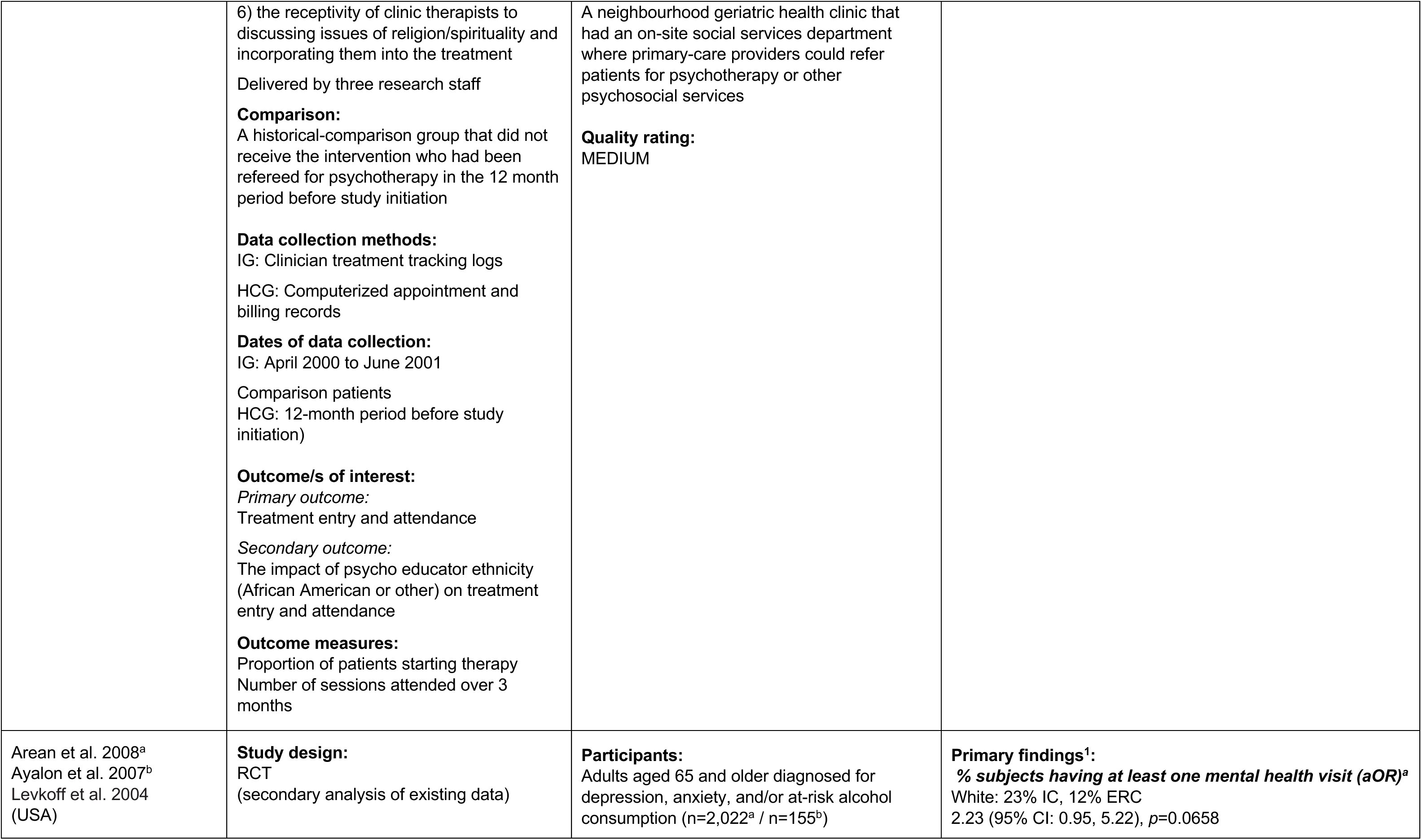
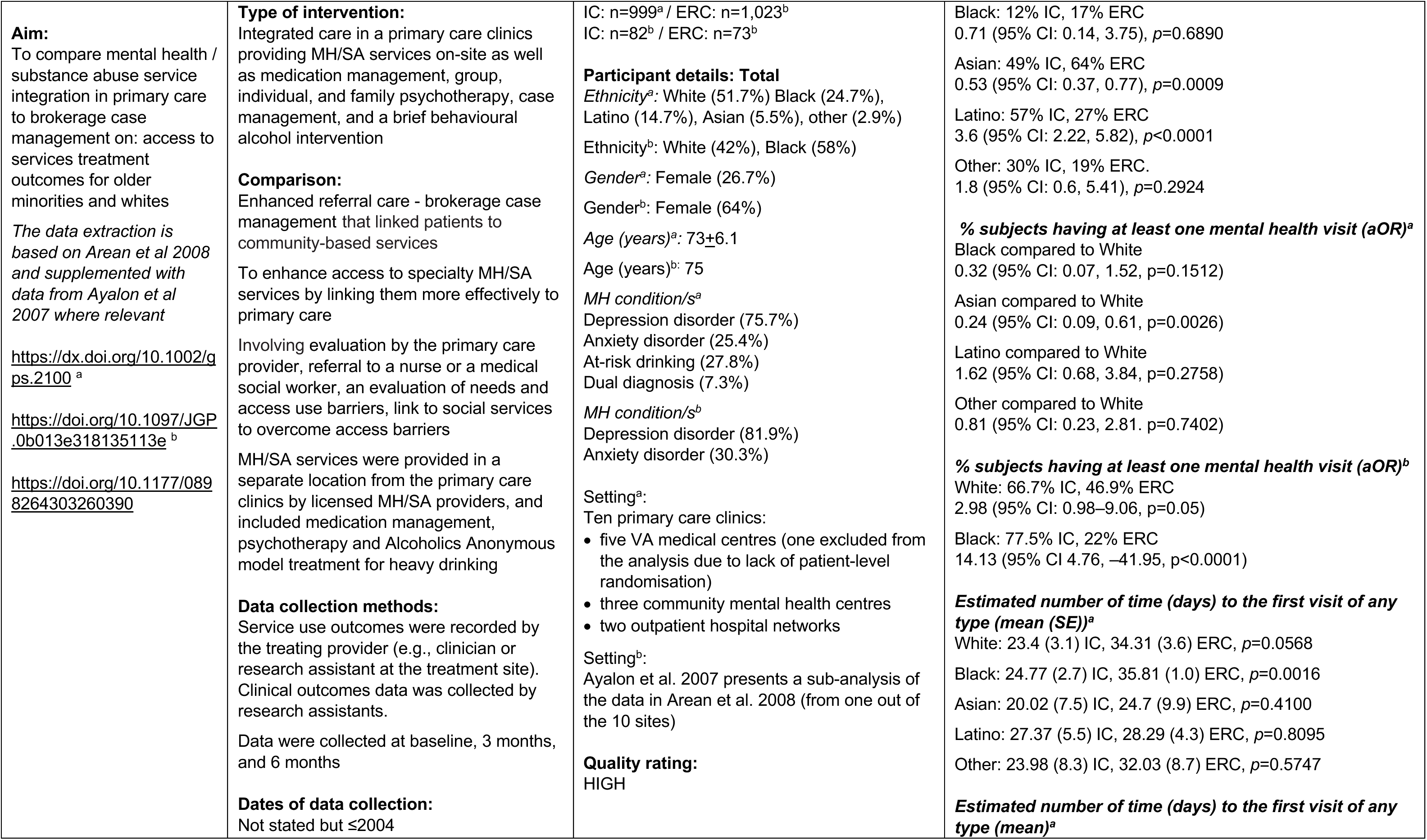
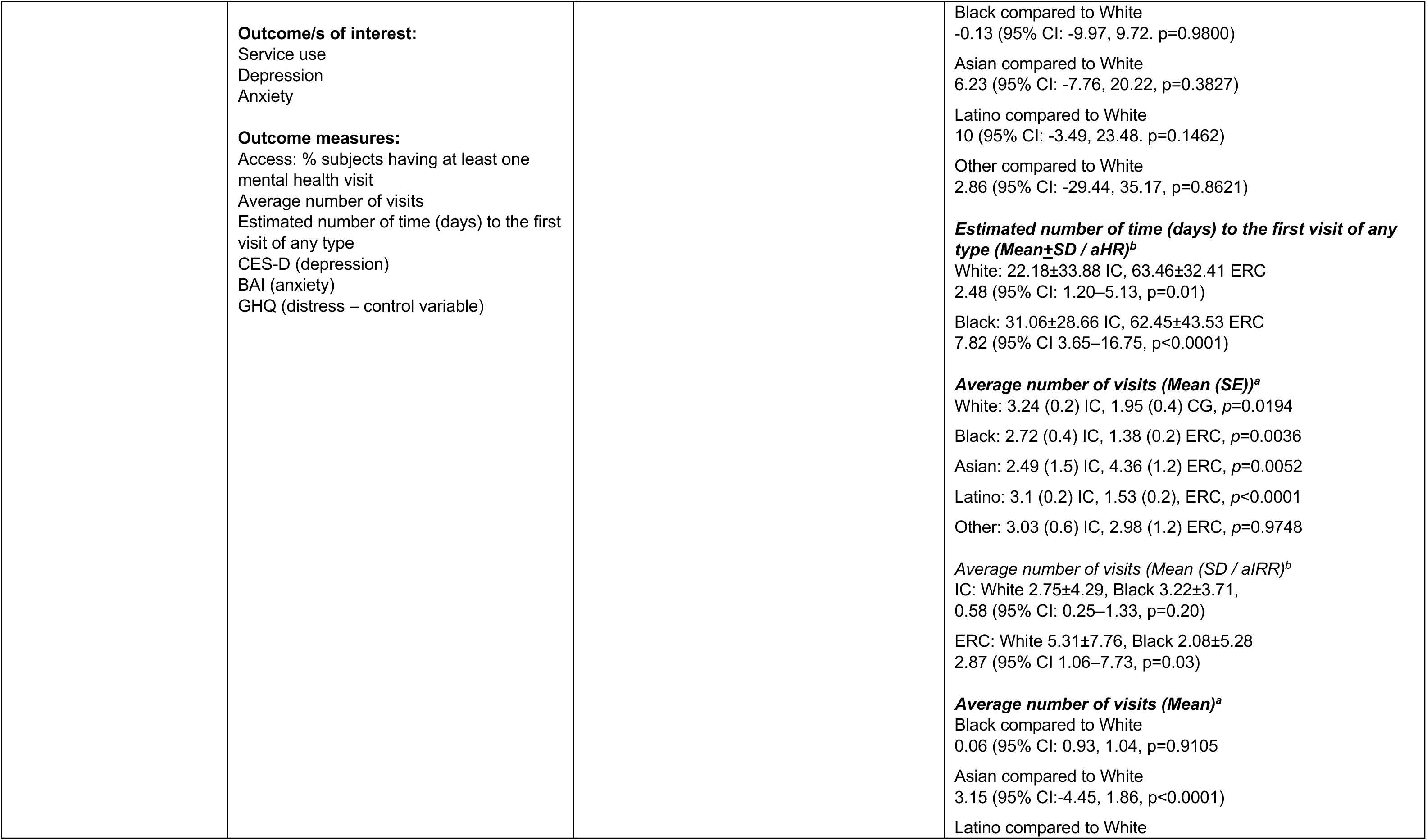
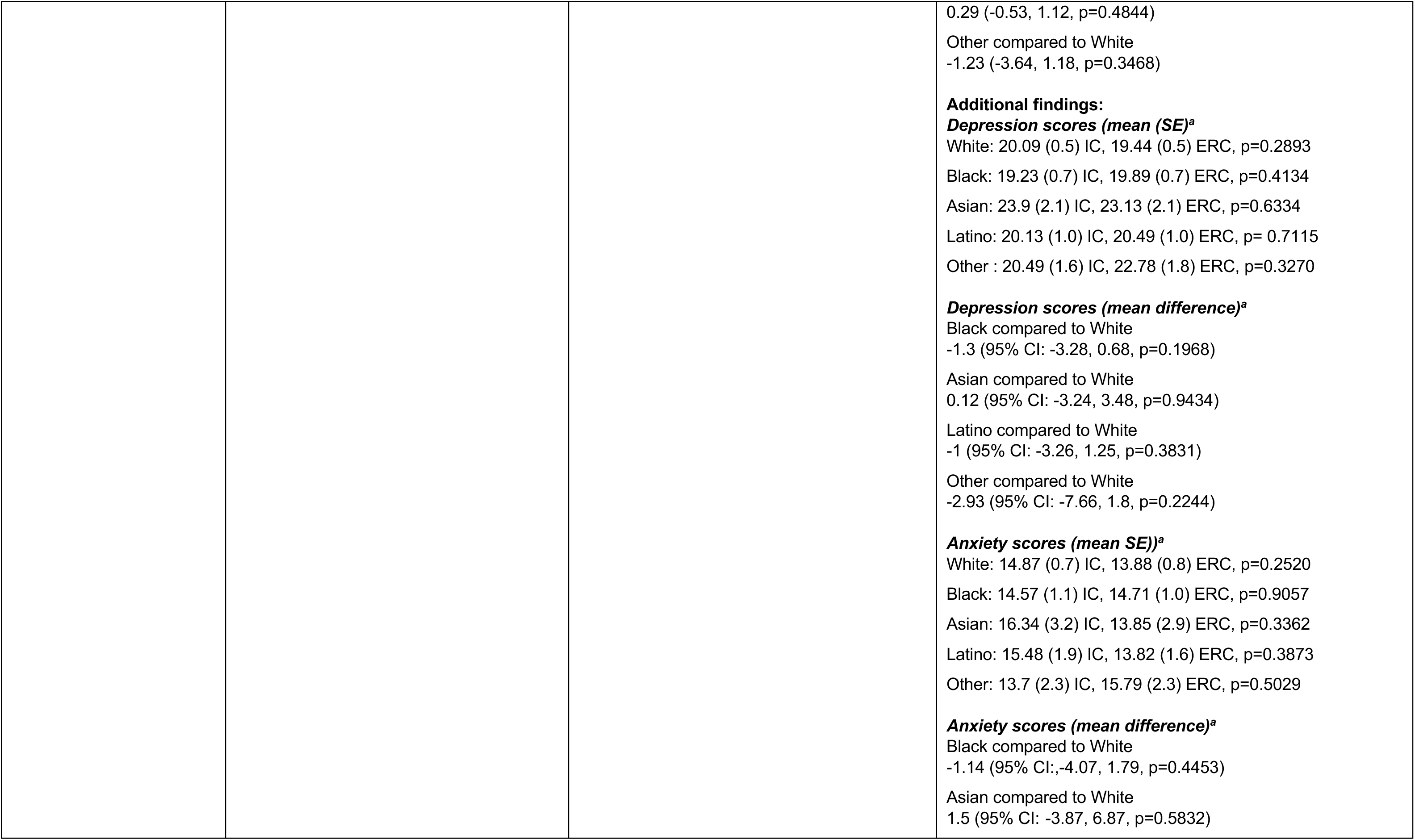
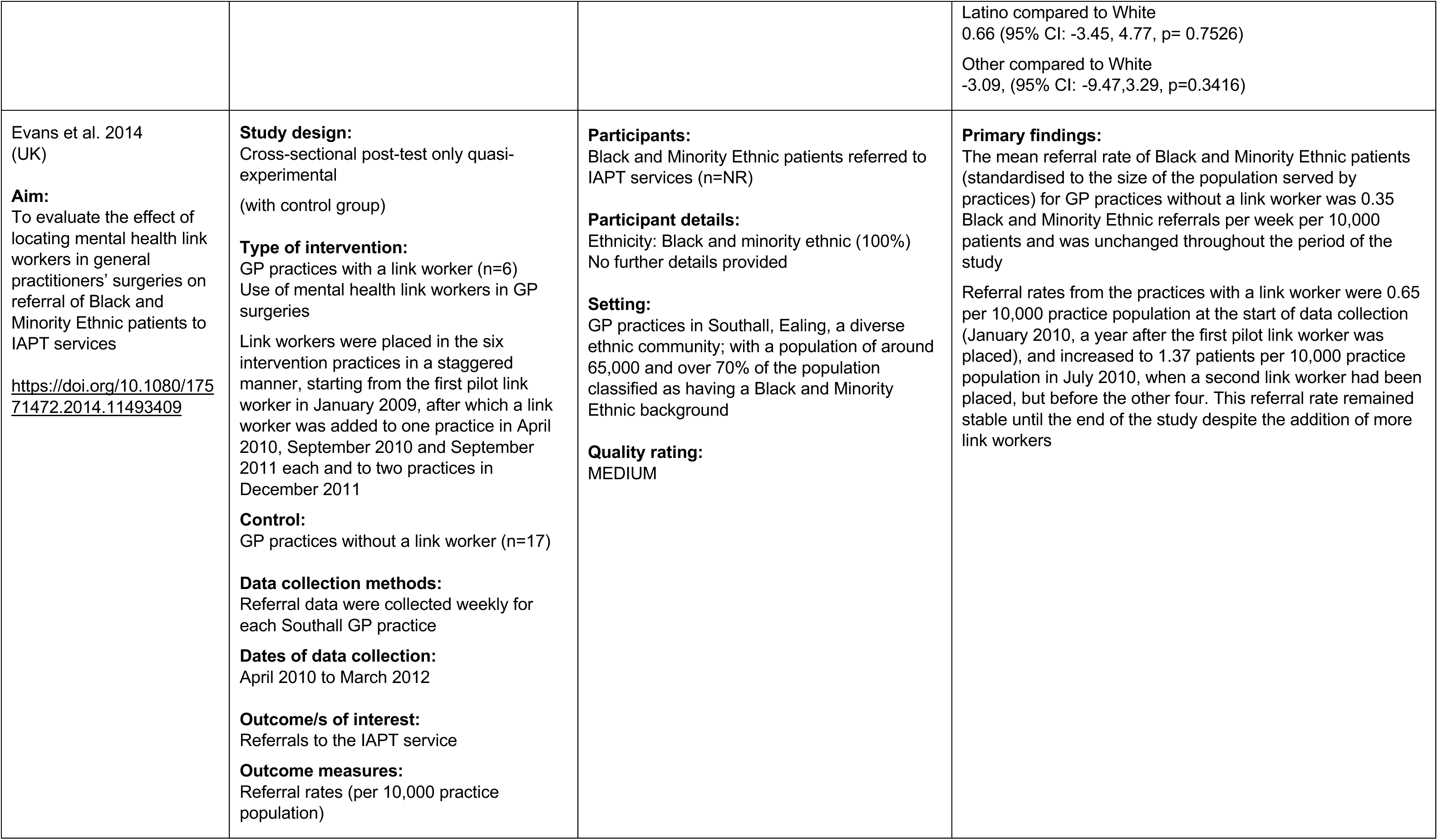
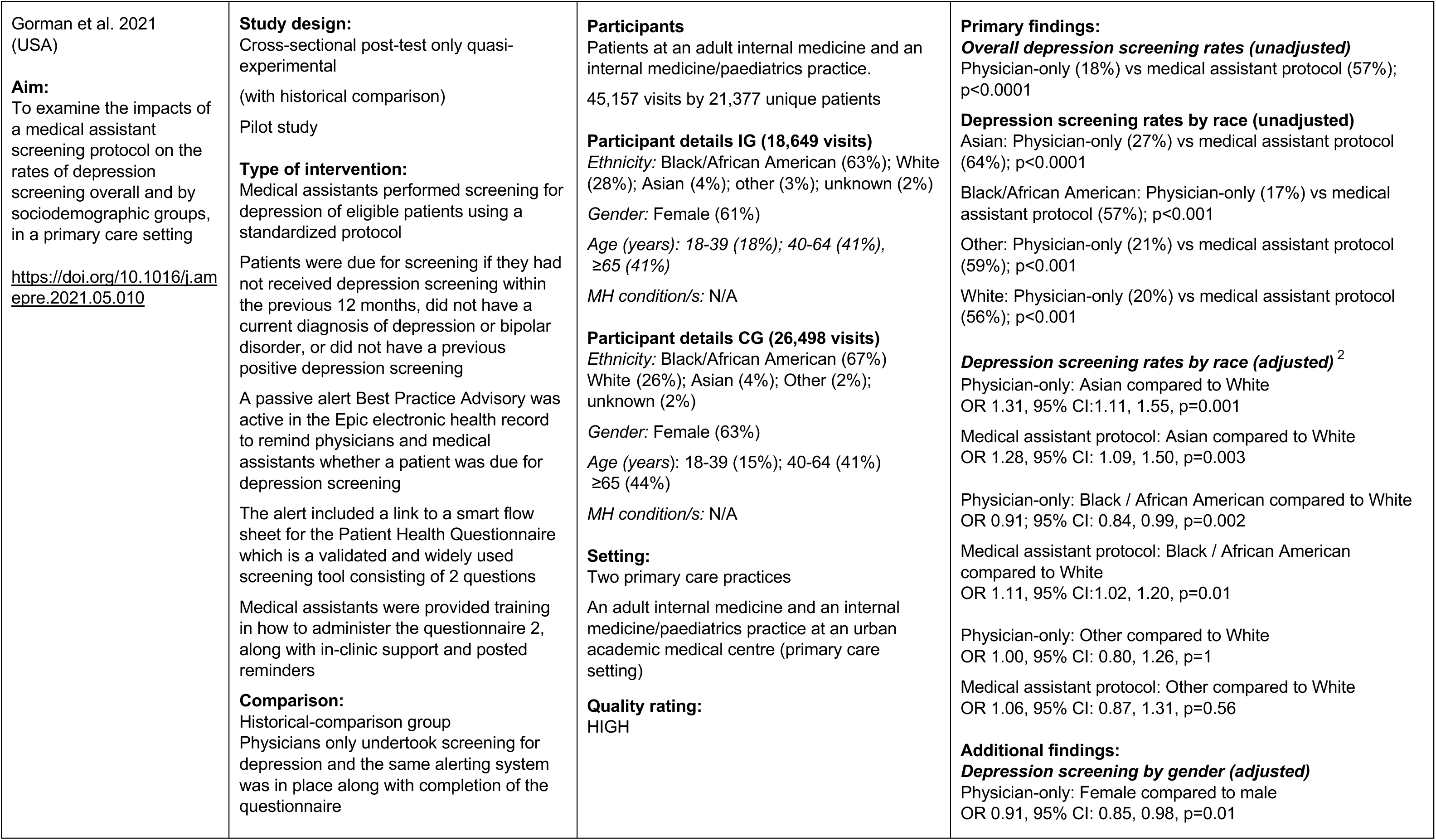
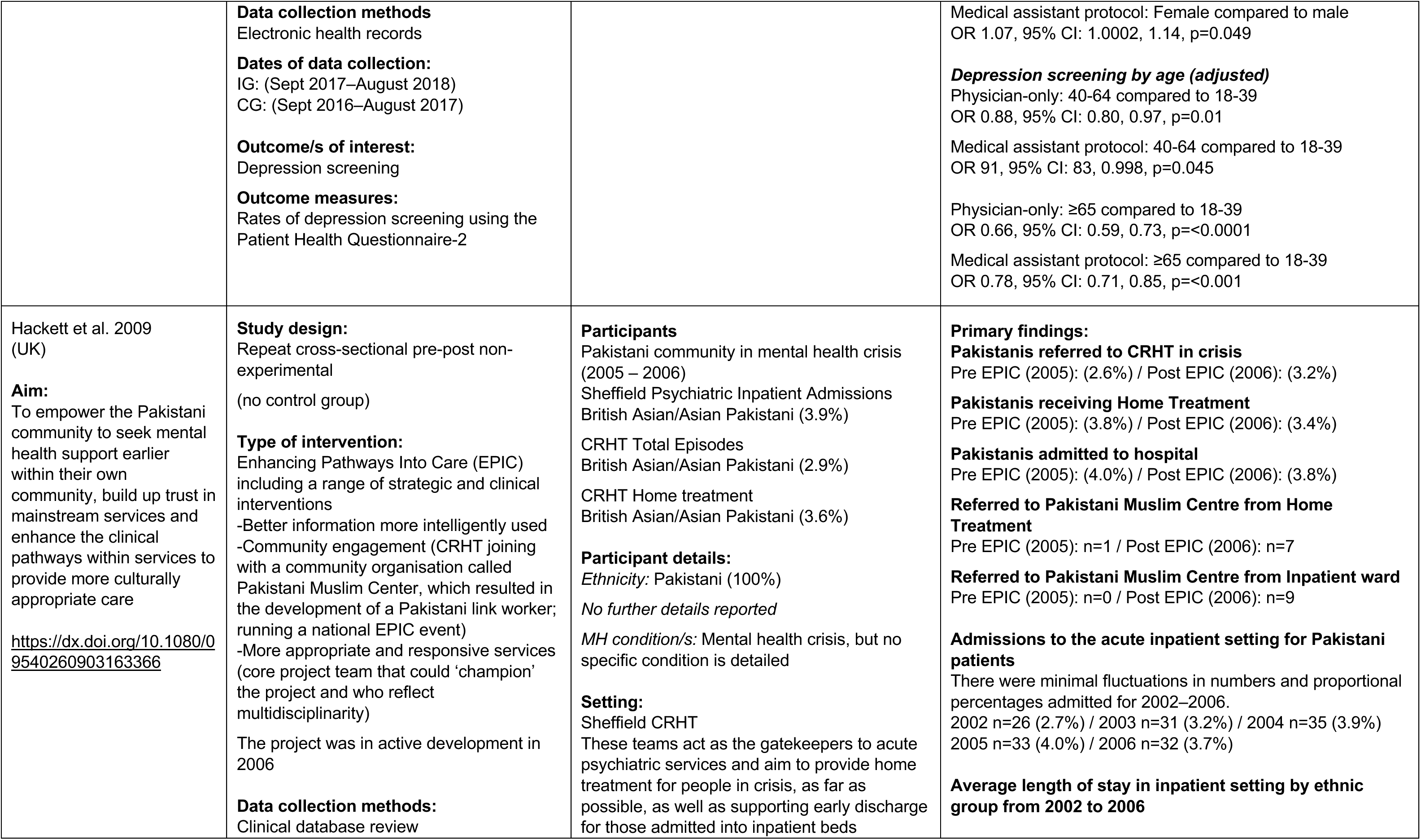
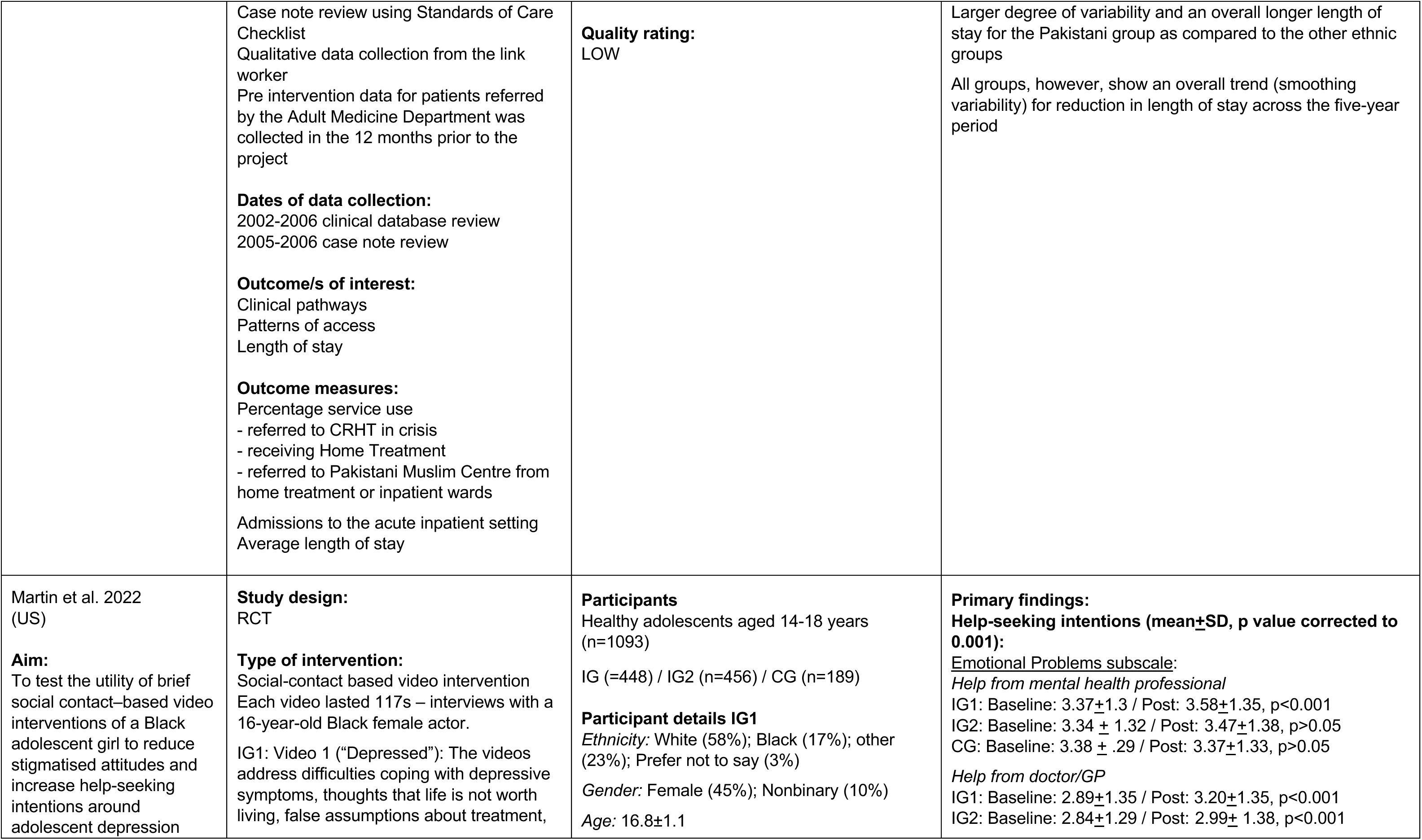
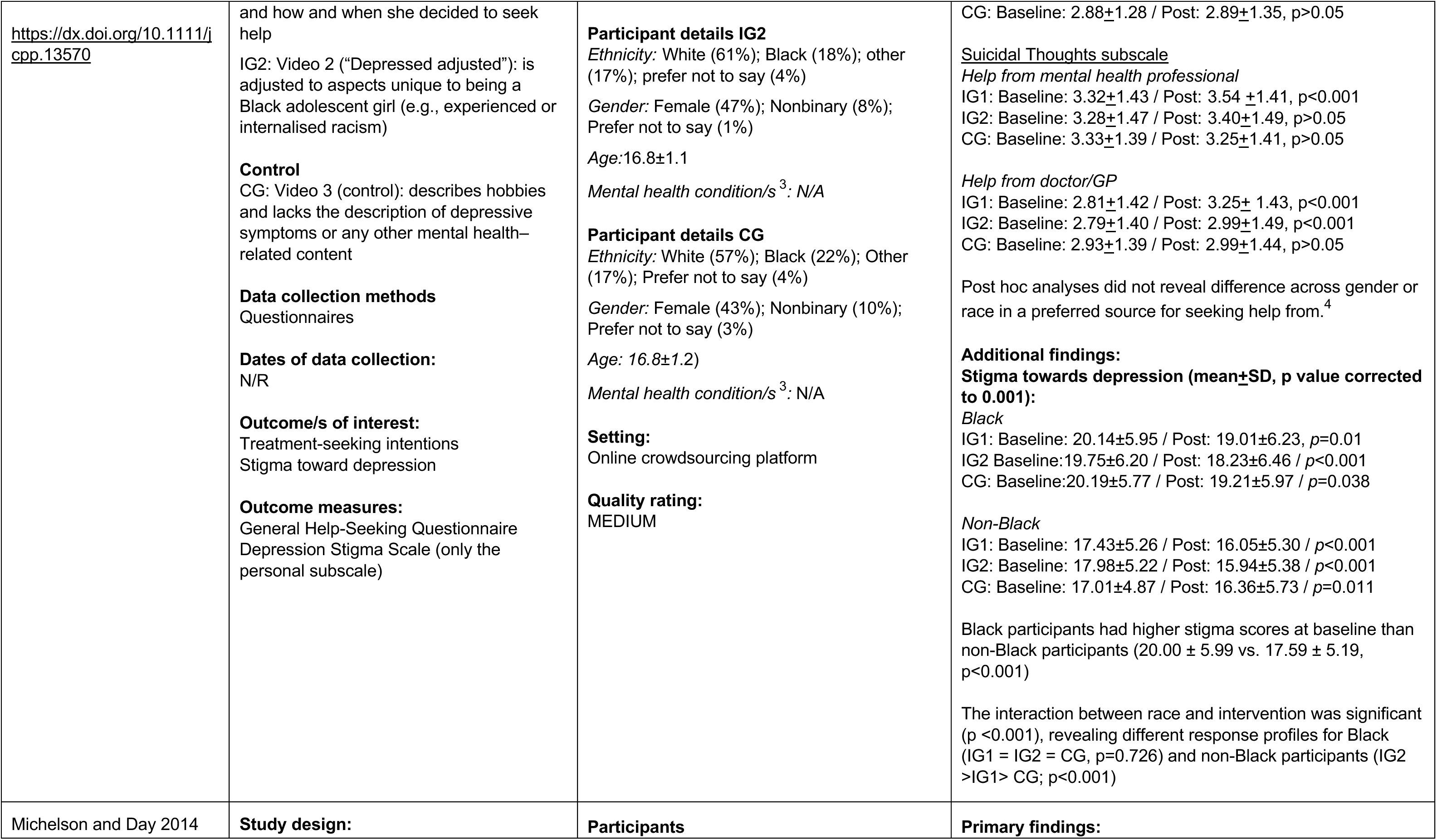
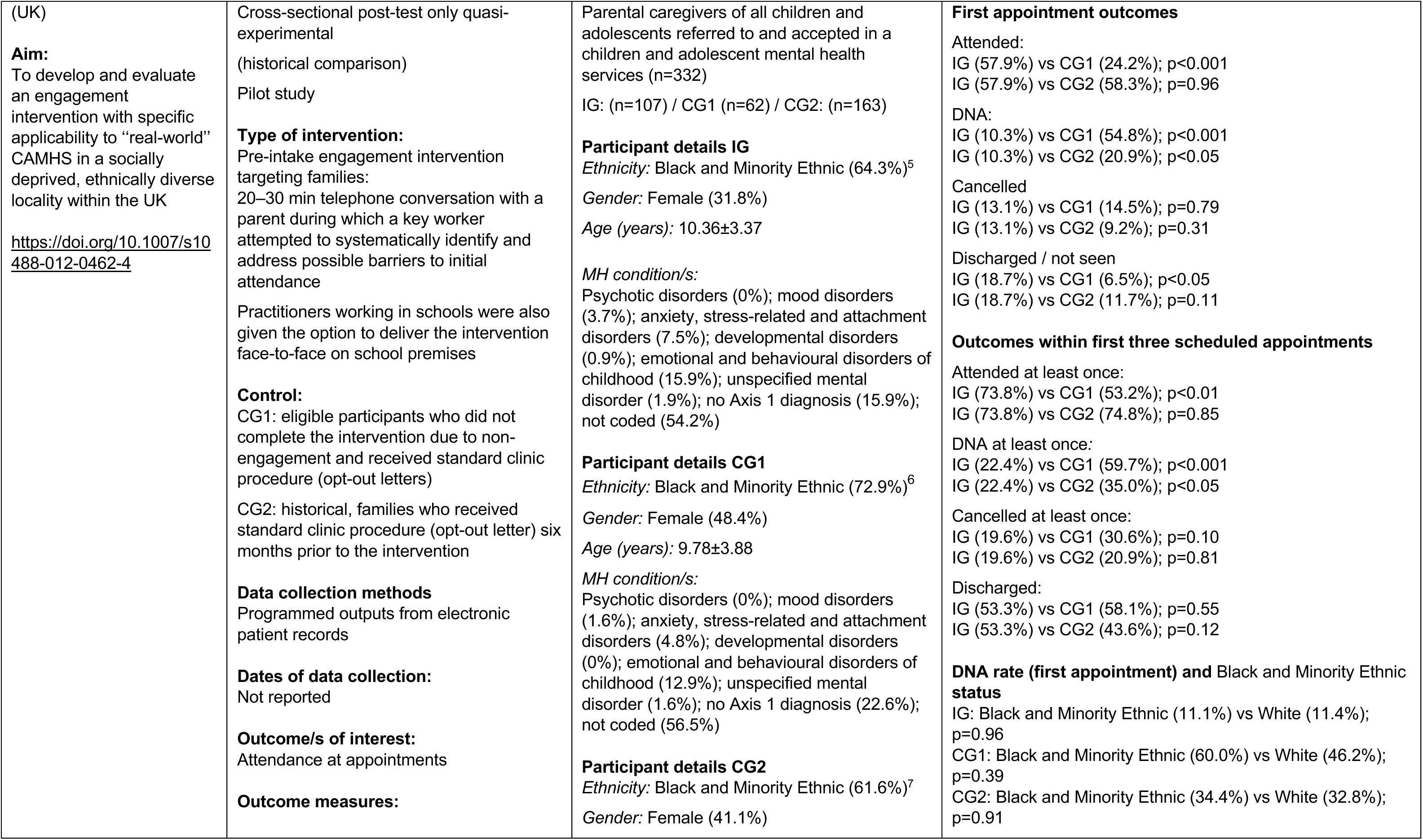
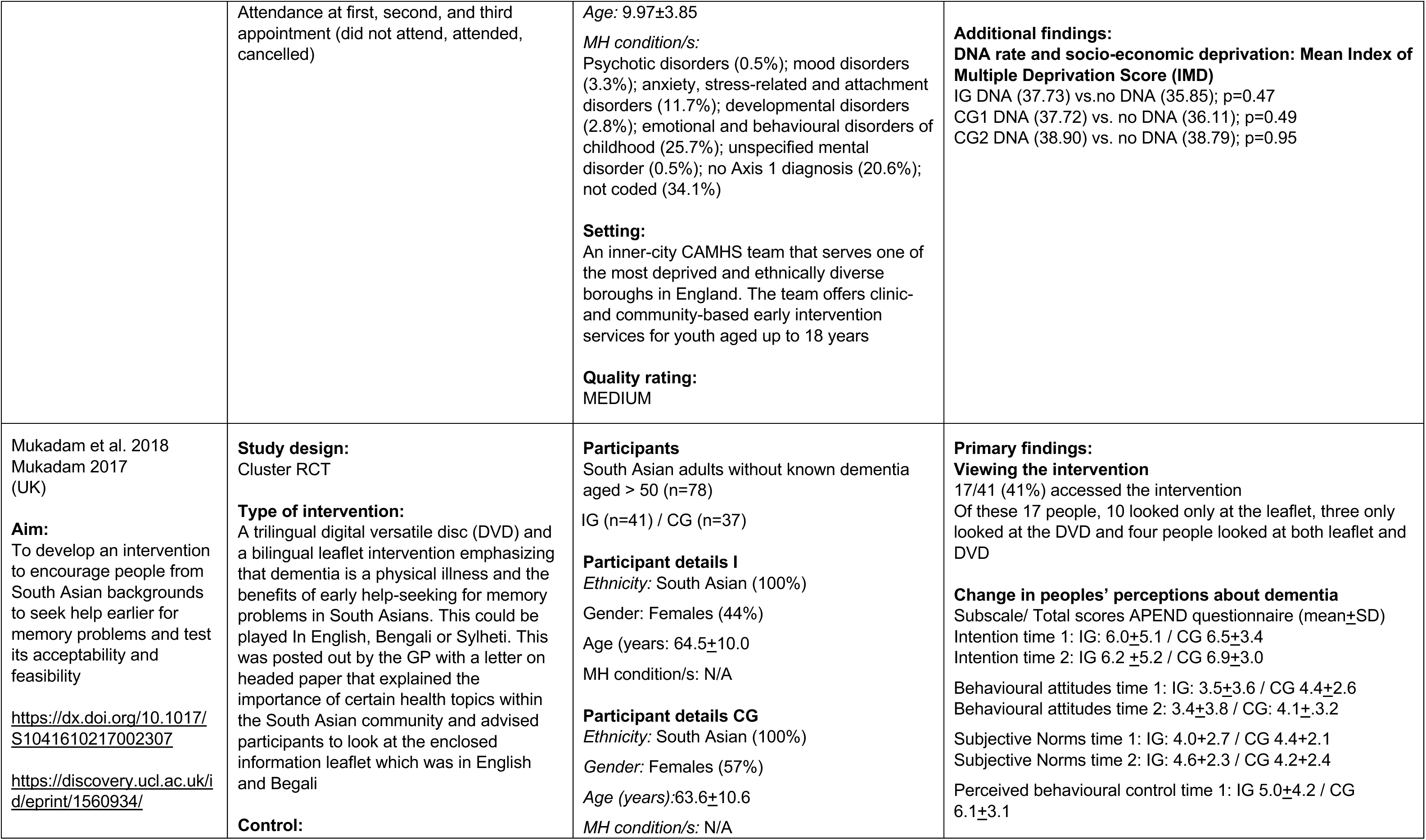
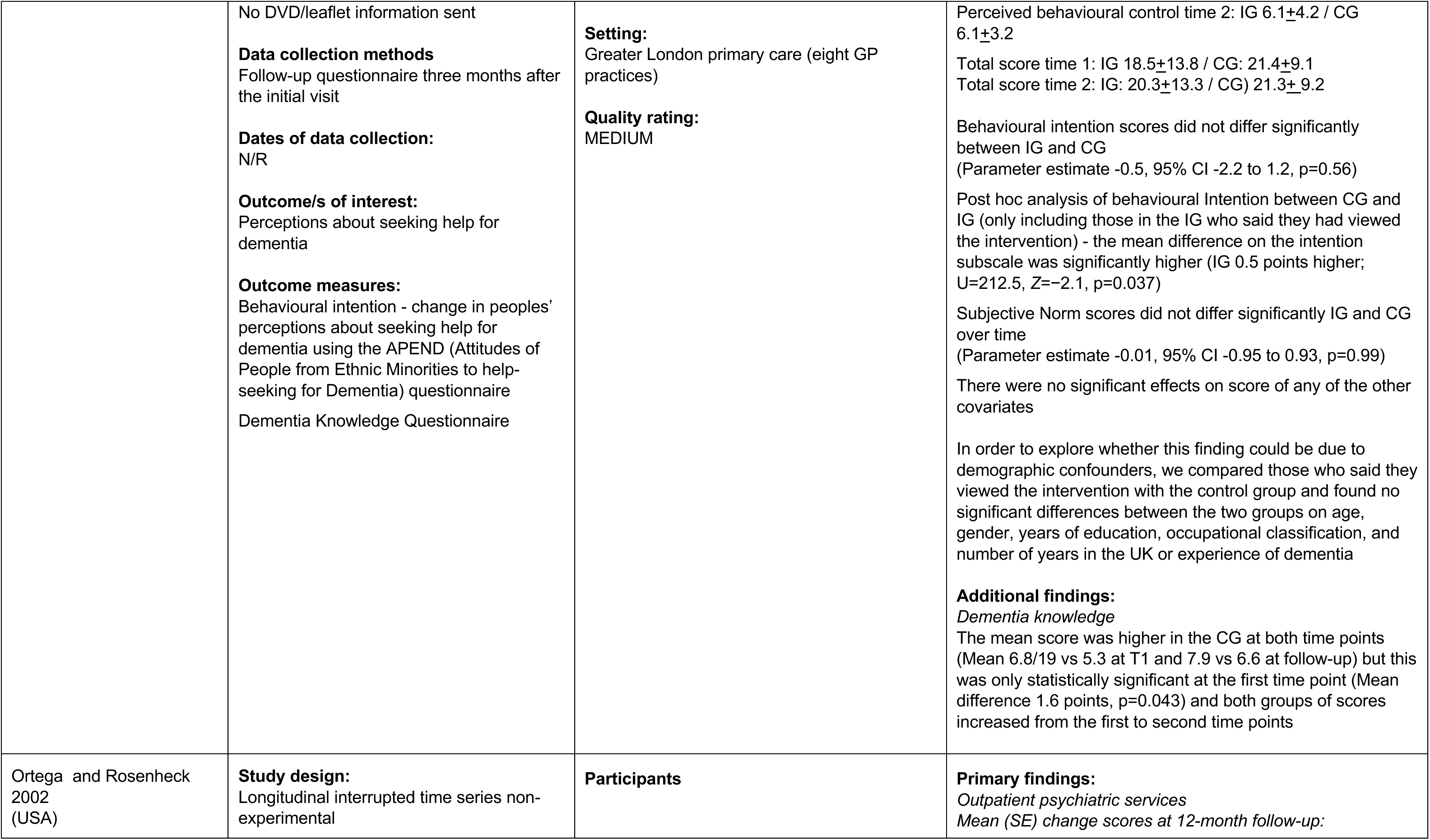
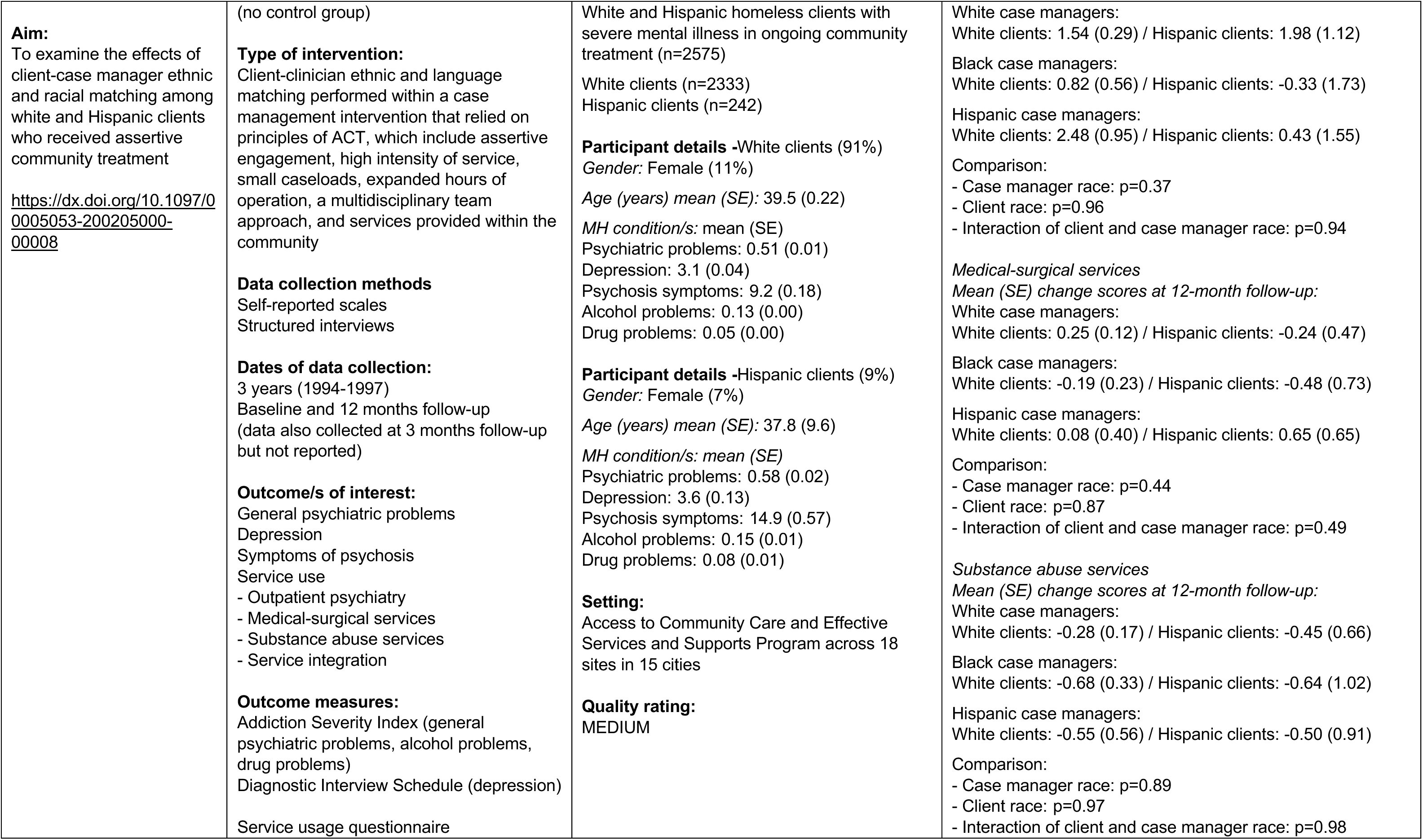
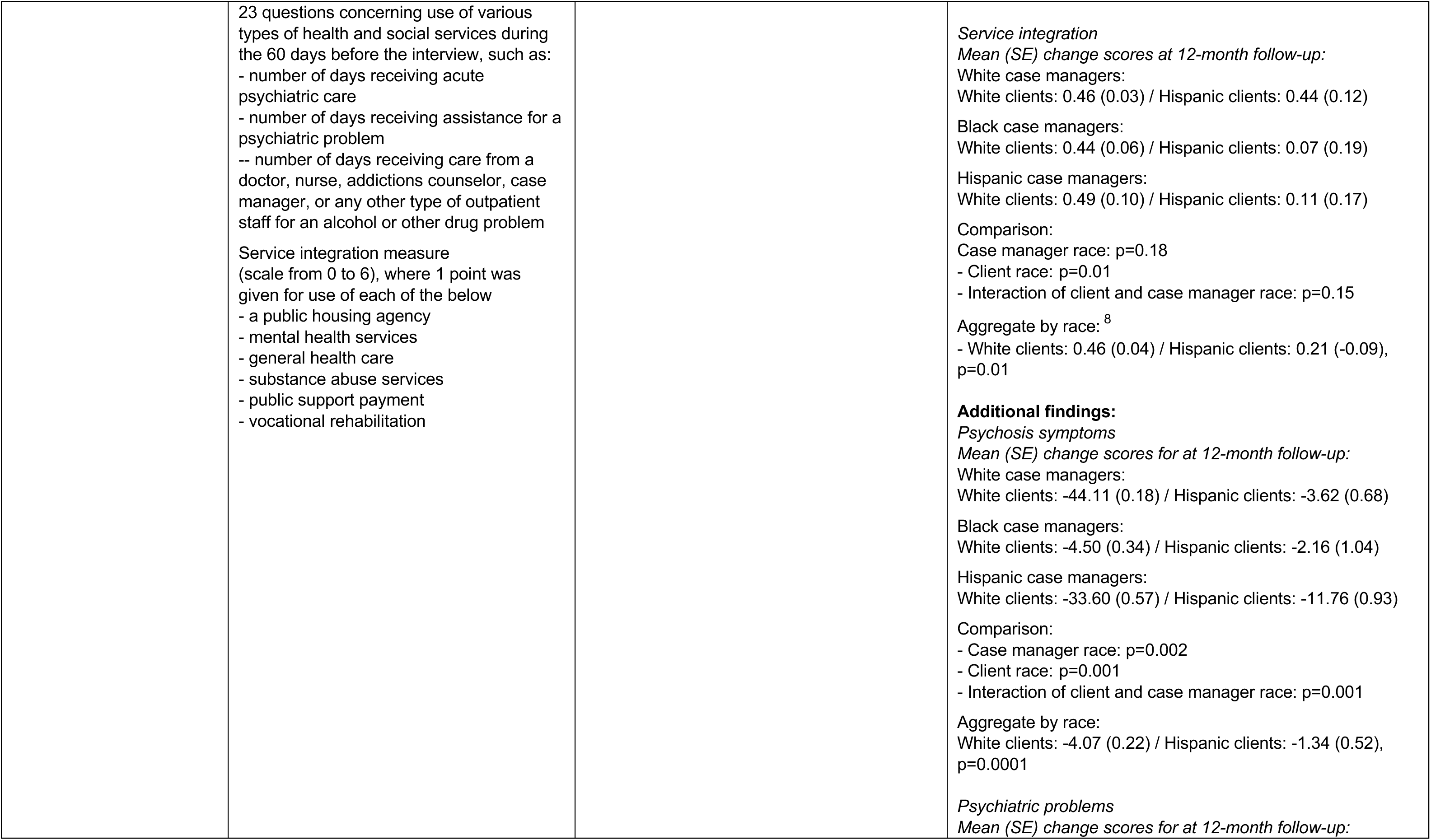
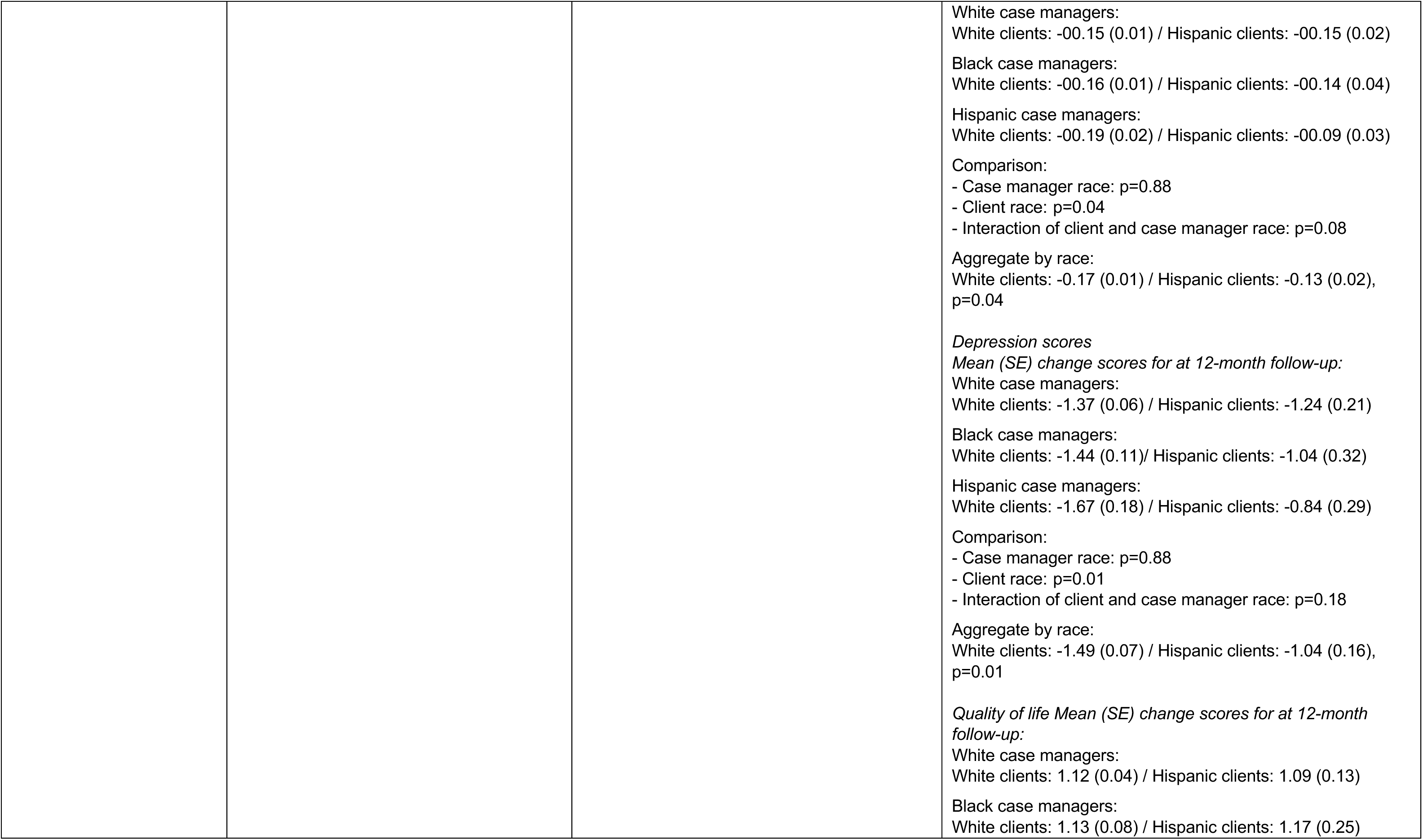
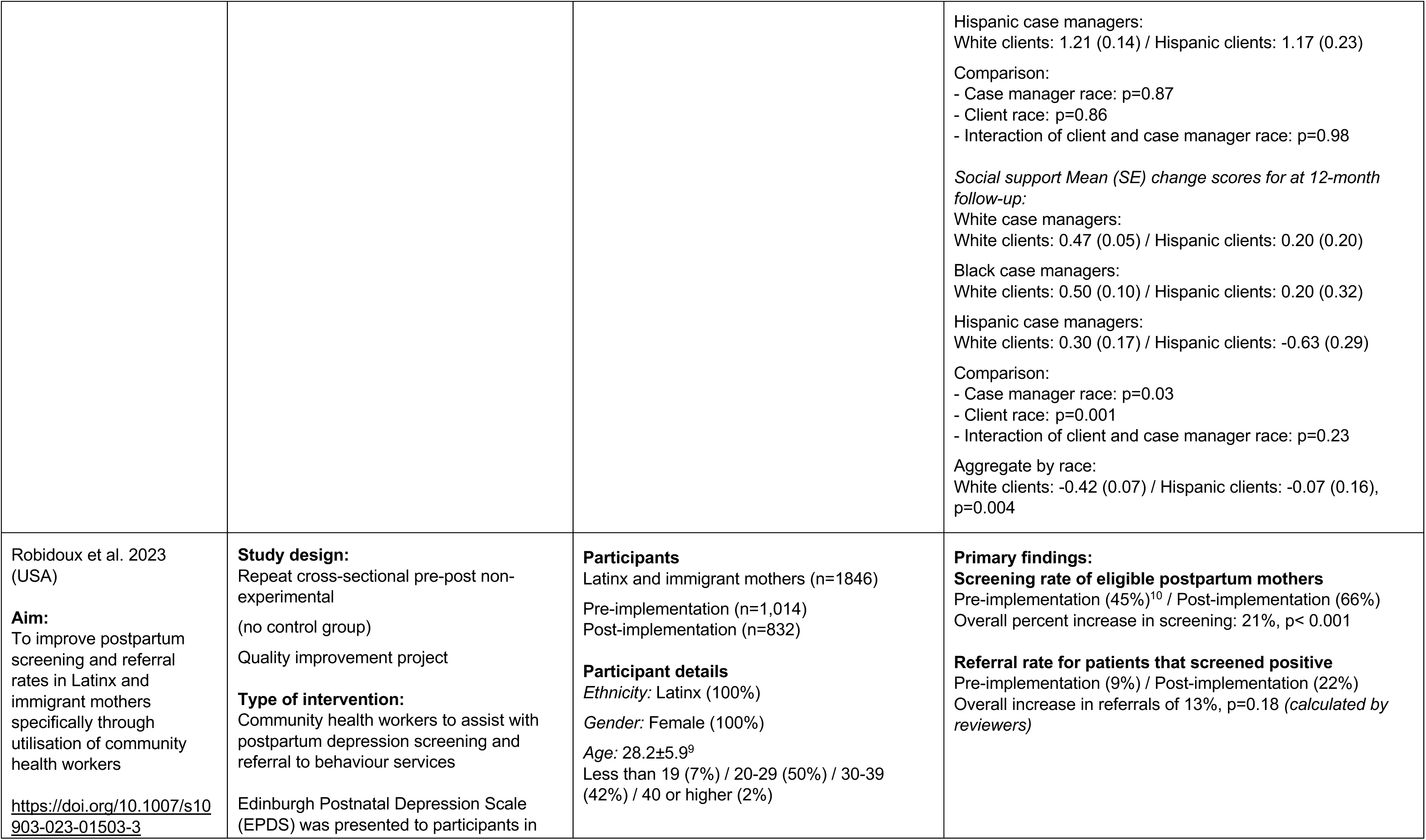
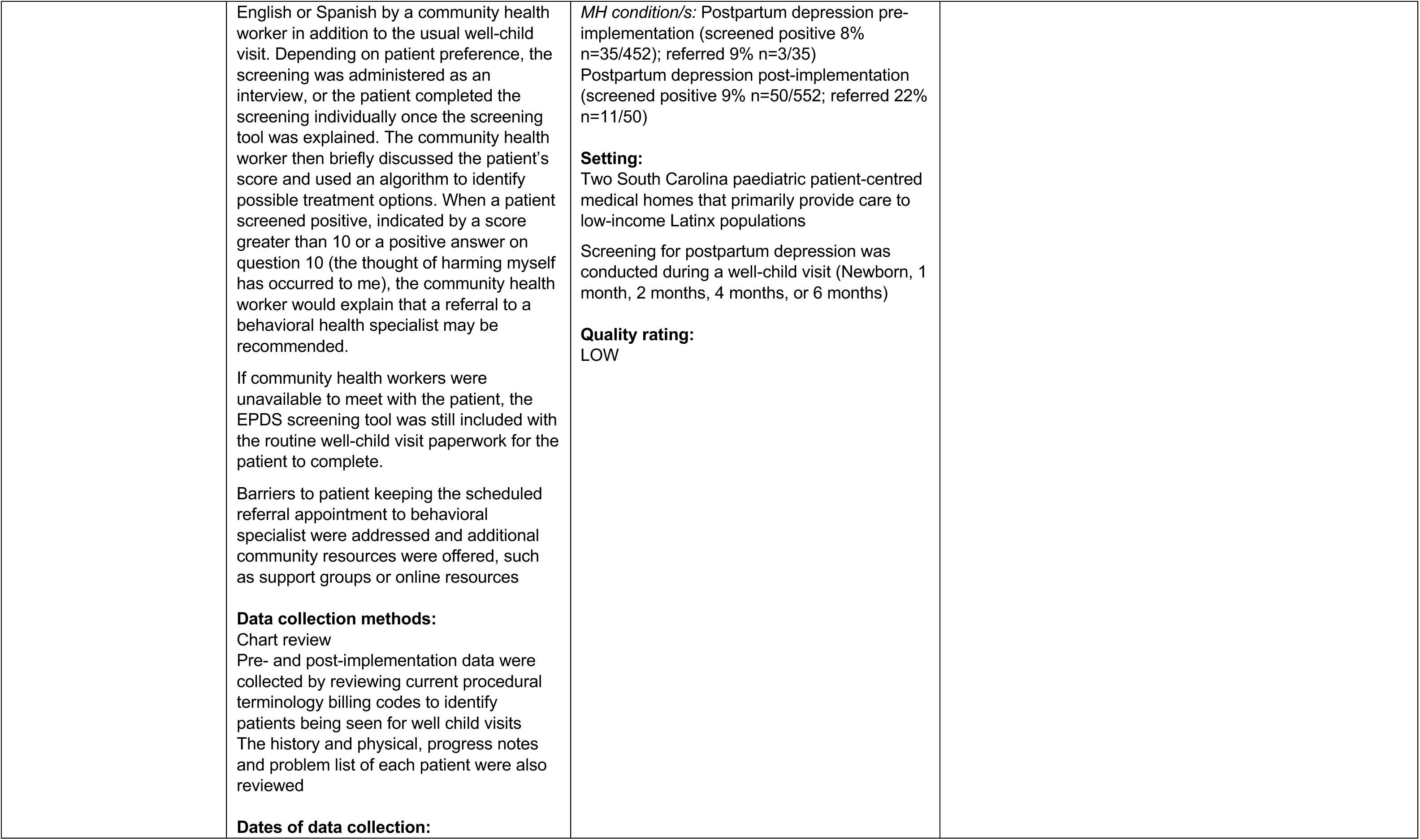
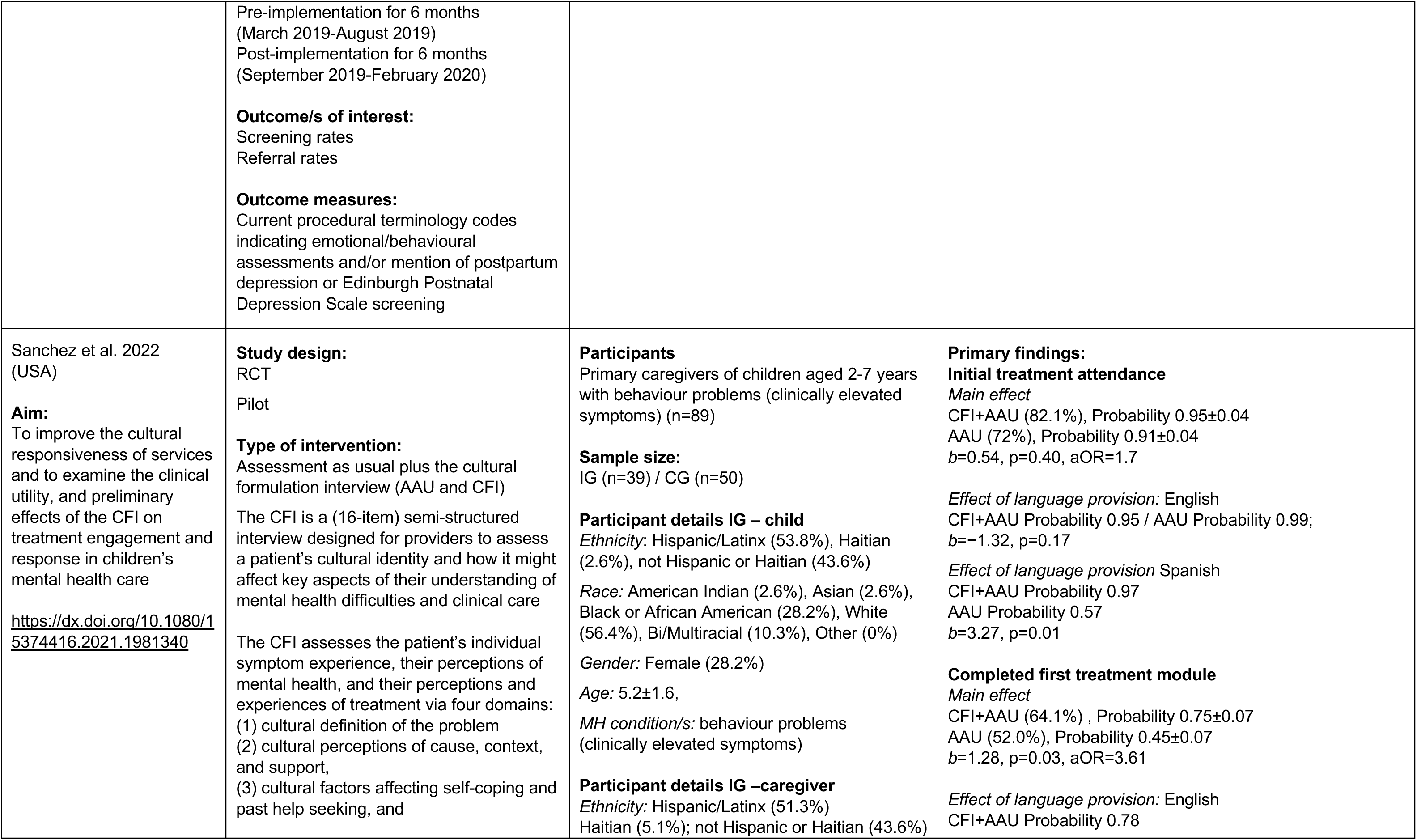
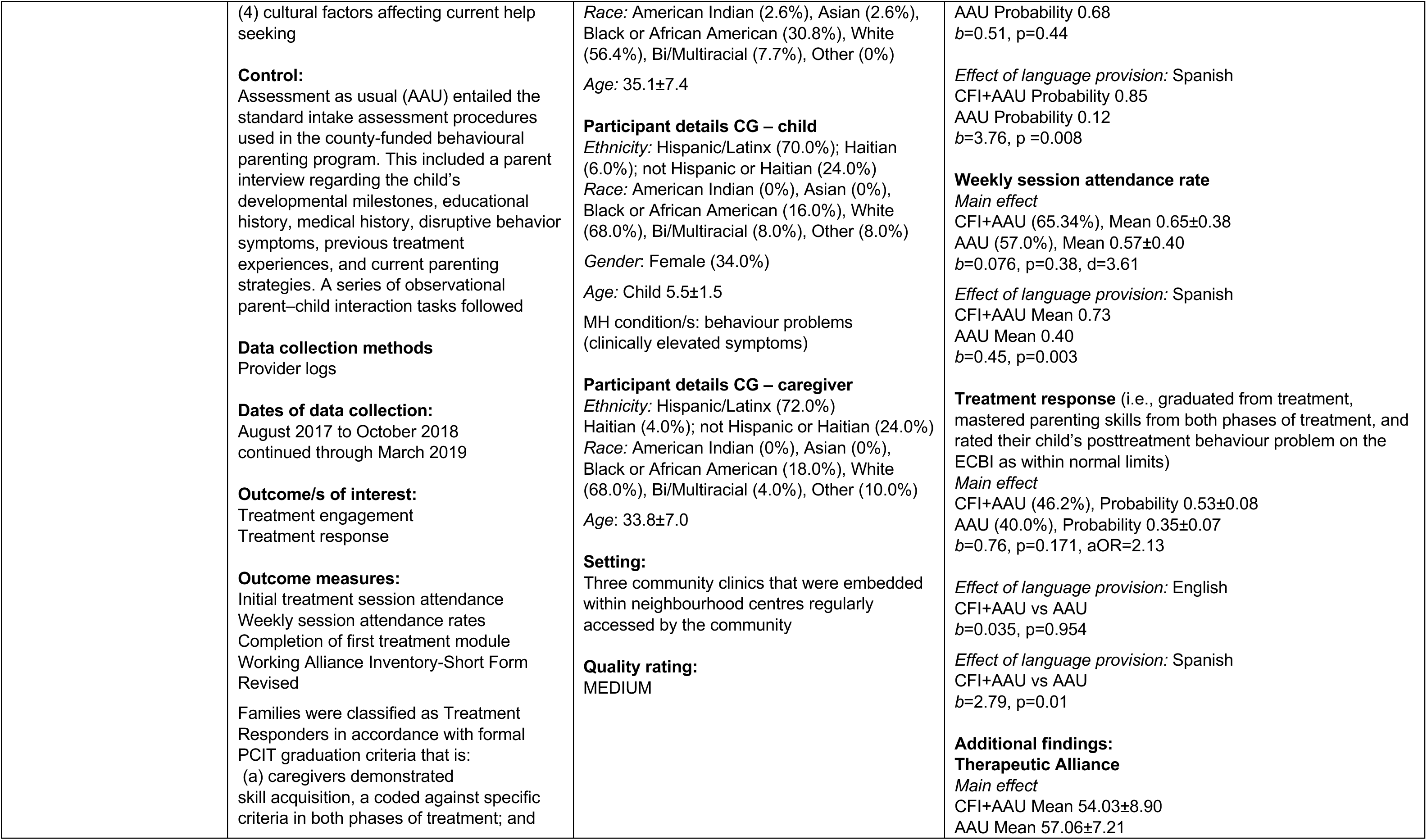
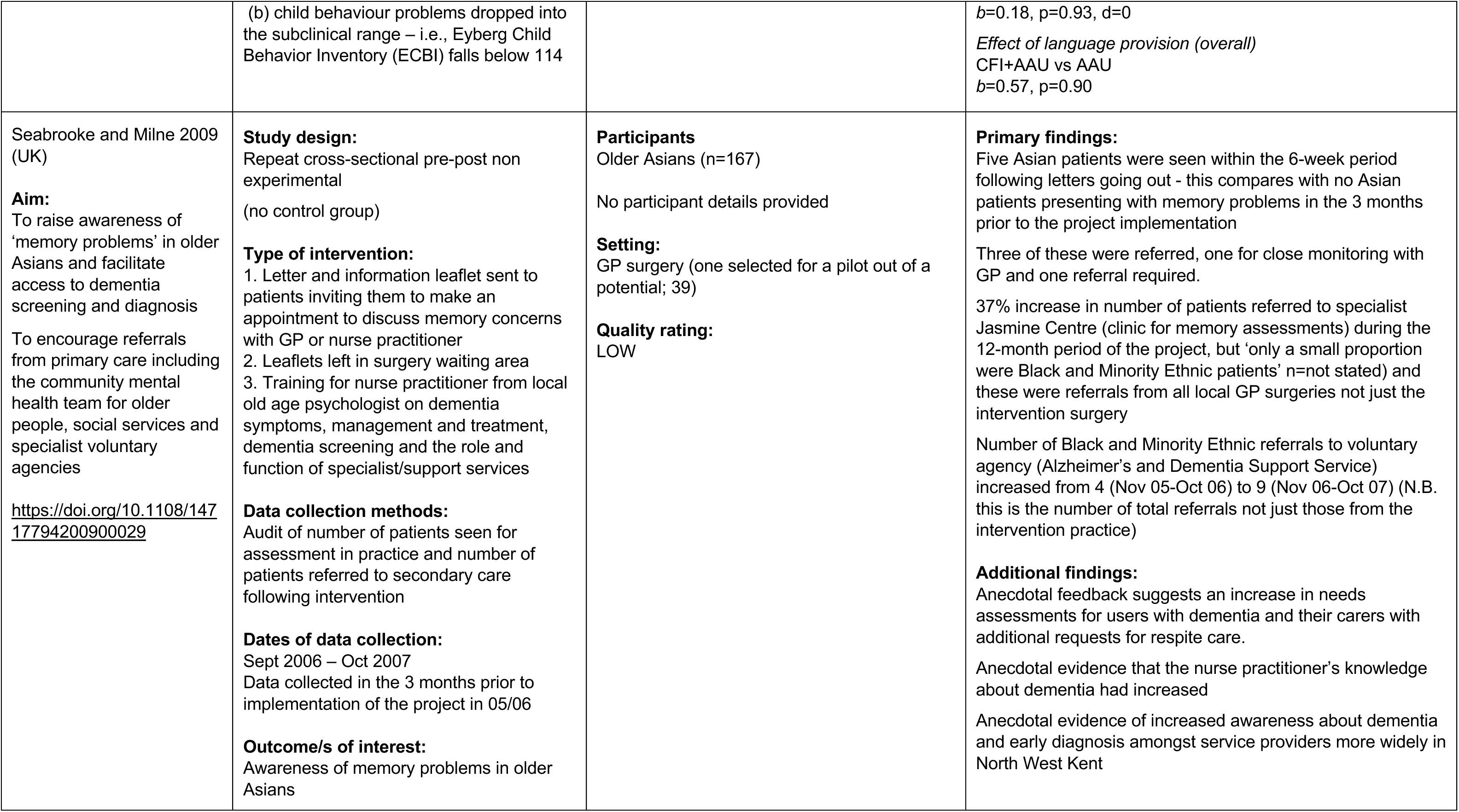
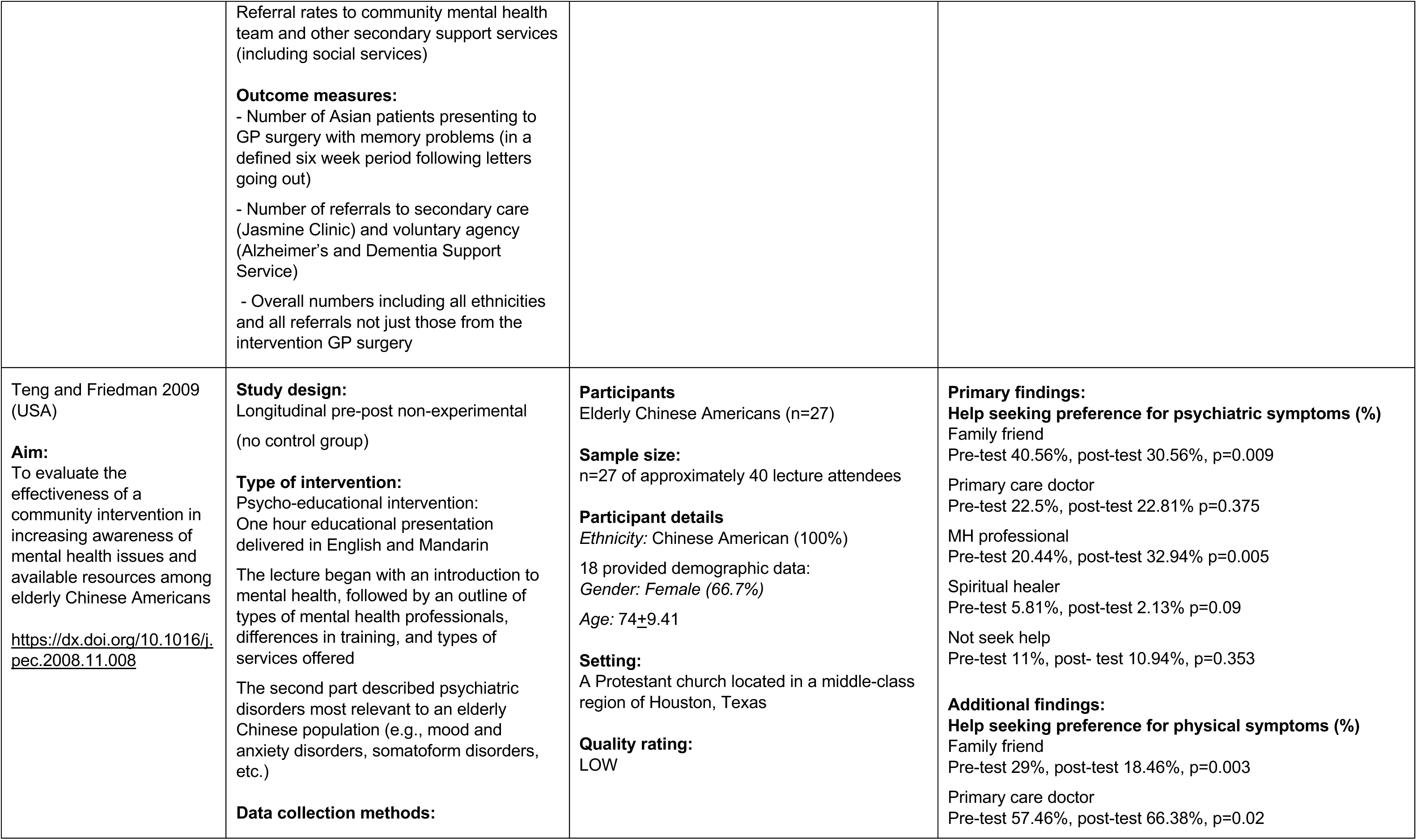
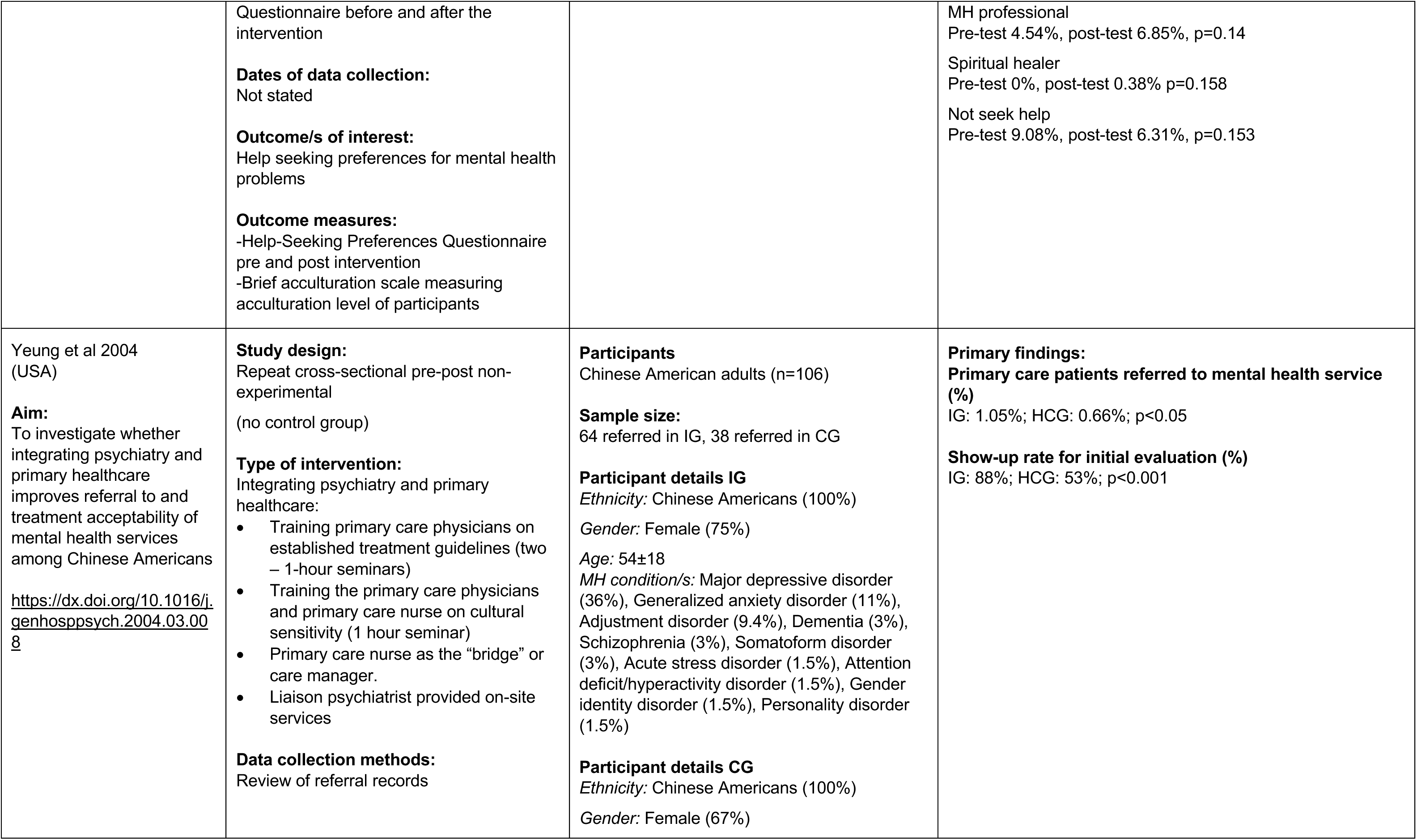
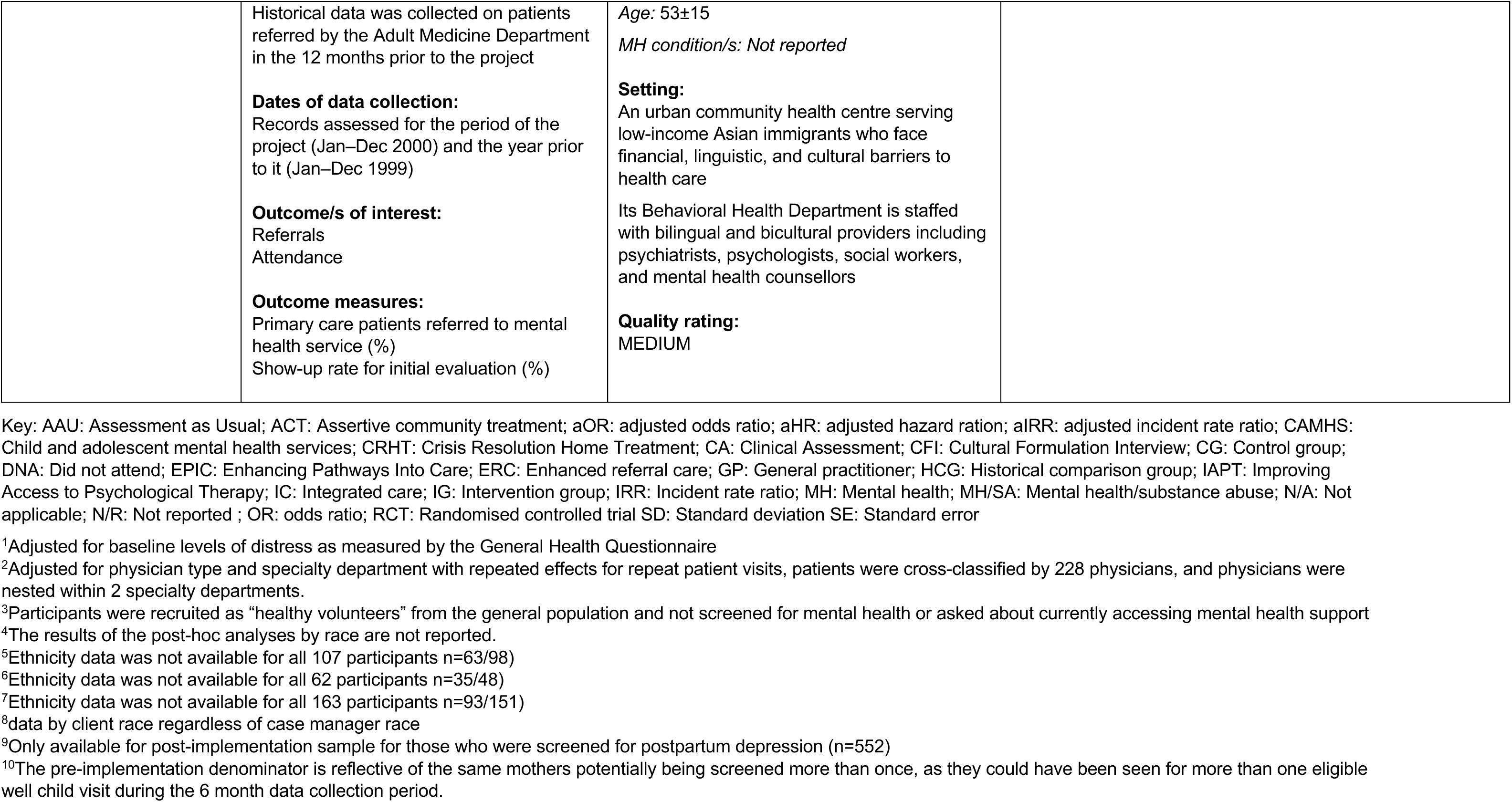
Characteristics of included studies.

Interventions that focused on instigating change at the *level of the individual* include:

- Implementing psycho-educational programs to promote psychotherapy entry and attendance among older African Americans (Alvidrez et al 2005).
- Psycho-educational videos to increase help seeking intentions around addressing difficulties coping with depressive symptoms for adolescent from ethnic minority groups (Martin et al. 2022).
- Patient education DVD and bilingual leaflet about dementia and getting help for memory symptoms for South Asian adults without known dementia aged >50 years (Mukadam 2017; Mukadam et al. 2018).
- Psycho-educational intervention to increase awareness of mental health issues and available resources among elderly Chinese Americans (Teng and Friedman 2009).

Interventions that focused on instigating change at the *organisational level* include:

- Integrating mental health and substance abuse services into primary care settings for older adults (> 65 years) from ethnic minority groups (Arean et al. 2008, Ayalon et al. 2007).
- Placing mental health link workers in GP surgeries to increase referrals of Black and Minority Ethnic individuals to the IAPT (Improving Access to Psychological Therapies) programme (Evans et al. 2014).
- Routine depression screening by medical assistants across ethnic minority groups (Gorman et al. 2021).

Interventions that focused on instigating change across multiple levels include:

#### Interpersonal and organisational levels Interventions

- Client-clinician ethnic and language matching for White and Hispanic homeless clients with severe mental illness in ongoing community treatment (Ortega and Rosenheck 2002).
- Psychiatry and primary healthcare service integration for Chinese American adults (Yeung et al. 2004).

#### Community and organisational levels Interventions

- Enhancing Pathways into Care project: information provision, community engagement; link worker and more appropriate and responsive services for those from the Pakistani community in mental health crisis (Hackett et al. 2009).
- Providing postpartum depression screening and referral to behavioural services by community health workers for Latinx immigrant mothers (Robidoux et al. 2023).

#### Individual and organisational levels Interventions

- Patient education and health care professional training to facilitate access to dementia screening and diagnosis for older Asians (Seabrooke and Milne 2009)

#### Individual, interpersonal and organisation levels Interventions

- Pre-intake intervention designed to enhance initial engagement at Child and Adolescent Mental Health Services (CAMHS) targeted at primary parental caregivers of all children and adolescents from black and minority ethnic groups referred and accepted into CAMHS (Michelson and Day 2014).
- Conducting person-centred cultural assessments with primary caregivers of children aged 2-7 from ethnic minority groups experiencing behaviour problems (Sanchez et al. 2022).

Seven of the interventions were conducted within primary care settings (Arean et al. 2008, Evans et al. 2014; Gorman et al. 2021, Mukadam 2017, Seabrooke and Milne 2009, Yeung et al 2004, Robidoux et al 2023); five within secondary care, specifically in community based or outpatient mental health services (Alvidrez et al 2005, Ortega and Rosenheck 2002); in Child and Adolescent Mental Health Services (Michelson and Day 2014, Sanchez et al 2022) or crisis or home treatment teams (Hackett at al 2009). One study reported that the study author delivered the intervention which consisted of a one-hour educational presentation at a faith-based organisation, but no further detail was provided (Teng and Friedman 2009). In the study by Martin et al. (2022) the participants were recruited by researchers from a crowd sourcing platform.

The mode of delivery was face to face for 11 studies (Alvidrez et al 2005, Arean et al. 2008, Evans et al 2014, Gorman et al 2021, Hackett et al. 2009, Martin et al. 2022, Ortega and Rosenheck 2002, Robidoux et al. 2023, Sanchez et al. 2022, Teng and Friedman 2009, Yeung et al. 2004), via telephone for one study (Michelson and Day 2014 and two studies sent information sent out in the post (Mukadam et al. 2018 and Seabrooke and Milne 2009).

#### Participants

The number of participants in each study ranged from 23 (Evans et al. 2014) to 21,377 (Gorman et al. 2021). Ten studies focused on adults (Alvidrez et al. 2005, Arean et al. 2008; Evans et al. 2014; Gorman et al. 2021, Hackett et al. 2009; Mukadam et al. 2018, Ortega and Rosenheck 2002, Seabrooke and Milne 2009, Teng and Friedman 2009 ;Yeung et al. 2004), one on new mothers (Robidoux et al. 2023), two on caregivers of children, with children as the target population for accessing services (Michelson and Day 2014, Sanchez et al. 2022), and one on adolescents (Martin et al. 2022).

Four studies described the population group as being older adults (Alvidrez et al. 2005; Arean et al. 2008; Seabrooke and Milne 2009; Teng and Friedman 2009). The mean age of participants was reported by 10 studies though one only reported the age of the child being cared for (Michelson and Day 2014). In the nine other studies, the mean age ranged from 16.8 years old (Martin et al. 2022) to 74 years old (Teng and Friedman 2009, Alvidrez et al. 2005). Gender was reported across 11 studies and the percentage of participants who were female ranged from 9.4% (Ortega and Rosenheck 2002) to 100% (Robidoux et al. 2023).

#### Ethnic minority groups

Six studies included participants from specific ethnic minority groups - Pakistani (Hackett et al. 2009) South Asian (Mukadam 2017); Latina (Robidoux et al. 2023); Chinese American (Teng and Friedman 2009; Yeung et al. 2004); African American (Alvidrez et al. 2005); Hispanic (Ortega and Rosenheck 2002) The remaining eight studies included participants from a variety of ethnic minority groups (Arean et al. 2008; Evans et al. 2014; Gorman et al. 2021; Hackett et al. 2009; Martin et al. 2022; Michelson and Day 2014; Sanchez et al. 2022; Seabrooke and Milne 2009) and the details are presented as described by the study authors.

- Asian (4 studies): Arean et al. 2008; Gorman et al. 2021; Sanchez et al. 2022; Seabrooke and Milne 2009.
- Black and minority ethnic (2 studies): Evans et al. 2014; Michelson and Day 2014.
- Black/African American (5 studies): Arean et al. 2008; Gorman et al. 2021; Martin et al. 2022; Sanchez et al. 2022.
- American Indian (1 study): Sanchez et al. 2022.
- Latino or Hispanic (2 studies): Arean et al. 2008; Sanchez et al. 2022.
- Haitian (1 study): Sanchez et al. 2022.
- Bi/multiracial (1 study): Sanchez et al. 2022.

#### Mental health conditions

Six studies focused on participants with various mental health conditions as follows:

- Anxiety (3 studies): Arean et al. 2008; Michelson and Day 2014; Yeung et al. 2004.
- Depression (4 studies): Arean et al. 2008; Martin et al. 2022; Ortega and Rosenheck 2002; Yeung et al. 2004.
- Psychotic symptoms or disorders (2 studies): Michelson and Day 2014; Ortega and Rosenheck 2002.
- Psychiatric problems (1 study): Ortega and Rosenheck 2002.
- Mood disorders (2 studies): Alvidez et al. 2005; Michelson and Day 2014.
- Developmental disorders (1 study): Michelson and Day 2014.
- Emotional and behavioural disorders of childhood (1 study): Michelson and Day 2014.
- Other mental health issues (1 study): Yeung et al. 2004.

Three studies focused on specific mental health conditions which were behaviour problems (Sanchez et al. 2022), dementia (Mukadam 2017) and post-partum depression (Robidoux et al. 2023). Two further studies did not provide specific details of the mental health conditions of the participants who were described as those referred for Improving Access to Psychological Therapy (Evans et al. 2014) or those experiencing mental health crisis (Hackett at al. 2009).

A further four studies included healthy participants being screened for depression (Gorman et al. 2021), help seeking intentions around depression (Martin et al. 2018); raising awareness of mental health issues (Teng and Friedman 2009) and raising awareness of dementia and memory problems (Seabrooke and Milne 2009).

#### Study design

There were eight experimental studies (Alvidrez et al. 2005; Arean et al. 2008; Evans et al. 2014; Gorman et al 2021; Martin et al. 2022; Michelson and Day 2014; Mukadam 2017, Sanchez et al. 2022). Of these four were randomised controlled trials (Arean et al. 2008; Martin et al. 2022; Mukadam 2017; Sanchez et al. 2022) and four were quasi-experimental studies (Alvidrez et al. 2005, Evans et al. 2014; Gorman et al. 2021, Michelson and Day 2014). There were six non-experimental studies (Hackett et al. 2009; Ortega & Rosenheck 2002; Robidoux et al. 2023; Teng and Friedman 2009, Seabrooke and Milne 2009; Yeung et al. 2004). For further detail see Section 6.2.

#### County of origin

Nine studies originated from USA (Alvidrez et al. 2005, Arean et al. 2008, Gorman et al 2021 Sanchez et al. 2022, Martin et al. 2022, Teng and Friedman 2009, Yeung et al 2004, Ortega and Rosenheck 2002, Robidoux et al. 2023) and five from UK (Evans et al. 2014, Michelson and Day 2014. Mukadam 2017, Seabrooke and Milne 2009, and Hackett et al. 2009).

#### Outcomes

The 14 studies reported a total of 40 relevant outcomes. These were grouped in to seven categories: help seeking intentions (Martin et al, 2022; Mukadam et al. 2018; Teng and Friedman 2009), screening (Gorman et al. 2021; Robidoux et al. 2023), initial attendance (Alvidrez et al. 2005, Arean et al. 2008, Michelson and Day 2014, Sanchez et al. 2022, Seabrooke and Milne 2009, and Yeung et al. 2004), ongoing attendance (Alvidrez et al. 2005, Arean et al. 2008, Michelson and Day 2014, Sanchez et al. 2022.), referrals (Yeung et al. 2004, Seabrooke and Milne 2009, Evans et al. 2014, Hackett et al. 2009, Robidoux et al. 2023), service use (Hackett et al. 2009; Ortega and Rosenheck 2002; Sanchez et al. 2022) and clinical outcomes (Arean et al. 2008, Ortega and Rosenheck 2002, Sanchez et al. 2022).

Outcomes that were categorised as screening were:

- Rates of depression screening (Gorman et al 2021).
- Screening rate of eligible postpartum mothers (Robidoux et al. 2023).

Outcomes that were categorised as referrals were:

- Number of referrals to secondary care (Seabrooke and Milne 2009).
- Primary care patients referred to mental health service (Yeung et al 2004).
- Referral rate for patients that screened positive (Robidoux et al. 2023).
- Referral rates for IAPT (Evans et al. 2014).
- % referrals to crisis resolution home treatment or inpatient ward (Hackett et al. 2009).

Outcomes that were categorised as help seeking intentions were:

- Attitudes to help-seeking for memory problems (Mukadam et al. 2018).
- Help seeking preference for physical and psychiatric symptoms (Teng and Friedman 2009).
- Help-seeking intentions (Martin et al. 2022).
- Depression stigma^5^ (Martin et al. 2022).

Outcomes that were concerned with initial attendance were:

- Proportion of patients starting therapy (Alvidrez et al. 2005).
- Attendance at first appointment (did not attend, attended, cancelled) (Michelson and Day 2014).
- Initial treatment session and attendance (Sanchez et al. 2022).
- Show-up rate for initial evaluation (Yeung et al 2004).
- Estimated number of time (days) to the first visit of any type (Arean et al. 2008).
- % subjects having at least one mental health visit (Arean et al. 2008).
- Presenting to the GP with memory problems in older Asians (Seabrooke and Milne 2009).

Outcomes that were categorised as ongoing attendance were:

- Attendance within first three appointments (did not attend, attended, cancelled) (Michelson and Day 2014).
- Number of sessions attended over 3 months (Alvidrez et al. 2005).
- Weekly session attendance rates (Sanchez et al. 2022).
- Average number of visits (Mean) (Arean et al. 2008).

Outcomes that were categorised as service use were:

- Medical-surgical services use, outpatient psychiatric service use and substance abuse services use (Ortega and Rosenheck 2002).
- Percentage service use (home treatment, by British/Asian Pakistani; Hackett et al. 2009).
- Number and proportional percentages of admissions (Hackett et al. 2009).
- Average length of stay (Hackett et al. 2009).
- Total services used (Ortega and Rosenheck 2002).
- Completion of first treatment module (Sanchez et al. 2022).

Clinical outcomes reported in the studies were:

- Depression scores (Ortega and Rosenheck 2002).
- Psychiatric problems (Ortega and Rosenheck 2002).
- Psychosis symptoms (Ortega and Rosenheck 2002).
- Anxiety (Arean et al. 2008).
- Depression (Arean et al. 2008).
- Treatment response (child behaviour problems dropped into the subclinical range (Sanchez et al. 2022).

Additionally, five studies compared the results of the ethnic minority participants with White participants (Arean et al. 2008; Gorman et al.2021; Martin et al. 2022; Michelson and Day 2014; Ortega and Rosenheck 2002) across a range of outcomes.

#### Methodological quality

The quality of the included studies was assessed using the Academy of Nutrition and Dietetics Quality Criteria Checklist (QCC) (Academy of Nutrition and Dietetics 2022). The results are summarised in Table 7. Only two studies (Arean et al. 2008; Gorman et al. 2021) were rated as high quality and four studies (Hackett et al. 2009; Robidoux et al. 2023; Seabrooke & Milne 2009; Teng and Friedman 2009) were rated as low quality. The remaining eight studies (Alvidrez et al. 2005; Evans et al. 2014; Martin et al 2022; Michelson and Day 2014; Mukadam et al. 2018; Ortega and Rosenheck; Sanchez et al. 2022; Yeung et al 2004) were rated as medium quality.

**Table 7:** Summary results of the QCC critical appraisal.

| Study | QCC questions |  |  |  |  |  |  |  |  |  | Overall designation |
| --- | --- | --- | --- | --- | --- | --- | --- | --- | --- | --- | --- |
|  | 1 | 2 | 3 | 4 | 5 | 6 | 7 | 8 | 9 | 10 |  |
| Alvidrez et al. 2005 | Y | Y | N | Y | N | Y | Y | N | Y | Y | Ø |
| Arean et al. 2008 | Y | Y | Y | Y | N | Y | Y | Y | N | Y | + |
| Evans et al. 2014 | Y | N | N | Y | N | Y | N | N | Y | Y | Ø |
| Gorman et al 2021 | Y | Y | Y | Y | N | Y | Y | Y | Y | Y | + |
| Hackett et al. 2009 | N | Y | UC | NA | UC | UC | Y | N | N | Y | - |
| Martin et al. 2022 | Y | N | Y | Y | UC | Y | Y | N | Y | Y | Ø |
| Michelson and Day 2014 | Y | Y | N | Y | N | Y | N | N | Y | Y | Ø |
| Mukadam et al. 2018 | Y | Y | N | Y | N | UC | Y | Y | Y | Y | Ø |
| Ortega & Rosenheck 2002 | Y | UC | N | N | N | Y | Y | Y | Y | Y | Ø |
| Robidoux et al. 2023 | Y | N | UC | NA | UC | N | Y | N | N | Y | - |
| Sanchez et al. 2022 | Y | Y | Y | Y | N | Y | Y | Y | Y | Y | Ø |
| Seabrooke & Milne 2009 | N | N | UC | NA | N | N | N | N | N | UC | - |
| Teng and Friedman 2009 | Y | N | NA | Y | N | Y | N | Y | N | UC | - |
| Yeung et al 2004 | Y | Y | UC | NA | N | Y | Y | Y | Y | Y | Ø |

Y=Yes, N=No, UC=Unclear, NA=Not applicable, +=High, Ø = Medium, - = Low

**Table 8:** Results of the assessment of the strength of evidence elements for studies included in the rapid review.

| Strength of Evidence Elements of interventions focusing on change at the individual level |  |  |  |  |  |
| --- | --- | --- | --- | --- | --- |
| Study author | Quality* | Consistency | Quantity | Clinical Impact | Generalisability |
| Alvidrez<br>et al. 2005 | Quasi-experimental<br>study<br>QCC rating – Medium | - ethnicity of psycho educator on initial or ongoing appointment attendance<br>- Initial appointment attendance<br>↑ ongoing appointment attendance | Sample size<br>n=69 | No effect size data<br>(Mean±SD, p values) | USA<br>Older adults<br>African American participants<br>Psycho-educational intervention |
| Martin<br>et al. 2022 | RCT<br>QCC rating – Medium | - depression stigma (video 1 for Black participants)<br>↓ depression stigma (video 2 for Black participants; (video 1 and 2 for non-Black participants)<br>↑ help seeking intention doctor/GP (video 1 and 2 for both Black and non-Black participants)<br>↑ help seeking intention MH professionals (Video 1 for both Black and non-Black participants)<br>- help seeking intention MH professionals (Video 2 for both Black and non-Black participants)<br>- Black relative to non-Black on all outcomes | Sample size<br>n=1093 | No effect size data<br>(Mean±SD, p values) | USA<br>Adolescents aged 14-18 years<br>Black participants<br>Psycho-educational intervention |
| Mukadam<br>et al. 2018 | RCT<br>QCC rating – Medium | - help seeking intentions | Sample size<br>n=78 | No effect size data<br>(Mean±SD, p values) | UK<br>Adults >50 years |
|  |  |  |  |  | South Asian participants<br>Psycho-educational intervention |
| Teng and Friedman 2009 | Non-experimental study<br>QCC rating – Low | ↑ help seeking intention MH professional<br>- help seeking intention GP | Sample size: n=27 | No effect size data (Mean±SD, p values) | USA<br>Older adults<br>Chinese American participants<br>Psycho-educational intervention |
| **Strength of Evidence Elements of interventions focusing on change at the organisational level** |  |  |  |  |  |
| Arean et al. 2008 | RCT<br>QCC rating - High | ↓ number days until first visit (Blacks)<br>- number days until first visit (Whites, Latino, Asian)<br>- number days until first visit (Black, Latino, Asian relative to White)<br>↑ initial appointment attendance (Latinos)<br>↓ initial appointment attendance (Asians)<br>- initial appointment attendance (Whites, Blacks)<br>- initial appointment attendance (Blacks, Latino relative to White)<br>↓ initial appointment attendance (Asians relative to White)<br>↑ ongoing appointment attendance (Whites, Blacks, Latino)<br>↓ ongoing appointment attendance (Asians)<br>- depression scores (Black, Latino, Asian, White)<br>- anxiety scores (Black, Latino, Asian, White) | Sample size n=2022 | Effect sizes calculated | USA<br>Older adults (>65 years)<br>Ethnic minority groups<br>Integrated MH care in a primary care |
| Evans et al. 2014 | Quasi-experimental study<br>QCC rating - Medium | ↑ referrals |  | No effect size data<br>Descriptive statistics | UK<br>Adults<br>Black and Minority Ethnic<br>Link worker |
| Gorman et al 2021 | Quasi-experimental study<br>QCC rating - High | ↑ screening<br>↑ screening<br>(Black/African American and Asian participants to White)) | Sample size<br>45,157 visits<br>by 21,377<br>unique<br>patients | Effect sizes calculated | USA<br>Adults<br>Ethnic minority groups<br>Routine depression screening by medical assistants |
| **Strength of Evidence Elements of interventions focusing on change on change at both interpersonal and organisational levels** |  |  |  |  |  |
| Ortega and Rosenbeck 2002 | Non-experimental study<br>QCC rating -Medium | - service use<br>- clinical outcomes <sup>a</sup><br>↓ Hispanic clients with Hispanic case manager – psychosis symptoms | Sample size<br>n=2575<br>White<br>n=242<br>Hispanic | No effect size data<br>(Mean±SE, p values) | USA<br>Adults (Homeless clients)<br>Hispanic<br>Client-case manager ethnic matching |
| Yeung et al. 2004 | Non-experimental study<br>QCC rating - Medium | ↑ referral<br>↑ initial appointment attendance | Sample size<br>n=106 | No effect size data<br>Proportions, p values | USA<br>Adults<br>Chinese Americans<br>Integrating psychiatry and primary healthcare |
| **Strength of Evidence Elements of interventions focusing on change on change at both community and organisational levels** |  |  |  |  |  |
| Hackett et al. 2009 | Non-experimental study<br>QCC rating - Low | ↑ referrals<br>↑ service use<br>↓ length of stay | Sample size<br>3.9% of<br>Inpatient<br>admissions<br>(2005-2006)<br>2.9% of<br>Crisis | No effect size data<br>Descriptive statistics | UK<br>Adults<br>Pakistani participants<br>Community engagement<br>Link worker |
|  |  |  | Resolution total episodes (2005-2006)<br>3.6% of Crisis resolution home treatment (2005-2006) |  |  |
| Robidoux et al. 2023 | Non-experimental study<br>QCC rating -Low | ↑ screening<br>- referral | Sample size<br>n=1846 | No effect size data<br>Proportions, p values | USA<br>Mothers<br>Lantinx<br>Community health workers assisting with postpartum depression screening |
| **Strength of Evidence Elements of interventions focusing on change on change at both individual and organisational levels** |  |  |  |  |  |
| Seabrooke and Milne 2009 | Non-experimental study<br>QCC rating – Low | ↑ initial appointment attendance<br>↑ referral | Sample size | No effect size data<br>Descriptive statistics | UK<br>Older Adults<br>Asians<br>Educational leaflets about dementia<br>HCP education |
| **Strength of Evidence Elements of interventions focusing on change on change at both individual and organisational levels** |  |  |  |  |  |
| Michelson and Day 2014 | Quasi-experimental study<br>QCC rating - Medium | - initial appointment attendance<br>- ongoing appointment attendance<br>↓ Initial and ongoing appointment DNA rates | Sample size<br>n=332 | No effect size data<br>Proportions, p values | UK<br>Primary parental caregivers of all children and adolescents<br>Black and Minority Ethnic<br>Pre-appointment engagement intervention |
|  |  | - initial appointment DNA rates<br>(Black and Minority Ethnic compared to White) |  |  |  |
| Sanchez et al.<br>2022 | RCT<br>QCC rating - Medium | - ongoing treatment attendance<br>(overall results)<br>- initial treatment attendance<br>(overall results)<br>↑ treatment completion<br>(overall results)<br>- treatment response<br>(overall results)<br>↑ ongoing treatment attendance<br>(In Spanish)<br>↑ initial treatment attendance<br>(In Spanish)<br>↑ treatment completion<br>(In Spanish)<br>↑ treatment response)<br>(In Spanish) | Sample size<br>n=89 | Effect sizes calculated | USA<br>Primary caregivers of children aged 2-7 years<br>Ethnic minority groups<br>(Hispanic/Latinx, Haitian, or Not Hispanic or Haitian)<br>Cultural formulation interview to assess perceptions of mental health and experiences of treatment<br>Behavioural problems |

<sup>a</sup> The interaction of client and case manager race was only significant for psychosis symptoms where Hispanic clients of Hispanic case managers had less reduction in psychotic symptoms than other ethnic and racial pairings (p=0.001).
Key: **Green** – Intervention improved outcomes; **Amber** – No difference between intervention and comparison; **Red** – Outcomes favour the comparison, or ; No colour – only descriptive statistics, no statistical significance can be identified
\* The results of the QCC assessment for each study is presented in Table 7

### 2.2 Effectiveness of interventions focusing on change at the individual level

**Psycho-educational intervention to promote psychotherapy entry and attendance for African Americans** (Alvidrez et al. 2005; USA)

#### Initial attendance

- There were no significant differences in the proportion of older African Americans patients, who received the psycho-educational intervention, starting therapy compared those who had been referred for psychotherapy in the 12 months prior to the intervention (p>0.05).
- The ethnicity of the psycho-educators (whether they were African American or non-African American), did not have a significant effect (p=0.8) on the proportion of older African American patients starting psychotherapy following the psycho-educational intervention (p>0.05).

#### Ongoing attendance

- Those who began psychotherapy following the psycho-educational intervention were significantly more likely to attend more psychotherapy sessions in a 3-month period than those who had been referred for psychotherapy in the 12 months prior to the intervention (p<0.05).
- The ethnicity of the psycho-educators (whether they were African Americans or non-African Americans) did not significantly influence the number of sessions attended by older African Americans following the psycho-educational intervention (p>0.05).

Psycho-educational intervention to increase help seeking intentions around addressing difficulties coping with depressive symptoms for adolescents from black and non-Black ethnic minority groups **(Martin et al. 2022: USA)**

#### Depression stigma (surrogate outcome for help seeking intentions)

- There were no significant differences in depression stigma for Black participants who viewed a video that addressed difficulties coping with depressive symptoms (video 1) compared to baseline (p>0.001^6^).
- There was a significant reduction in depression stigma for Black participants who viewed a video tailored to address unique aspects of being a Black adolescent girl (video 2) compared to baseline (p<0.001^6^).
- For non-Black participants, those who watched video 1 and video 2 experienced a significant decrease in mean depression stigma scores (p<0.001^6^).
- There were no significant differences in depression stigma for Black participants relative to non-Black participants across video 1, 2 and the control video (hobbies and interests) (p>0.001^6^).

#### Help seeking intentions

- There were significant increases in intention to seek help for emotional problems and suicidal thoughts from a doctor/GP for both Black and non-Black participants who viewed a video that addressed difficulties coping with depressive symptoms, (video 1) compared to baseline (p<0.001^6^).
- There were significant increases in intention to seek help for emotional problems and suicidal thoughts from a mental health professional for both Black and non-Black participants who viewed a video that addressed difficulties coping with depressive symptoms, thoughts that life is not worth living, false assumptions about treatment, and decisions on how and when to seek help (video 1) compared to baseline (p<0.01^7^).
- There were significant increases in intention to seek help for emotional problems and suicidal thoughts from a doctor/GP for both Black and non-Black participants who viewed a video tailored to address unique aspects of being a Black adolescent girl (video 2) compared to baseline (p<0.01).
- There were no significant differences in intention to seek help for emotional problems and suicidal thoughts from a mental health professional for both Black and non-Black participants who viewed a video tailored to address unique aspects of being a Black adolescent girl (video 2) compared to baseline (p>0.01).
- While results of the post-hoc analysis by ethnicity were not reported, the authors stated that the analysis did not show difference across ethnicity in a preferred source for help.

Patient education DVD and bilingual leaflet about dementia and getting help for memory symptoms for South Asian adults without known dementia aged >50 years **(Mukadam et al. 2018: UK)**

#### Help seeking intentions

- There were no significant differences in help seeking intentions for memory problems for South Asian adults without known dementia aged >50 years who received a trilingual DVD and information leaflet (n=17) compared to those who did not (p>0.05).
- Post hoc analysis showed that those who viewed the DVD^8^, were significantly more likely to seek help for memory problems compared to those in the control group (not receiving a DVD) (p<0.05), however these results were based on small numbers and should be treated with caution.

Psycho-educational intervention to increase awareness of mental health issues and available resources among elderly Chinese Americans **(Teng and Friedman 2009: USA)**

#### Help seeking intentions

- Immediately following a psycho-educational intervention (one-hour didactic lecture to raise mental health awareness) for older Chinese American participants, a significant increase in intention to seek help from mental health professional for psychiatric symptoms was detected (p=0.005). However, there were no significant differences in intention to consult primary care doctors (p=0.05)^9^.

### 2.3 Bottom line results of interventions focusing on change at the individual level

There were four studies (two RCTs, one quasi-experimental study and one non-experimental study) that investigated the effectiveness of four different psycho-educational interventions focusing on change at the individual level, and targeted at different ethnic minority groups and age ranges. Three were conducted in the USA and one in the UK.

- A quasi-experimental study (<u>medium quality rating</u>) found that a **psycho-educational intervention** aimed at promoting **psychotherapy entry and attendance** among **African Americans, increased ongoing attendance** but had **no effect on initial attendance** compared to those referred for psychotherapy in the 12 months previous. Additionally, the ethnicity of psycho-educators did not impact initiation or attendance (Alvidrez et al. 2005; USA)
- A RCT (<u>medium quality rating</u>) found that a **psycho-educational intervention** that addressed difficulties coping with depressive symptoms led to **increased help-seeking intentions in both Black and non-Black participants,** although **did not reduce depression stigma for Black participants.** However, intervention effects on depression stigma and help-seeking were comparable regardless of ethnicity (Martin et al. 2022: USA)
- The same RCT (<u>medium quality rating</u>) found that a **psycho-educational intervention** addressing unique aspects of being a Black adolescent girl with **depressive symptoms** led to **reduced depression stigma** and **increased help-seeking intentions in Black and non-Black participants,** with comparable effects observed regardless of ethnicity (USA) (Martin et al. 2022: USA)
- A RCT (<u>medium quality rating</u>) found that a trilingual DVD and information leaflet aimed at educating **South Asian adults aged over 50 years about dementia** and memory symptoms **did not impact help-seeking intentions** (Mukadam et al. 2018: UK).
- A non-experimental study (<u>low quality rating</u>) showed that a psycho-educational intervention to increase **awareness of mental health issues** and available resources among **elderly Chinese Americans increased their intention to seek help from mental health professionals**, but not from primary care doctors (Teng and Friedman

### 2.4 Effectiveness of interventions focusing on change at the organisational level

**Integrating mental health and substance abuse services into primary care settings for older adults (> 65 years) from ethnic minority groups** (Arean et al. 2008; Ayalon et al, 2007; Levkoff et al. 2004: USA).

#### Initial attendance

- **Asian participants** were significantly less likely **to have at least one mental health visit** when mental health and substance abuse services were integrated into primary care compared to speciality mental health care (aOR^10^ 0.53 (95% CI: 0.37, 0.77).
- However, **Latino participants** were significantly more likely to have **at least one mental health visit** when mental health and substance abuse services were integrated into primary care compared to speciality mental health care (aOR^7^ 3.6 (95% CI: 2.22, 5.82).
- Subgroup analysis on data from one of 10 GP practices revealed **Black participants** were significantly more likely to have **at least one mental health visit** when mental health and substance abuse services were integrated into primary care compared to speciality mental health care (aOR^7^ 14.13 (95% Cl: 4.76, 41.95).
- The odds of having **at least one mental health visit** when mental health and substance abuse services were integrated into primary care was not significantly different **for Black, Latino or Other ethnicities** relative to White participants. However, the odds of **Asian participants** having **at least one mental health visit** when mental health and substance abuse services into primary care was significantly lower relative to White participants (aOR^7^ 0.24; 95% CI: 0.09, 0.61)^11^.
- The estimated **number of days from baseline evaluation to engagement in the first mental health visi**t was significantly lower for **Black participants** (p<0.05) when mental health and substance abuse services were integrated into primary care compared to speciality mental health care.
- There were no significant differences **in the number of days from baseline evaluation to engagement in the first mental health visit across** the **different ethnic minority groups** compared to White participants (p>0.05).
- Subgroup analysis on data from one of 10 GP practices revealed that the time from baseline evaluation to engagement in the first mental health visit was significantly lower when mental health and substance abuse services were integrated into primary care for both **Black** (aHR^12^ 7.82 (95% CI 3.65, 16.75) and **White participants** (aHR^8^ 2.48 (95% CI: 1.20, 5.13) relative to speciality mental health care.

#### Ongoing attendance

- **White, Black and Latino participants** had a significantly **greater number of visits** when mental health and substance abuse services integrated into primary care compared to speciality mental health (p<0.05).
- **Asian participants** had a significantly **greater number of visits in specialty mental health services** compared to when mental health and substance abuse services were integrated into primary care (p<0.05).^13^
- Subgroup analysis on data from one of the 10 GP practices revealed that when services were offered in a speciality mental health setting, **Black participants** had a significantly smaller number of overall mental health visits relative to White participants (aIRR^8^ 2.87 (95% CI 1.06, 7.73).

#### Clinical outcomes

- There were no significant differences in levels of anxiety or depression at six months for White and ethnic minority participants when mental health and substance abuse services were integrated into primary care or when mental health services were offered in speciality mental health care.
- Across both groups mean post treatment symptoms of anxiety and depression, remained relatively high.

Placing mental health link workers in GP surgeries to increase referrals of Black and Minority Ethnic individuals to the IAPT programme **(Evans et al. 2014:UK)^14^**

#### Referrals

- Referral rates of **Black and Minority Ethnic patients** from primary care to IAPT from **practices who employed a link worker** increased from January 2010 to July 2020 (0.65 to 1.37 patients per 10,000 practice population).
- This referral rate remained stable until the end of the study despite the addition of more link workers.
- The mean referral rate of Black and Minority Ethnic patients (standardised to the size of the population served by practices) to IAPT for practices without a link worker was 0.35 Black and Minority Ethnic referrals per week per 10,000 patients and this was unchanged throughout the period of the study.

Routine depression screening by medical assistants across ethnic minority groups (Gorman et al 2021: USA).

#### Screening

- Depression screening rates significantly increased for Black/African, Asian and White and other ethnicities when routine depression screening was performed by medical assistants compared to routine depression screening performed by physicians in the 12 months prior to the intervention (p<0.001).
- When **routine depression screening was performed by physicians,** screening was significantly less likely to occur at visits by **Asian patients** than at visits by White patients (OR 0.82, 95% CI 0.76, 0.89) and at visits by **Black/African American**

**patients** than at visits by White patients (OR 0.91, 95% CI 0.84, 0.99).

- When **routine depression screening was performed by medical assistants,** screening was significantly more likely to occur at visits by **Asian patients** than at visits by White patients (OR 1.28, 95% CI 1.09, 1.50) and at visits by **Black/African American** patients than at visits by White patients (OR 1.11, 95% CI 1.02, 1.20)^15^.

### 2.5 Bottom line results of interventions focusing on change at the organisational level

There were three studies (one RCT and two quasi-experimental studies) that investigated the effectiveness of three different interventions focusing on change at the organisational level. Two were conducted in the USA and one in the UK.

- A RCT (<u>high quality rating</u>) that sought to **integrate specialist mental health services into primary care** for **older adults from ethnic minority groups** found that the number of days until first visit, initial and ongoing attendance rates varied depending on the ethnicity of the participants. Additionally, there was **no impact on levels of depression and anxiety** (Arean et al. 2008; Ayalon et al, 2007; Levkoff et al. 2004: USA).
- A quasi-experimental study (<u>medium quality rating</u>) found that placing **mental health link workers in GP surgeries** appeared to **increase referrals of Black and Minority Ethnic individuals to an IAPT program** compared to practices without link workers (Evans et al. 2014:UK UK).
- A quasi-experimental study (<u>high quality rating</u>) reported **improved depression screening rates for Black/African American, Asian,** White and other ethnic participants when **routine depression screening was undertaken by medical assistants** compared to physicians. Additionally, depression screening by medical assistants for Black/African American and Asian participants was more likely to occur compared to White participants (Gorman et al 2021: USA).

### 2.6 Effectiveness of interventions focusing on change across multiple levels

#### 2.6.1 Interpersonal and organisational levels

**Client-clinician ethnic and language matching for White and Hispanic homeless clients with severe mental illness in ongoing community treatment** (Ortega and Rosenheck 2002: USA).

##### Service use

- There were no significant differences for any of the service use outcomes (outpatient psychiatric services, medical-surgical services, substance abuse services, service integration - the receipt of a range of service by an individual client) for **Hispanic clients** (p>0.05).
- **Hispanic clients** had a significant increase in service integration (the total receipt of a range of services by an individual client) compared to **White clients** (p<0.05).^16^
- There were no significant interactions between **client and case manager race** in any of the service use outcomes (p>0.05).
- **Hispanic clients** had more problems at baseline in psychiatric and substance abuse domains and showed less improvement on several measures of psychiatric status over the next 12 months than White clients.

##### Clinical outcomes

- **White clients** had significantly better improvement on a number of clinical outcomes than **Hispanic clients**, namely psychosis symptoms (p=0.0001), psychiatric problems (p=0.04) and depression scores (p<0.05).
- The interaction of **client and case manager race** was only significant for psychosis symptoms where **Hispanic clients of Hispanic case managers** had less reduction in psychotic symptoms than other ethnic and racial pairings (p<0.05)^17^.

Psychiatry and primary healthcare service integration for Chinese American adults (Yeung et al. 2004: USA).

##### Referrals

- Following the **integration of psychiatry and primary care services**, there was a significant increase in the number of **Chinese American patients** referred to see a liaison psychiatrist within primary care compared to the preceding 12 months prior to implementation (p<0.05).

##### Initial attendance

- Following the **integration of psychiatry and primary care services**, there was a significant increase in the number of **Chinese American** patients who showed up for their initial mental health evaluation, compared to the preceding 12 months prior to implementation (p<0.05).

#### 2.6.2 Community and organisational level interventions

**Enhancing Pathways into Care project: information provision, community engagement; link worker and more appropriate and responsive services for those from the Pakistani community in mental health crisis** (Hackett et al. 2009: UK)^18^

##### Referrals

- There was an increase in the percentage of **Pakistani patients** receiving crisis referrals to a crisis resolution and home treatment service following the intervention (3.2% compared to 2.6%). No significance testing was reported.
- There was an increase in the number of referrals to the Pakistani Muslim Centre from home treatment (one in 2005 compared to seven in 2006) and from the inpatient ward (from zero in 2005 compared to nine in 2006).

##### Service use

- There was an increase in the proportion of **Pakistani patients** receiving home treatment (3.4% in 2005 compared to 3.8% in 2006) and admitted to hospital (3.8% in 2005 compared to 4% in 2006).
- There were minimal fluctuations in numbers and proportional percentages of **Pakistani patients** admitted to inpatient setting.
- The authors reported that there was an overall trend for reduction in inpatients length of stay across the five-year period.

Providing postpartum depression screening and referral to behavioural services by community health workers for Latinx immigrant mothers **(Robidoux et al. 2023: USA).**

##### Screening

- During the project, out of 832 well-child visits, 552 (66%) patients were screened for post-partum depression, compared to 452 (45%) prior to the implementation of the project. The difference in screening was significant (p < 0.001).

##### Referrals

- There were no significant differences in referral rates to behavioural health services for Latinx immigrant mothers after positive postpartum screening during routine well-child visits compared to the 6 months prior to implementation of the project (p<0.05)^19^.

#### 2.6.3 Individual and organisational level interventions

**Patient education and health care professional training to facilitate access to dementia screening and diagnosis in for older Asians** (Seabrooke and Milne 2009: UK)^20^

##### Initial attendance

- During the 6-week period following the **patient education element** of the interventions **(letters and information leaflets)**, five **Asian patients** presented at the GP surgery, compared to no patients in the three months prior to project implementation.

##### Referrals

- There was an increase in referrals to a specialist memory clinic and to a local voluntary agency.

#### 2.6.4 Individual, interpersonal and organisation level interventions

**Pre-intake intervention designed to enhance initial engagement at CAMHS targeted at primary parental caregivers of all children and adolescents from Black and Minority Ethnic groups referred and accepted into CAMHS** (Michelson and Day 2014: UK).

##### Initial and ongoing attendance

- There were no significant differences in **first appointment attendance rates** and **attending at least one appointment of the first three scheduled CAMHS appointments** between **children and adolescents from Black and Minority Ethnic groups** who received a **pre-intake engagement intervention** and those in the historical comparison group who received standard clinic procedures (p>0.05).
- The classification of reasons for not attending were (cancelled, discharged and did not attend). There were no significant differences for the categories of cancelled and discharged between **children and adolescents from Black and Minority Ethnic groups** in the **pre-intake engagement intervention** and the historical comparison group (p>0.05). However, significantly fewer **children and adolescents from Black and Minority Ethnic groups** were recorded as “did not attend” in the **pre-intake engagement intervention** group compared to those in the historical comparison group (p<0.05).

Conducting person-centred cultural assessments with primary caregivers of children aged 2-7 from ethnic minority groups experiencing behaviour problems **(Sanchez et al. 2022: USA).**

##### Initial attendance

- Cultural formulation interview with assessment as usual (CFI+AAU) was not found to be significantly more effective in improving initial treatment attendance of ethnically diverse children^21^ with behaviour problems and their primary caregivers compared to assessment as usual only (AAU) (p=0.40). However, when moderating for language provision, for those who received the intervention (CFI+AAU) in Spanish, the probability of attending the first treatment session was significantly higher (p<0.05).

##### Ongoing attendance

- Weekly session attendance of ethnically diverse children^15^ with behaviour problems and their primary caregivers was not significantly different between CFI+AAU and AAU (p=0.38). However, mean session attendance was significantly higher for CFI+AAU when services were provided in Spanish (p<0.05).

##### Treatment module completion

- Cultural formulation interview with assessment as usual was significantly more effective in improving first treatment module completion of ethnically diverse children^22^ with behaviour problems and their primary caregivers compared to AAU (p=0.03). Additionally, when moderating for language provision, for those who received CFI+AAU in Spanish, probability of first module completion was significantly higher (p<0.05), while no significant difference was observed between CFI+AAU and AAU when these were provided in English (p>0.05).

##### Clinical outcomes

- Treatment response (child behaviour problems dropped into the subclinical range) was not significantly different between groups receiving CFI+AAU or AAU (p>0.05). However, when services were provided in Spanish, children and their primary caregivers in the CFI+AAU group were significantly more likely to be treatment responders compared to AAU (p<0.05).

### 2.7 Bottom line results for interventions focusing on change across multiple levels

There were seven studies (one quasi-experimental study and six non-experimental studies) that investigated the effectiveness of seven different interventions focusing on change at across multiple levels. Four were conducted in the USA and three in the UK.

- A non-experimental study (<u>medium quality rating</u>) found that an intervention that involved **client-clinician ethnic and language matching** showed **no difference in service use and clinical outcomes** for homeless clients across **different ethnic minority groups with severe mental illness**. However, **Hispanic clients of Hispanic case managers** had **less reduction in psychotic symptoms** than other ethnic and racial pairings (Ortega and Rosenheck 2002: USA).
- A non-experimental study (<u>medium quality rating</u>) of an intervention **integrating psychiatry and primary healthcare services** showed **increased referrals to liaison psychiatrists within primary care** and i**mproved initial attendance** rates for mental health evaluations among **Chinese American adults** (Yeung et al. 2004: USA).
- A non-experimental study (<u>low quality rating</u>) that investigated the **Enhancing Pathways Into Care project** suggested improved crisis referrals to mental health services and increased utilization of crisis resolution and home treatment services among **Pakistani patients**, alongside improvement in referrals to community centres, home treatment and inpatient services and decreased inpatient length of stay (Hackett et al. 2009: UK).
- A non-experimental study (<u>low quality rating</u>) of **community health workers providing postpartum depression screening** during well-child visits was found to **improve screening rates for Latinx immigrant mothers** compared to pre-project rates but did **not have a significant impact on referral rates** to behavioural health services following positive screenings (Robidoux et al. 2023: USA).
- A non-experimental study (<u>low quality rating</u>) that involved **patient education and healthcare professional training** appeared to **increase initial attendance at GP surgeries by older Asians for dementia screening and diagnosis**, with subsequent **rises in referrals to specialist memory clinics** and local voluntary agencies (Seabrooke and Milne 2009: UK).
- A quasi experimental study (<u>medium quality rating</u>) of a **pre-intake engagement intervention** targeting **primary not show any significant differences in initial or ongoing appointment attendance rates** compared to a historical comparison group Michelson and Day 2014: UK).
- The classification of reasons for not attending were (cancelled, discharged and did not attend). There were no significant differences for the categories of cancelled and discharged between **children and adolescents from Black and Minority Ethnic groups** in the **pre-intake engagement intervention** and the historical comparison group (p>0.05). However, significantly fewer **children and adolescents from Black and Minority Ethnic groups** were recorded as “did not attend” in the **pre-intake engagement intervention** group compared to those in the historical comparison group (p<0.05).
- A RCT (<u>medium quality rating</u>) that investigated conducting **person-centred cultural assessments** alongside treatment as usual did **not impact initial or ongoing attendance or clinical outcomes** for **ethnically diverse children with behaviour problems** and their caregivers, yet when services were provided **in Spanish, attendance and treatment response (child behaviour problems dropped into the subclinical range) improved** (Sanchez et al. 2022: USA).

## 3. DISCUSSION

The purpose of this rapid review was to assess the effectiveness of interventions that enhance equitable or overall access to mental health services by ethnic minority groups. Studies included a diverse range of participants, encompassing various age groups and ethnic backgrounds with a range of mental health conditions. The most frequently cited mental health conditions were anxiety and depression.

There is a plethora of evidence over five decades indicating that inequalities in access to, experience, and outcomes of mental healthcare among ethnic minority groups (Bansal et al. 2022). Individuals from these groups are more likely to experience undiagnosed and untreated mental illness, enter healthcare services during crises or through other challenging pathways, and receive diagnoses of severe mental illnesses such as schizophrenia, bipolar and psychosis compared to individuals from White backgrounds (Halvorsrud et al. 2019). Delayed provision of mental health care is associated with poorer outcomes for both common and severe mental disorders, as well as increased reliance on crisis interventions (Boonstra et al. 2012). Additionally, international research has identified that the barriers to accessing mental health care for individuals from ethnic minority groups are associated with their ability to seek services and the availability of services (Lowther-Payne et al. 2023). One of the ways that these barriers can be addressed is through the integration of specialist mental health services within the primary care setting and two of the studies. This review found that the integration of specialist mental health services into primary care improved referrals and initial attendance rates among Chinese American adults (Yeung et al. 2004), whilst Arean et al. (2008) investigated the integration of mental health and substance abuse services into primary care targeting older adults from ethnic minority groups and reported varied attendance rates by ethnicity and no notable effect on depression and anxiety levels.

Insufficient knowledge about mental health and available services acts as a significant barrier, impeding individuals from seeking and accessing mental health care often leading to delays or avoidance of seeking help (Aggarwal et al. 2016, Interian et al. 2013) and can hinder decisions around treatment (Aggarwal et al. 2016). Across the included studies in this review several of the interventions have sought to address this by providing culturally appropriate information. For example, psycho-educational interventions have utilised resources such as leaflets (Mukadam et al. 2018; Seabrooke & Milne 2009), videos (Martin et al. 2022) and a lecture style presentation (Teng and Friedman 2009). The effectiveness of interventions showed mixed result for help seeking behaviour which was influenced by ethnicity and only one study (Seabrooke & Milne 2009), investigated the impact of psycho-educational interventions on initial attendance rates and ongoing referrals making it difficult to draw conclusions.

Other interventions sought to provide information verbally alongside various other elements in healthcare settings, ensuring that information about mental health and available services is effectively conveyed during consultations (Hackett et al. 2009, Michelson and Day, 2014, Robidoux et al. 2023; Sanchez et al. 2002). The effectiveness of the interventions varied, with some showing positive outcomes such as increased help seeking intentions, improved initial and ongoing attendance rates when verbal information was provided, while others did not show significant differences in outcomes compared to control groups.

Cultural stigma surrounding mental health issues may deter individuals from ethnic minority groups from seeking help. Therefore, addressing stigma is crucial in improving access to mental health care (Aggarwal et al. 2016; Giebel et al. 2015). A number of interventions in this review attempted to address stigma but this was not the main focus of the studies. For example, by normalising discussions about mental health (Arean et al. 2008), increasing understanding surrounding mental health issues (Teng and Friedman 2009) addressing misconceptions (Teng and Friedman 2009), encouraging individuals to feel more comfortable to seek help and engage in therapy without fear of judgment (Alvidrez et al. 2005; Hackett et al. 2009, Martin et al. 2022). Out of these studies, only one assessed stigma as an outcome, revealing that a psycho-educational intervention aimed at reducing depression stigma through a social-contrast based video was effective in enhancing help-seeking intentions and diminishing depression stigma in both Black and non-Black participants (Martin et al. 2022).

Mental health care providers may not be proficient in the languages spoken by ethnic minority groups in their community. This lack of language proficiency can hinder effective communication between the patient and the provider, making it difficult for the patient to express their thoughts, feelings, and concerns accurately (Aggarwal et al. 2016; Arundell et al. 2020; Giebel et al. 2015; Lock et al. 2023). This review identified six studies (Alvidrez et al. 2005, Teng and Friedman 2009, Mukadam et al. 2018, Robidoux et al. 2023; Ortega and Rosenbeck 2002; Sanchez et al. 2002) that incorporated language support into mental health interventions to improve accessibility and effectiveness for individuals from a diverse range of ethnic minority groups. The effectiveness of the interventions varied, with some showing positive outcomes such as increased intention to seek help or improved attendance and treatment response when provided in the patient’s preferred language, while others did not show significant differences in outcomes compared to control groups. However, incorporating language support into mental health care settings is a vital step toward addressing communication barriers and promoting equitable access to mental health services for individuals from ethnic minority groups. A recent report by the Equality and Social Justice Committee (Welsh Parliament 2024) recommends providing translators to patients to help avoid overlooking health conditions, while also emphasising the need to eliminate the use of family members as interpreters in medical settings. The wider evidence based has established that the use of professional interpreters can improve the quality and appropriateness of care (Public Health Wales and Swansea University 2023). The HEAR 2 study which explored the health experiences of asylum seekers and refugees in Wales found that patient satisfaction with interpretation services if generally high although sometimes there were issues in terms of dialect, gender, or cultural specificity (Public Health Wales and Swansea University 2023).

Ethnic minority groups often have distinct cultural norms, beliefs, and values surrounding mental health. These cultural differences can affect how individuals perceive and experience mental health issues and influence their willingness to seek help from mainstream mental health services. Developing culturally tailored mental health interventions that resonate with the cultural values and beliefs of ethnic minority groups have the potential to improve engagement and outcomes (Aggarwal et al. 2016; Lock et al. 2023). Six studies incorporated various approaches to enhance the cultural competency of mental health services. For example training for healthcare professionals to offer culturally sensitive care and onward referral to appropriate services (Seabrooke & Milne 2009; Yeung et al. 2004), developing culturally appropriate services like the Pakistani link worker (Hackett et al. 2009), tailoring interventions to address unique cultural aspects, such as racism’s impact on mental health for Black adolescent girls (Martin et al. 2022), utilising tools like the Cultural Formulation Interview (Sanchez et al. 2022), and customizing interventions to suit diverse cultural backgrounds (Michelson and Day 2014). These strategies aim to ensure that mental health services are sensitive, relevant, and effective across diverse cultural contexts. The effectiveness of these approaches varied across studies. While some interventions demonstrated positive outcomes, such as reduced depression stigma or improved attendance and treatment response when services were provided in Spanish, others did not show significant impacts on attendance or clinical outcomes. Therefore, the effectiveness of cultural competency interventions in mental health services depends on various factors, including the specific intervention, target population, and context of implementation. It is important to consider that some interventions may have shown effectiveness if they had better study designs, larger sample sizes and accounted for confounding factors.

### 3.1 Strengths and limitations of the available evidence

A wide range of interventions targeted at different ethnic minority groups and age ranges were described in the identified evidence. However, although we searched for evidence for Gypsy, Traveller, and Roma groups we were unable to find any research for this specific population group. Most of the interventions were conducted in real-life clinical settings. While in some cases this limited the generalisability of results due to the clinics or patient populations having unique features, it also bridged the gap between intervention development and implementation and demonstrate how they were employed in practice. Due to the high variation in the interventions and contexts in which they were implemented, evidence for the effectiveness of each intervention was limited.

The studies had many methodological limitations. Out of the 14 included studies, only four were RCTs, with another six having no control group. Control groups were often historical rather than concurrent, which could introduce additional confounding factors. As a result of the quality appraisal, only two studies were rated high (Arean et al. 2008, Gorman et al 2021), with eight medium (Alvidrez et al. 2005; Evans et al. 2014; Martin et al. 2022; Michelson and Day 2014; Mukadam et al. 2018; Ortega & Rosenheck 2002; Sanchez et al. 2022; Yeung et al. 2004), and four receiving a low quality rating (Hackett et al. 2009, Robidoux et al. 2023, Seabrooke & Milne 2009, Teng and Friedman 2009) due to methodological weaknesses. In many of the included studies, study groups were not comparable, which meant that any differences in the outcomes could be attributed to the differences at baseline. Most studies did not use blinding, while in the remaining ones it was not possible to establish if they did. However, due to the nature of the interventions, blinding those receiving or delivering the intervention was not always possible or appropriate. While blinding those collecting and analysing outcome data might be feasible, it was not reported or discussed in the included studies. Another common issue was problems with data analysis, particularly the lack of adjustment for potential confounders. In some cases, only descriptive statistics were available. The issues outlined here weaken the evidence for the effectiveness of the intervention and the generalisability of the included studies.

### 3.2 Strengths and limitations of this Rapid Review

This review employed systematic search methods, which included searching five complementary bibliographic databases using a comprehensive search strategy and conducting forward and backward tracking of citations in the included studies. Error and bias in study selection were mitigated by two reviewers independently screening the full texts of all the articles that passed the initial screening. These strategies helped to maximise the amount of identified relevant evidence. All such evidence was critically appraised and included regardless of its methodological quality to provide a full account of the state of the literature on the topic, but methodological limitations were taken into consideration when reporting the results. A single critical appraisal tool was used for all studies, which allowed the comparison of methodological quality across the different study designs while taking design into account when conducting the appraisal.

This work also has a number of limitations arising from the time constraints associated with a rapid review. Grey literature, such as pre-prints and research reported in non-academic sources (e.g., reports) was not searched, so the extent of potential publication bias is not known. In order for the review to be completed within the given timeframe, the scope had to be limited to studies of interventions aimed at ethnic minority groups, which meant that studies with other potentially relevant populations which may have a high percentage of ethnic minorities (e.g., refugees and asylum seekers) were excluded.

### 3.3 Implications for policy and practice

Findings from this review can help to inform the development of the draft Mental Health and Wellbeing Strategy (2024-2034) and draft Suicide and Self-harm Prevention Strategy (2024/2034) - which are currently out to consultation.

In summary, the findings support the following:

- Psycho-educational interventions that utilise culturally appropriate delivery methods, such as leaflets, videos, or lectures to encourage health seeking behaviour amongst ethnic minority groups.
- The integration of speciality mental health services within primary care setting.
- The implementation of language support such as the use of professional interpreters to improve the accessibility for individuals with limited English proficiency.
- The provision of training to emphasise cultural sensitivity and competence among healthcare providers to address disparities in mental healthcare access and outcomes.

### 3.4 Implications for future research

The majority of studies included in the review lacked methodological rigor, had small sample sizes, and lacked of control groups. Future research should prioritise rigorous study designs, including randomised controlled trials and longitudinal studies, to provide robust evidence on the effectiveness of interventions targeting ethnic minority populations in mental health care.

Additional research is required to compare outcomes between ethnic minority participants and White participants, aiming to evaluate the effectiveness of interventions in mitigating disparities in access to and outcomes of mental health care.

### 3.5 Economic considerations*

- Mental Health problems cost the Welsh economy £4.8 billion per annum. Seventy-two percent of these incurred costs are attributed to productivity losses of people living with mental health conditions and costs experienced by unpaid informal carers (Mcdaid 2022).
- Ethnic minority individuals are disproportionately affected by economic determinants of poor mental health including increased likelihood of low income (Money and Mental Health Policy Institute 2023, Bansal et al. 2022).
- Future research should investigate the economic benefit to both an NHS and societal perspective of improving access to mental health services for ethnic minority communities

*This section has been completed by the Centre for Health Economics & Medicines Evaluation (CHEME), Bangor University

## Data Availability

All data produced in the present study are available upon reasonable request to the authors

## 4. RAPID REVIEW METHODS

### 4.1 Eligibility criteria

Eligibility criteria for the review are presented in Table 4.

### 4.2 Literature search

#### 4.2.1 Evidence sources

Comprehensive literature searches were conducted across the following bibliographic databases without any publication date limits: PsycINFO via OVID, Medline via OVID, Cinahl via EBSCO, Web of Science, and Cochrane Central Register of Controlled Trials (CENTRAL).

#### 4.2.2 Search Strategy

Preliminary searches were undertaken during the development of the protocol in the Cochrane and NIHR Journals Library, Epistemonikos, JBI EBP database via Ovid, PubMed, Trip and PROSPERO databases using a combination of mental health terms (mental health OR psychological health OR mental illness OR bipolar OR schizophrenia OR psychosis OR depression OR anxiety OR post-traumatic stress disorder OT PTSD OR dementia OR alzheimer*), ethnicity terms (ethnic* OR minorit* OR racial*), intervention terms (intervention* OR improv*) and outcome terms (access* OR provision OR service* OR engag*). After that, targeted searches were conducted to identify literature focused on Gypsy, Traveller and Roma groups using specific search terms (gyps* OR roma OR romas OR romany OR romani OR romanis OR romanies OR traveler* OR traveller*).

Identified articles were then reviewed and analysed for the text words contained in the titles and abstracts. This informed the development of **a full search strategy designed on PsycINFO (on OVID) and adapted for other databases (see Appendix 1)**. The searches were conducted between 15^th^ and 19^th^ December 2023. Following database searches, forward and backward citation tracking were undertaken using citationchaser (Haddaway et al. 2022) and relevant studies were added to the review.

#### 4.2.3 Reference management

All citations retrieved from the database searches were imported into EndNote^TM^ (Thomson Reuters, CA, USA) and duplicates removed. At the end of this process, the citations that remained were exported as an TXT file and then imported to the software package Rayyan^TM^, where any remaining duplicates will be removed.

### 4.3 Study selection process

Two reviewers dual screened at least 20% of citations using the information provided in the titles and abstracts using Rayyan^TM^, resolving all conflicts where needed. The rest of the citations were screened by a single reviewer. For citations that appeared to meet the inclusion criteria, or in cases in which a definite decision could not be made based on the title and/or abstract alone, the full texts were retrieved. The full texts were then screened for inclusion by two reviewers and any disagreements resolved by a third reviewer. The flow of citations through each stage of the review process for each question are displayed in a PRISMA flowchart (Page et al. 2021).

### 4.4 Data extraction

All relevant data were extracted directly into tables by one reviewer and checked by another. The data extracted included specific details about the populations, including information about participants’ demographics and mental health status, study methods, interventions and outcomes of significance to the review questions and objectives. A data extraction template was developed and piloted for each of the included study designs. Minor amendments were made to the template as a result of the pilot.

### 4.5 Study design classification

We used the decision algorithm developed by Leatherdale (2019) to classify the research designs of the included studies

### 4.6 Quality appraisal

Eligible quantitative studies were assessed for risk of bias using the Academy of Nutrition and Dietetics **Quality Criteria Checklist (QCC)** (Academy of Nutrition and Dietetics 2022). Assessments were conducted by one reviewer and checked by a second, with any disagreements resolved by a third person. The checklist consists of 10 questions that address scientific soundness across a number of domains. The QCC tool can be applied to most quantitative study designs and is therefore suitable for reviews that include different study designs.

Based on responses to the 10 questions, a study is given an overall evaluation of positive, negative, or neutral. However, though for this review we adapted these to ‘High’, ‘Medium’ and ‘Low’ as per Duval et al. (2023).

- High: Strong quality - limited bias A study is given a high rating if the majority (6 or more) of the answers were ‘yes’ including criteria 1, 2, 3, and 4.
- Medium: Neither exceptionally strong nor exceptionally weak A study is given a medium if the answers to any of the first four validity questions (1- 4) is “No” but other criteria indicate strengths.
- Low: Weak quality - likely bias A study is given a low rating if the majority (six or more) of the answers across the 10 validity questions are “No”.

If any of the ten validity questions are marked ot applicable (N/A), the report requires a majority of “Yes” answers (including 1, 2, 3,4, as applicable) for a plus (+), or a majority or “No” answers for a minus (-) rating.

Studies were not excluded based on their risk of bias score. Methodological quality assessment was conducted by one reviewer and checked by a second, with any disagreements resolved by a third person.

### 4.7 Synthesis

The data is reported narratively as a series of thematic summaries (Thomas et al. 2017) and will be structured around the outcomes, target population characteristics, and type of intervention.

### 4.8 Assessment of body of evidence

The strength of the evidence (overall findings based on each outcome) was established and graded based on the quality, quantity and consistency of the available evidence (see Table 5), as well as the findings and the likely clinical impact (Academy of Nutrition and Dietetics 2022). Due to the heterogeneity between the included studies (i.e. each study evaluated a different intervention using different outcomes) this process was conducted at an individual study level. The Grade levels using this approach are as follows:

***Grade I: Good / strong***—The evidence consists of results from studies of strong design for answering the question addressed. The results are both clinically important and consistent with minor exceptions at most. The results are free of serious doubts about generalizability, bias, and flaws in research design. Studies with negative results have sufficiently large sample sizes to have adequate statistical power.

***Grade II: Fair***—The evidence consists of results from studies of strong design answering the question addressed, but there is uncertainty attached to the conclusion because of inconsistencies among the results from different studies or because of doubts about generalizability, bias, research design flaws, or adequacy of sample size. Alternatively, the evidence consists solely of results from weaker designs for the questions addressed, but the results have been confirmed in separate studies and are consistent with minor exceptions at most.

***Grade III: Limited / weak***—The evidence consists of results from a limited number of studies of weak design for answering the questions addressed. Evidence from studies of strong design is either unavailable because no studies of strong design have been done or because the studies that that have been done are inconclusive due to lack of generalizability, bias, design flaws, or inadequate sample sizes.

## 5. EVIDENCE

### 5.1 Search results and study selection

The study selection process is shown in Figure 5. The searches identified 3,451 records across the five databases that were searched. Deduplication removed 1,093 records and a further 637 were removed by text mining. The title and abstracts of the remaining 1,721 records were screened and 1,655 were excluded as irrelevant to the review. The full papers of 66 records were retrieved and assessed for inclusion and a further 51 records were excluded. The details of these records and reasons for exclusion are shown in Appendix 2. The remaining 15 records met the inclusion criteria of the review and reported on 14 studies. Citation searching identified two additional records that were both companion papers to studies identified in the database searches and included in the review. Therefore, a total of 17 records were included, reporting on 14 studies.

### 5.2 Study designs

Four of the included studies were classified as RCTs (Arean et al. 2008, Martin et al 2022, Mukadam et al. 2018, Sanchez et al. 2022). Using the decision algorithm developed by Leatherdale (2019) the remaining search designs were classified below and also presented across Figures 1- 4. *Cross-sectional post-test only quasi-experimental (four studies)* (Alvidrez et al. 2005, Evans et al. 2014; Gorman et al. 2021, Michelson and Day 2014). One study collected data during a control group (Evans et al. 2014) and three studies compared the post-intervention data to data from historical comparison groups (Alvidrez et al. 2005; Gorman et al 2021; Michelson and Day 2014).

**Figure 1:**
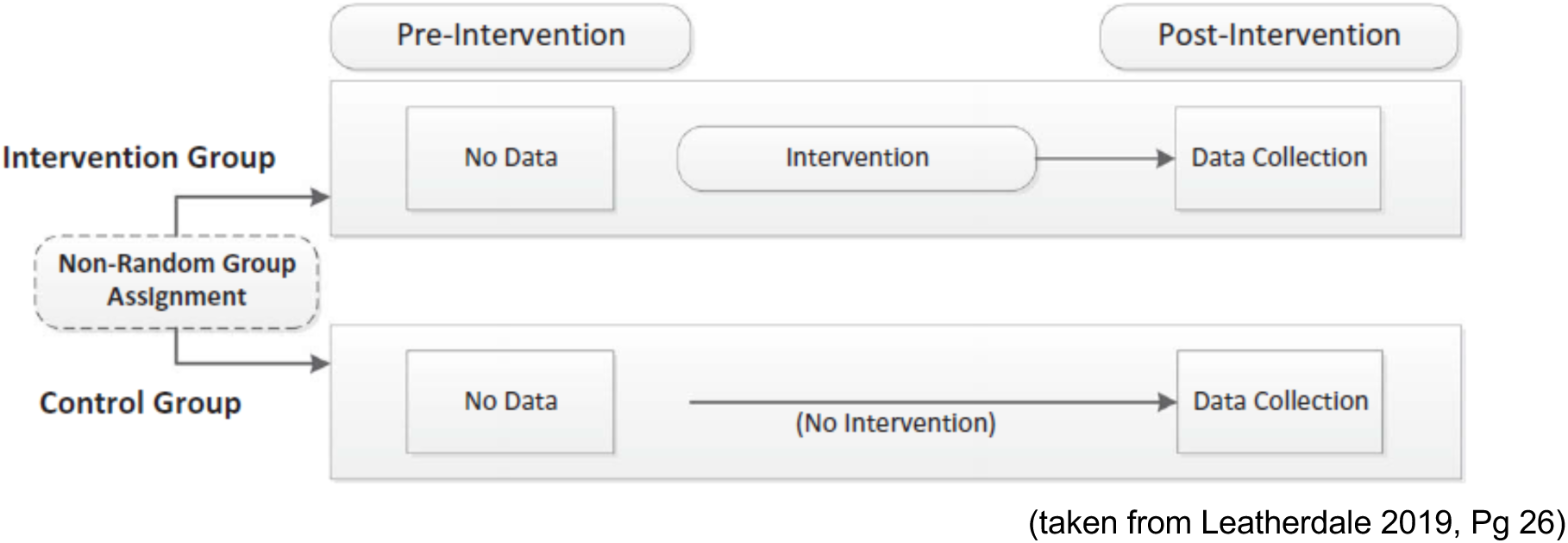
**Cross-sectional post-test only quasi-experimental**

*Repeat cross-sectional pre-post non-experimental studies (four studies)*.

(Hackett et al. 2009; Robidoux et al. 2023; Seabrooke and Milne 2009; Yeung et al. 2004).

**Figure 2:**
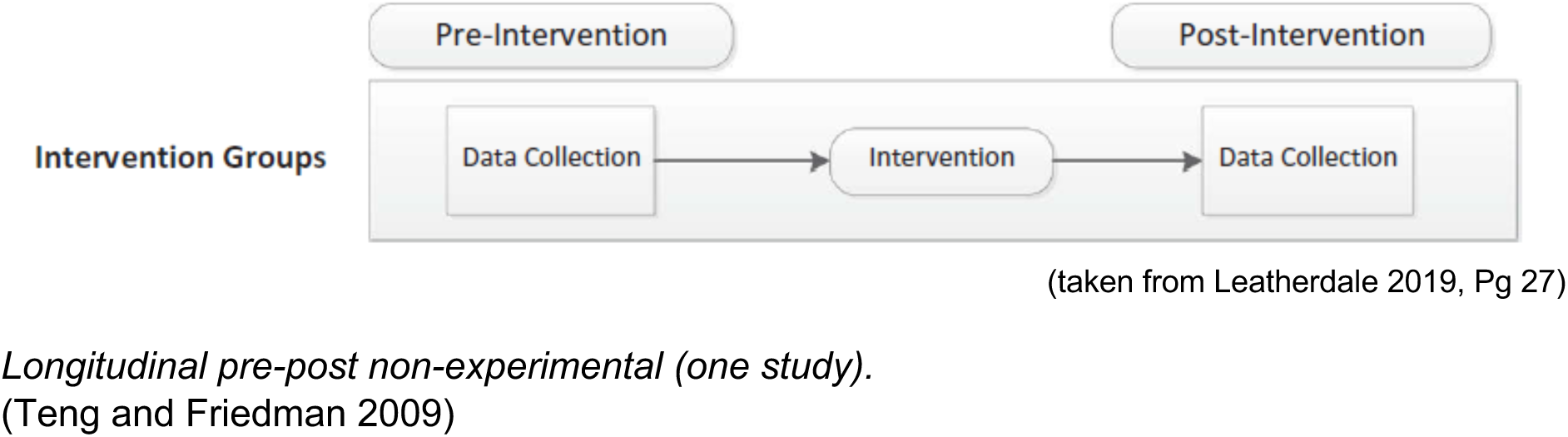
**Repeat cross-sectional pre-post non-experimental**

**Figure 3:**
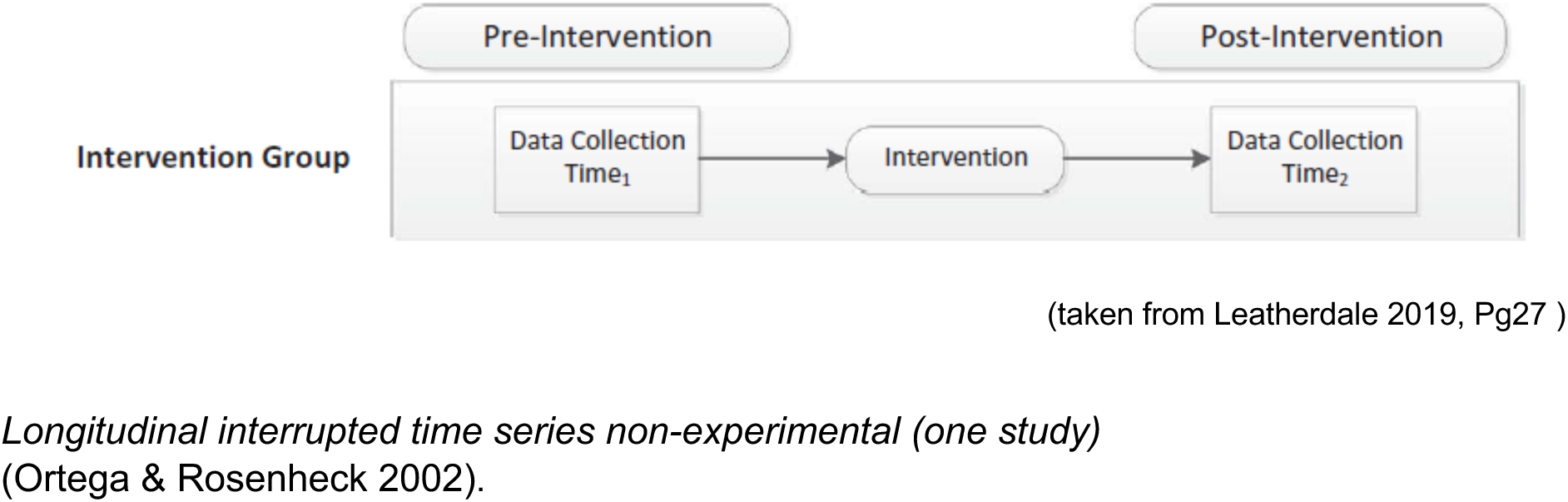
**Longitudinal pre-post non-experimental**

**Figure 4:**
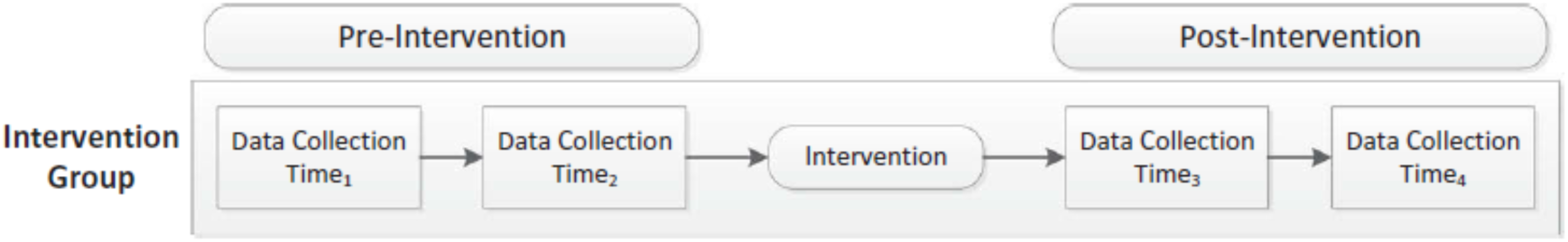
**Longitudinal interrupted time series non-experimental**

**Figure 5:**
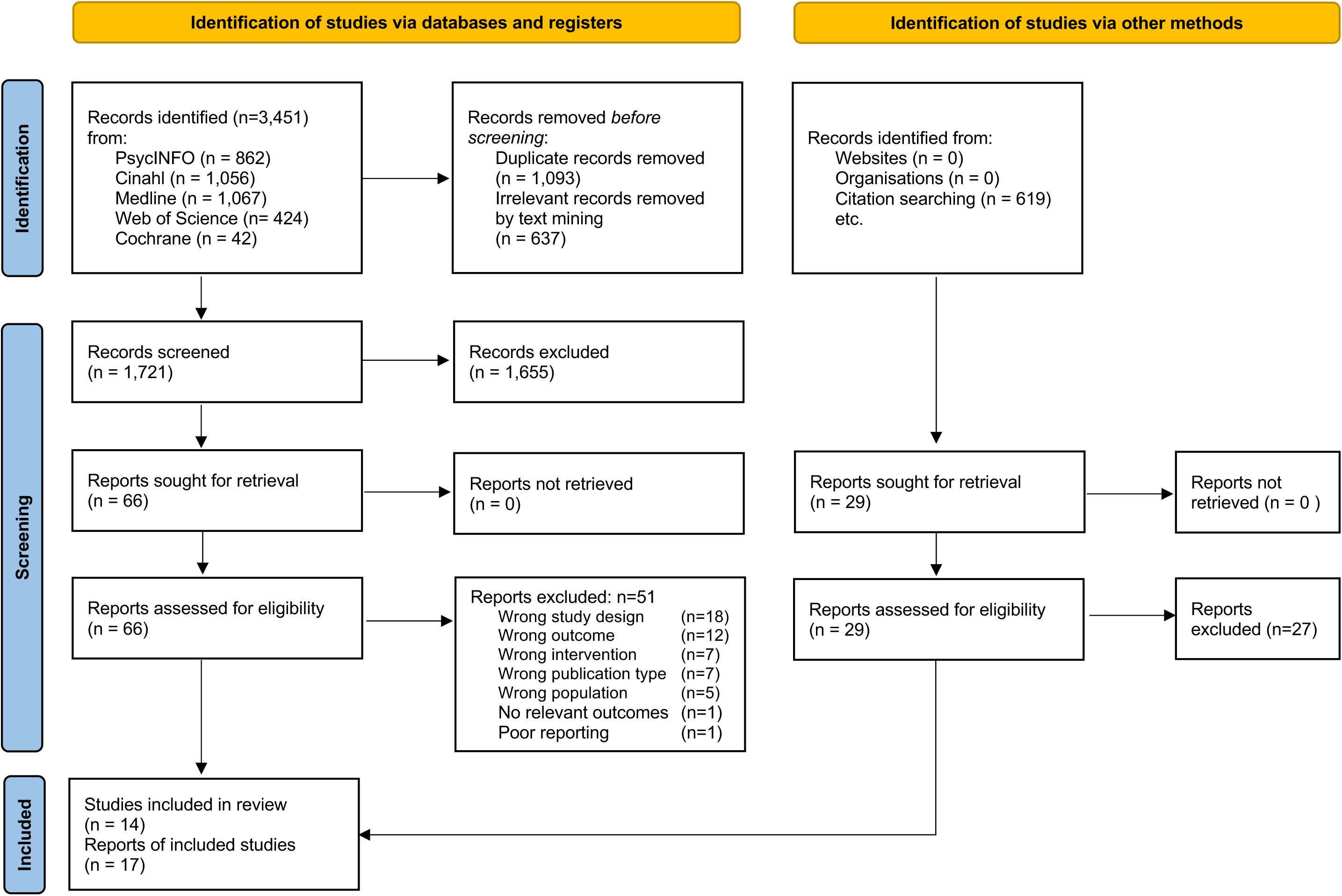
**PRISMA flow diagram of study identification**

### 5.3 Data extraction

Table 6 below presents the characteristics of the included studies. The data includes bibliographic information and the country in which the study was conducted, aim/s, study details, including details of the study design and intervention as well as outcome measures, information about the participants and setting, results of the critical appraisal, and key findings.

### 5.4 Quality appraisal

#### QCC Questions

1. Was the <u>research question</u> clearly stated?
2. Was the selection of study subjects/patients free from bias?
3. Were study groups comparable?
4. Was method of handling <u>withdrawals</u> described?
5. Was blinding used to prevent introduction of bias?
6. Were intervention/therapeutic regimens/exposure factor or procedure and any comparison(s) described in detail? Were intervening factors described?
7. Were outcomes clearly defined and the measurements valid and reliable?
8. Was the <u>statistical analysis</u> appropriate for the study design and type of outcome indicators?
9. Are <u>conclusions supported by results</u> with biases and limitations taken into consideration?
10. Is bias due to study’s <u>funding or sponsorship</u> unlikely?

#### QCC Overall designation criteria

- Low (-) If most (six or more) of the answers to the above validity questions are “No,” the report should be designated with low (-)
- Medium(Ø) If the answers to any of the first four validity questions (1-4) is “No”, but other criteria indicate strengths, the report should be designated medium (Ø
- High (+) If most of the answers to the above validity questions (six or more) are “Yes” (must include 1, 2, 3, and 4), the report should be designated high (+)
- When a validity criteria question is N/A
- `o` If any of the ten validity questions are N/A, the report requires a majority of “Yes” answers (including 1, 2, 3,4, as applicable) for a (+), or a majority or “No” answers for a (-) rating

## 6. ADDITIONAL INFORMATION

### 6.1 Protocol

The protocol for this rapid review is available on Open Science Framework https://osf.io/xpfje/

## Acknowledgements

The authors would like to thank Beverly Morgan, Kim Swain, Hannah Bayfield, Thomas Hoare, Holly Howe-Davies, Olivia Gallen and Mel (Melanie) McAulay for their contributions during stakeholder meetings in guiding the focus of the review, interpretation of findings and commenting on draft versions of the report.

## Research Implications and Evidence Gaps

Future research should prioritise rigorous study designs, including randomised controlled trials and longitudinal studies. More research is required to compare outcomes between ethnic minority participants and White participants.

## Economic considerations

Ethnic minority individuals are disproportionately affected by economic determinants of poor mental health including increased likelihood of low income. Future research should investigate the economic benefit from an NHS and societal perspective of improving access to mental health services for ethnic minority individuals.

## Abbreviations

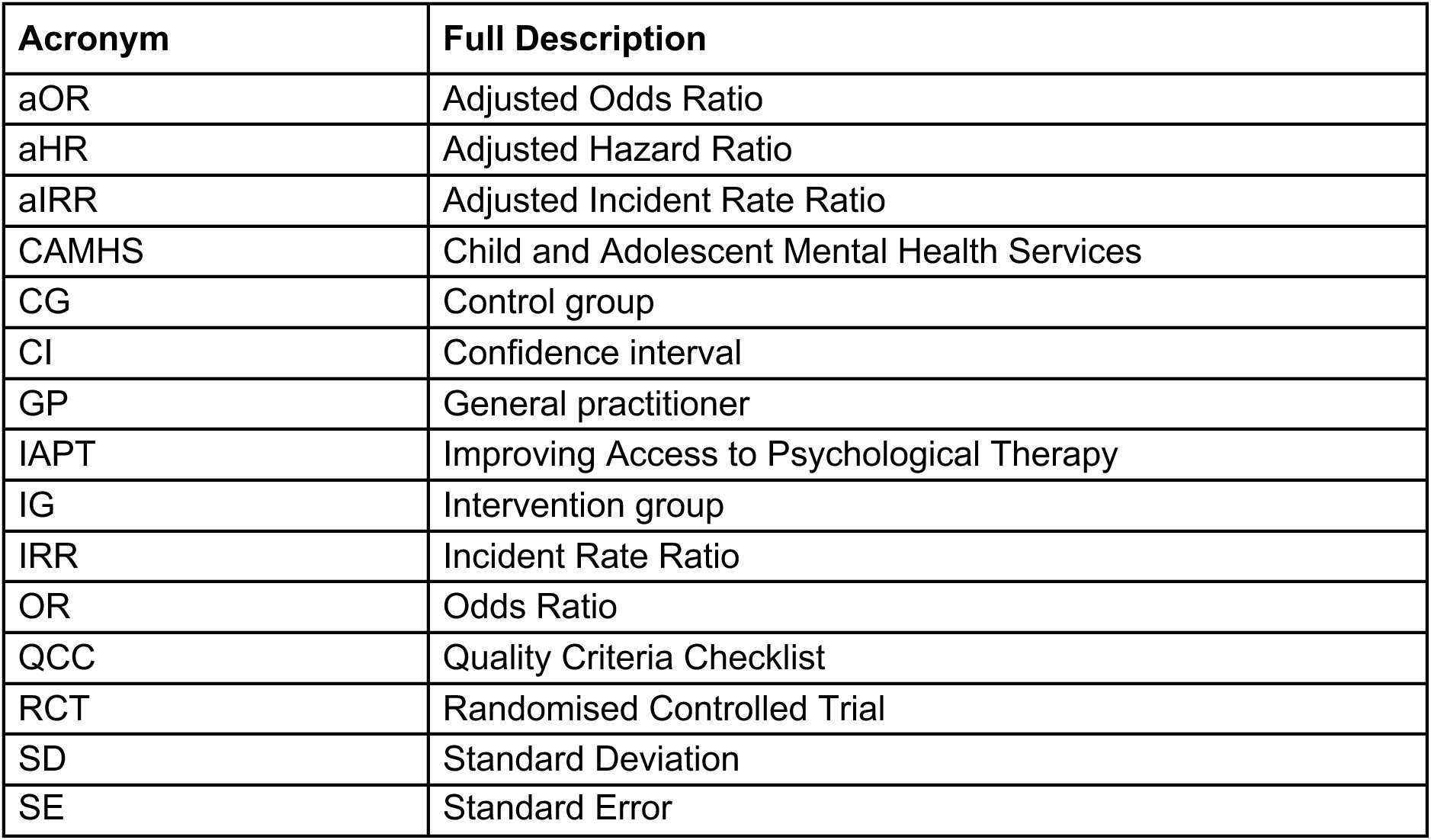

| Acronym | Full Description |
| --- | --- |
| aOR | Adjusted Odds Ratio |
| aHR | Adjusted Hazard Ratio |
| aIRR | Adjusted Incident Rate Ratio |
| CAMHS | Child and Adolescent Mental Health Services |
| CG | Control group |
| CI | Confidence interval |
| GP | General practitioner |
| IAPT | Improving Access to Psychological Therapy |
| IG | Intervention group |
| IRR | Incident Rate Ratio |
| OR | Odds Ratio |
| QCC | Quality Criteria Checklist |
| RCT | Randomised Controlled Trial |
| SD | Standard Deviation |
| SE | Standard Error |

## Glossary

### Disparity

“Health disparity and health inequality are broad terms that include health inequity and signify more than just difference or variation: they signify a health difference that raises moral or ethical concerns.” (Braveman et al. 2018, p.11)

### Equity

Health equity means that “everyone has a fair and just opportunity to be as healthy as possible. Achieving health equity requires removing obstacles to health such as poverty, discrimination, and their consequences, which include powerlessness and lack of access to good jobs with fair pay; quality education, housing, and health care; and safe environments. For the purposes of measurement, health equity means reducing and ultimately eliminating disparities in health and health determinants that adversely affect excluded or marginalized groups. Health equity is the ethical and human rights principle motivating efforts to eliminate health disparities; health disparities are the metric for assessing progress toward health equity.” (Braveman et al. 2018, p. 11)

### Ethnic minorities

refers to all ethnic groups except the White British group. Ethnic minorities include white minorities, such as Gypsy, Roma and Irish Traveller groups. (UK Government 2023)

### Mental health

Welsh Government defines mental health (in the draft Mental Health and Wellbeing Strategy – out to consultation February to June 2024) as a state of mental wellbeing that enables people to cope with the stresses of life, realise their abilities, learn well and work well, and contribute to their community. It is an integral component of health and wellbeing that underpins our individual and collective abilities to make decisions, build relationships and shape the world we live in. Mental health is a basic human right. And it is crucial to personal, community and socio-economic development. People with poor mental health can have a mental health condition but this is not always or necessarily the case.

### Mental health condition

Welsh Government defines mental health condition (in the draft Mental Health and Wellbeing Strategy – out to consultation February to June 2024) as a broad term covering conditions that affect emotions, thinking and behaviour, and which substantially interfere with our life. Mental health conditions can significantly impact daily living, including our ability to work, care for ourselves and our family, and our ability to relate and interact with others. This is a term used to cover several conditions (e.g. depression, post-traumatic stress disorder, schizophrenia) with different symptoms and impacts for varying lengths of time, for each person. Mental health conditions can range from mild through to severe and enduring illness. People with mental health conditions are more likely to experience lower levels of physical and mental wellbeing, but this is not always or necessarily the case. Some mental health conditions like eating disorders and schizophrenia are associated with a higher risk of mortality.

## APPENDIX

### Appendix 1: Search strategies

Database: APA PsycINFO

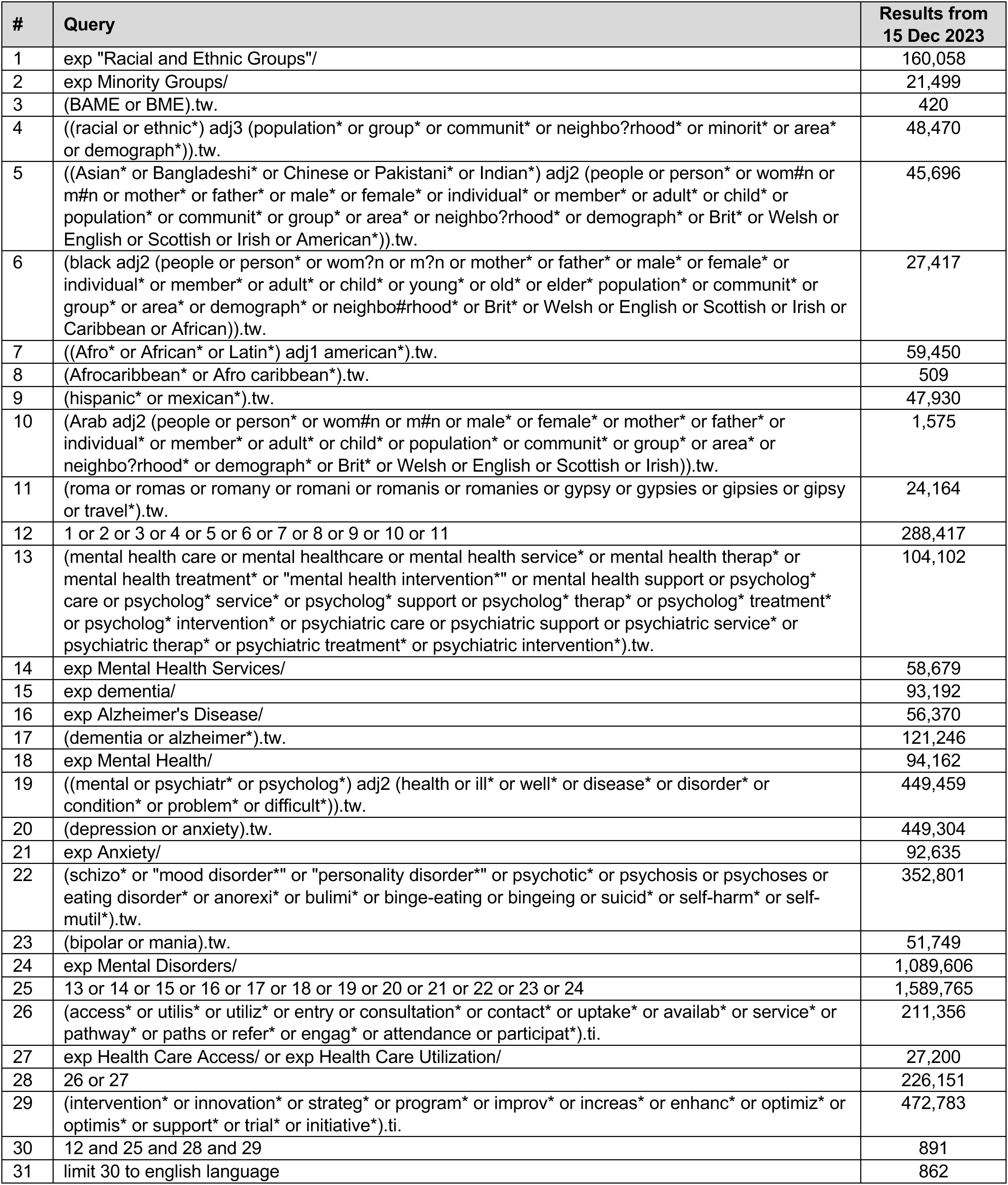
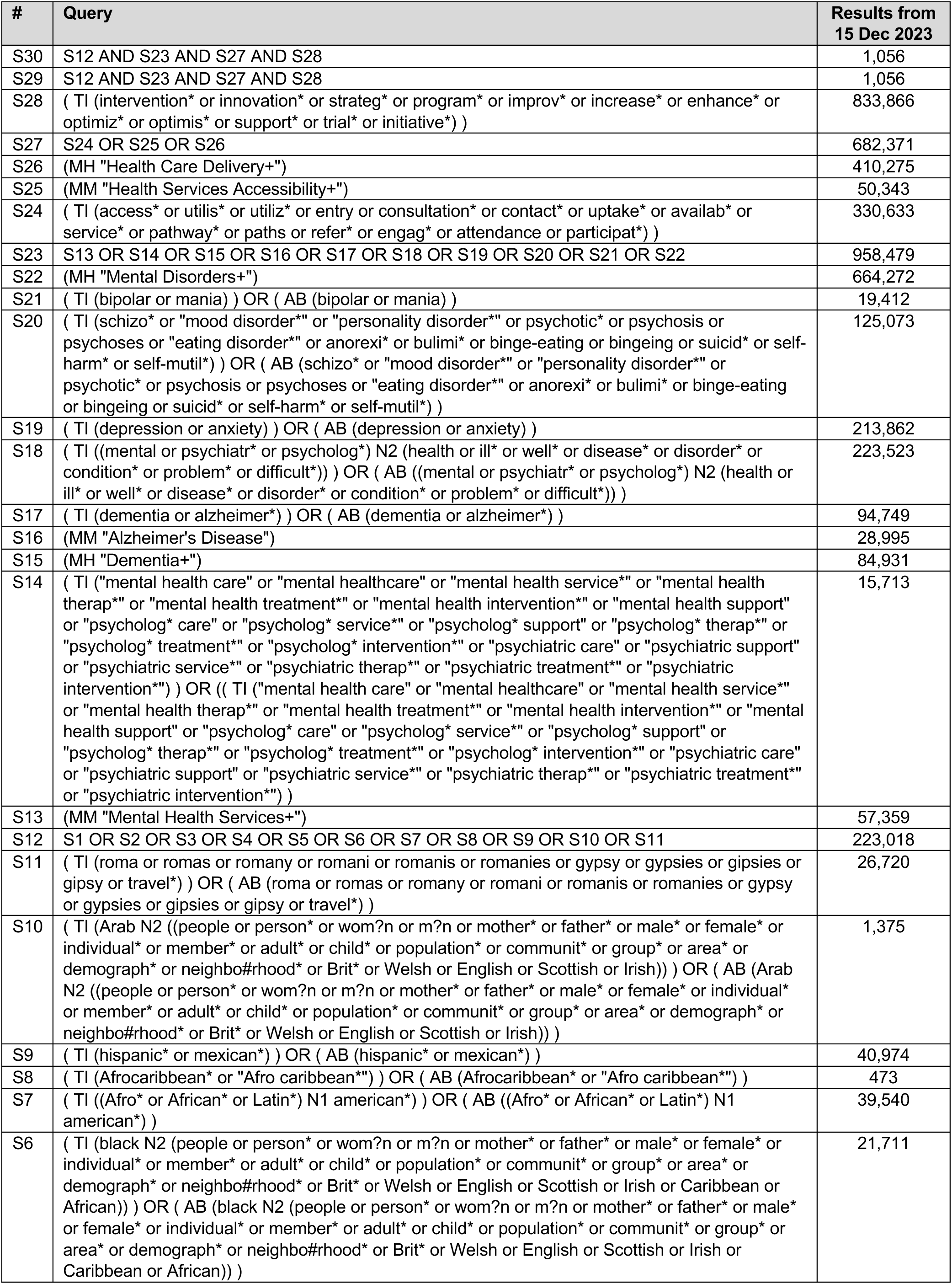
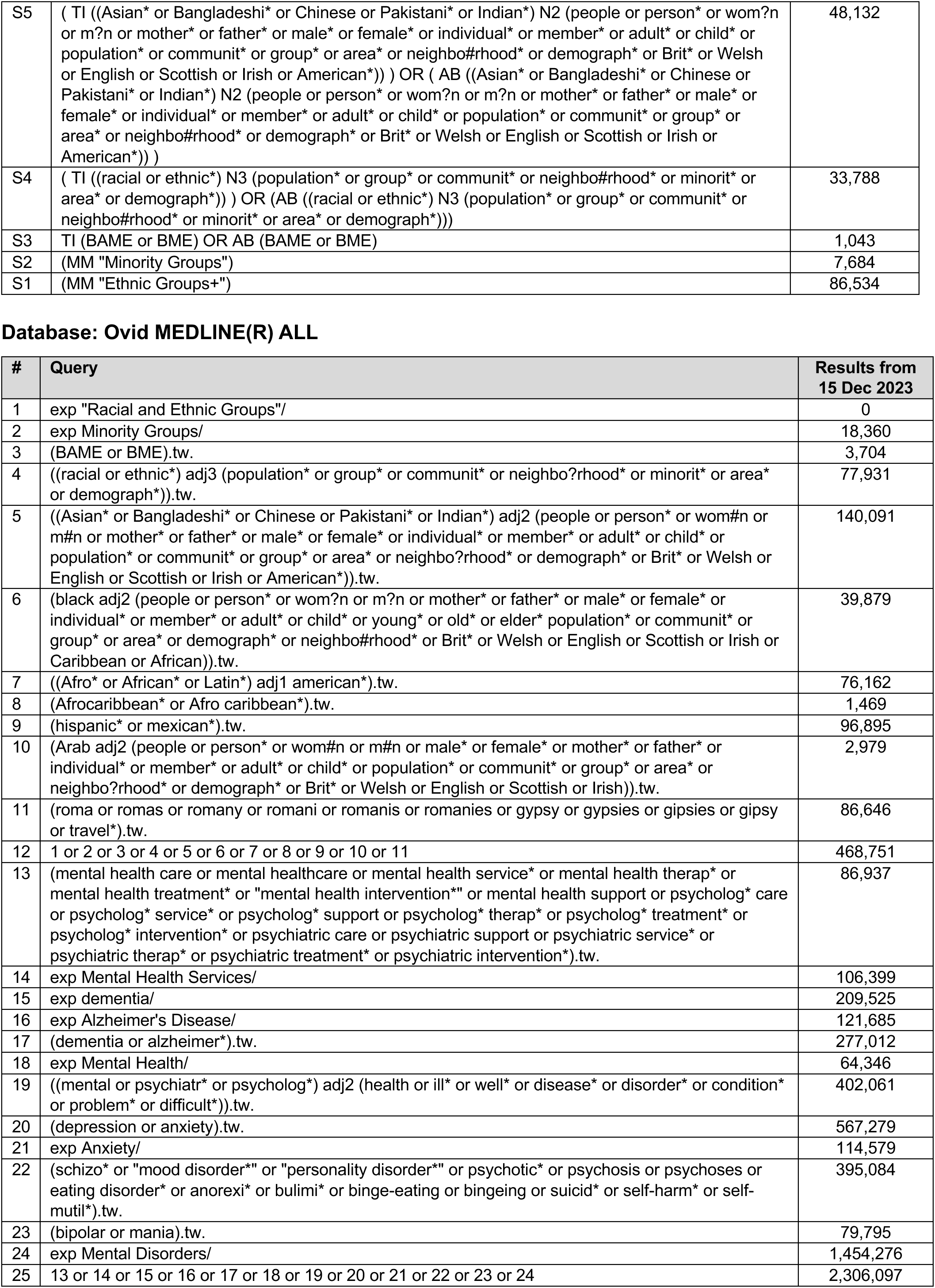
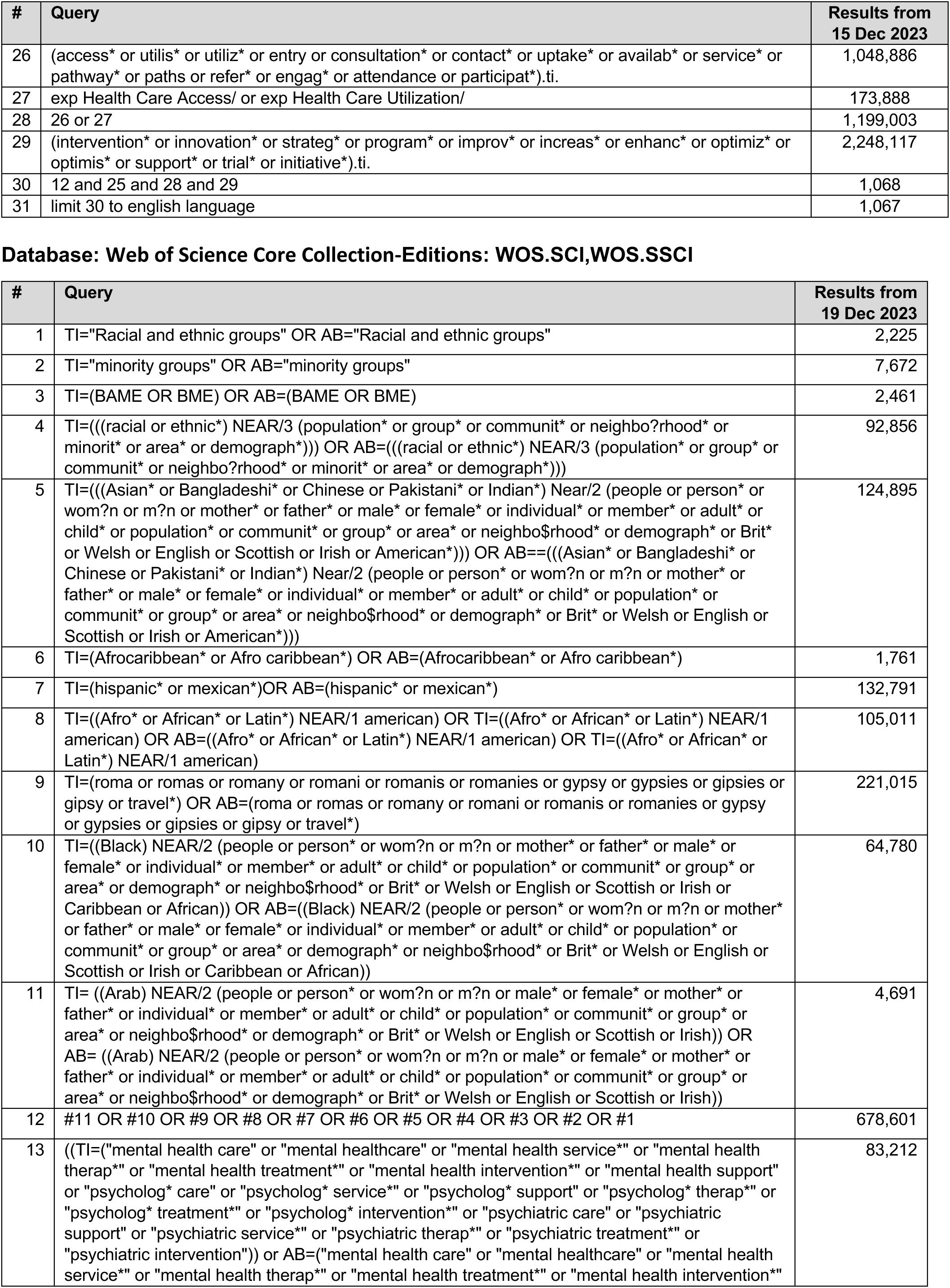
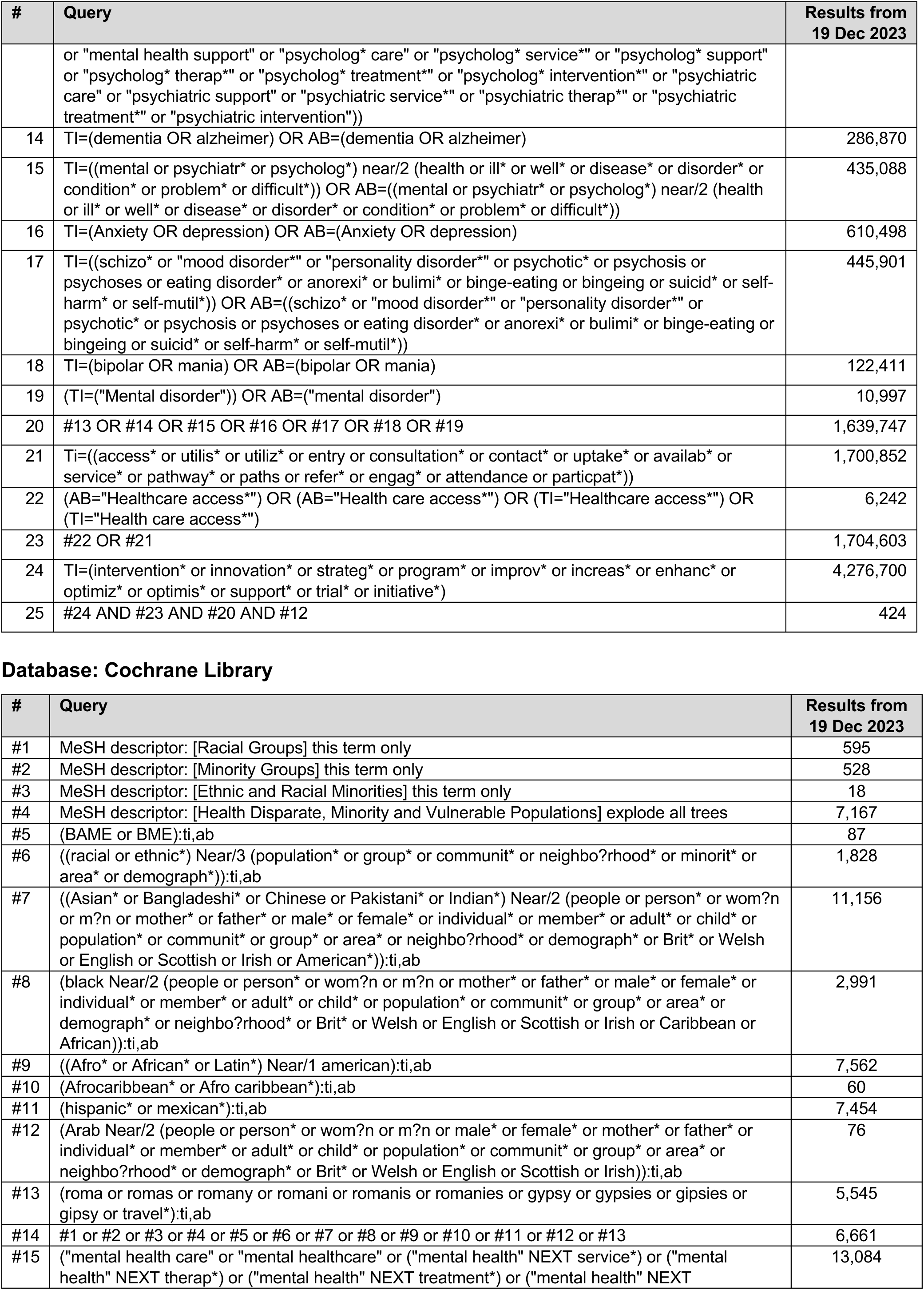
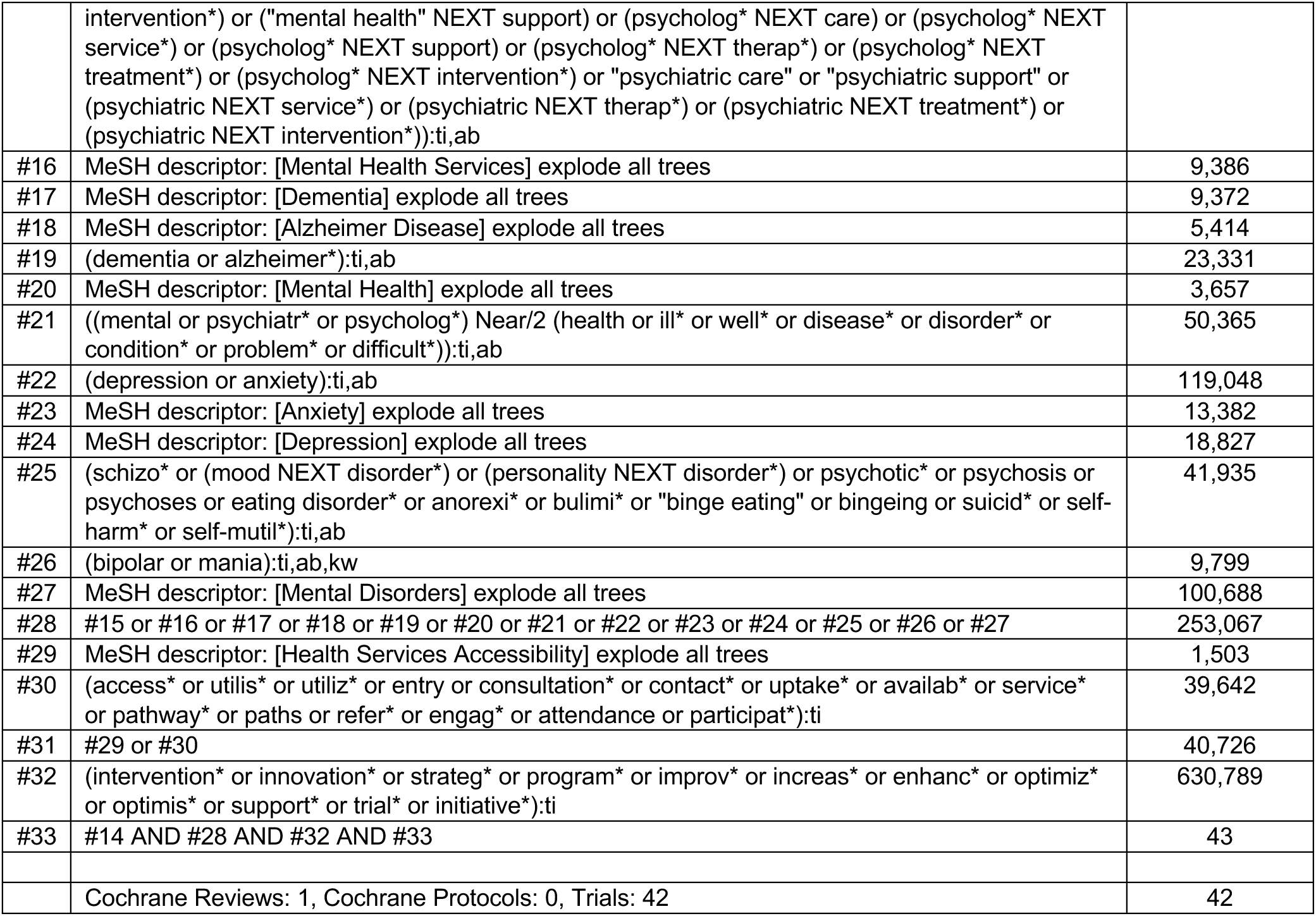

### APPENDIX 2: Excluded studies

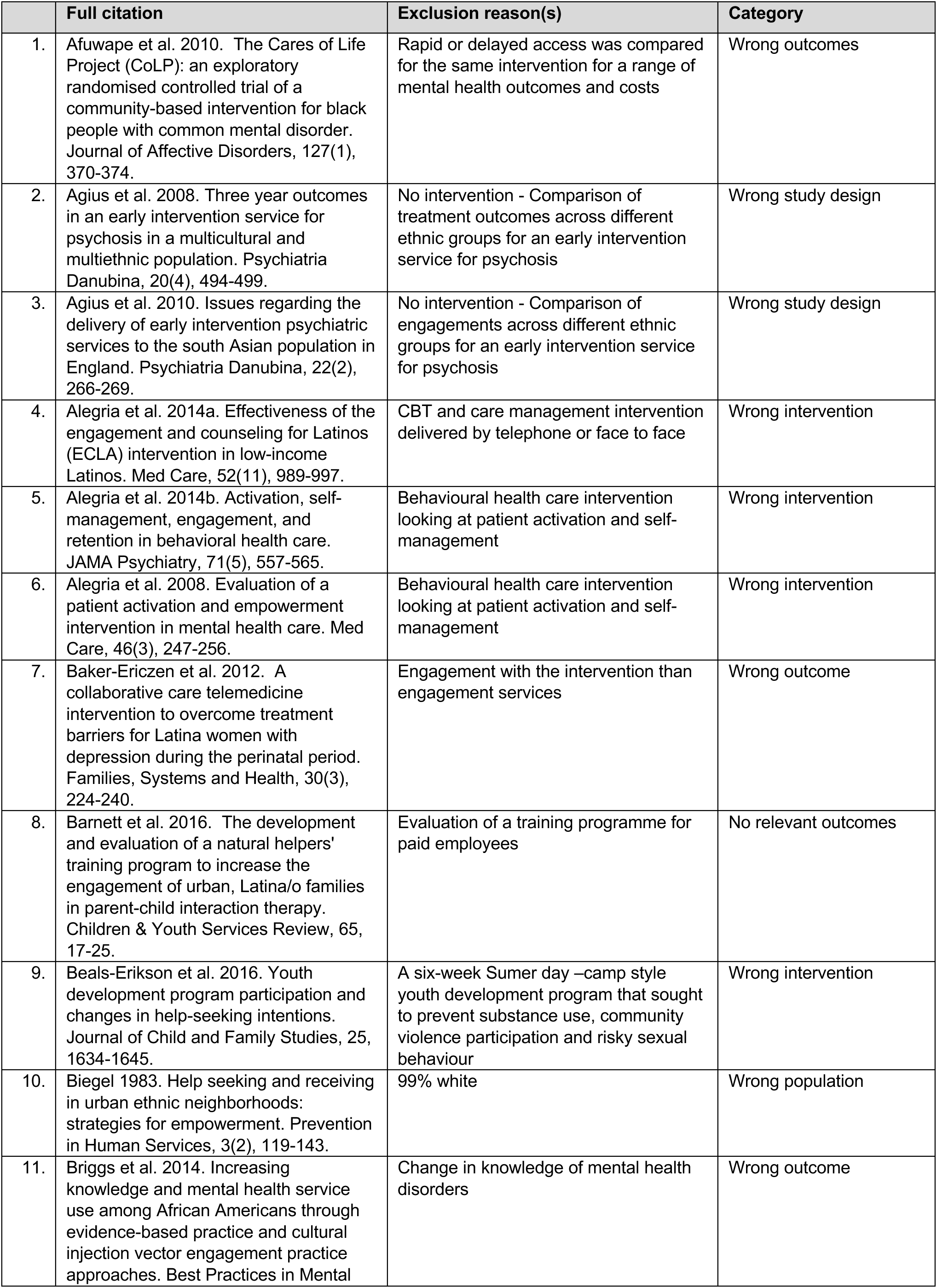
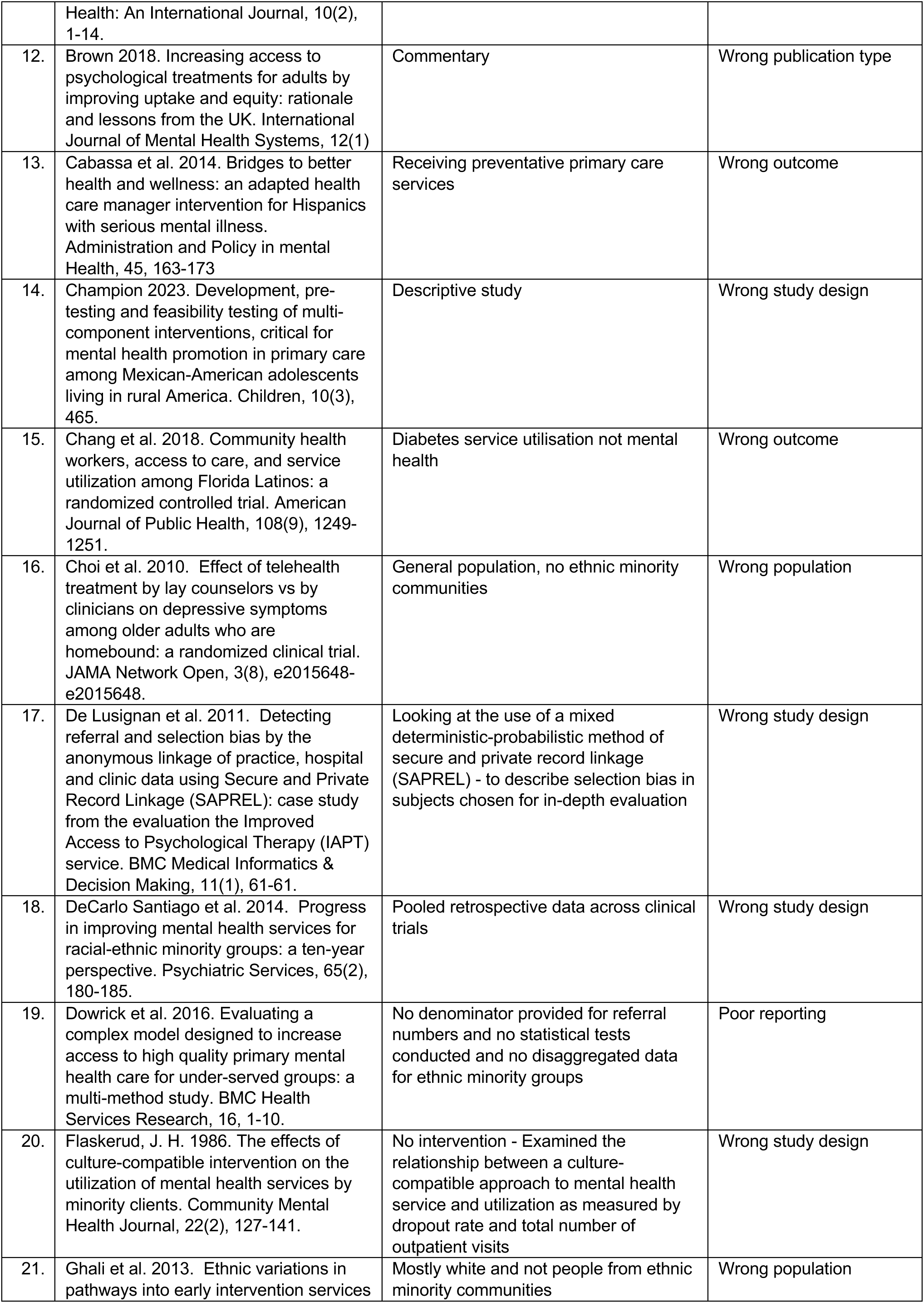
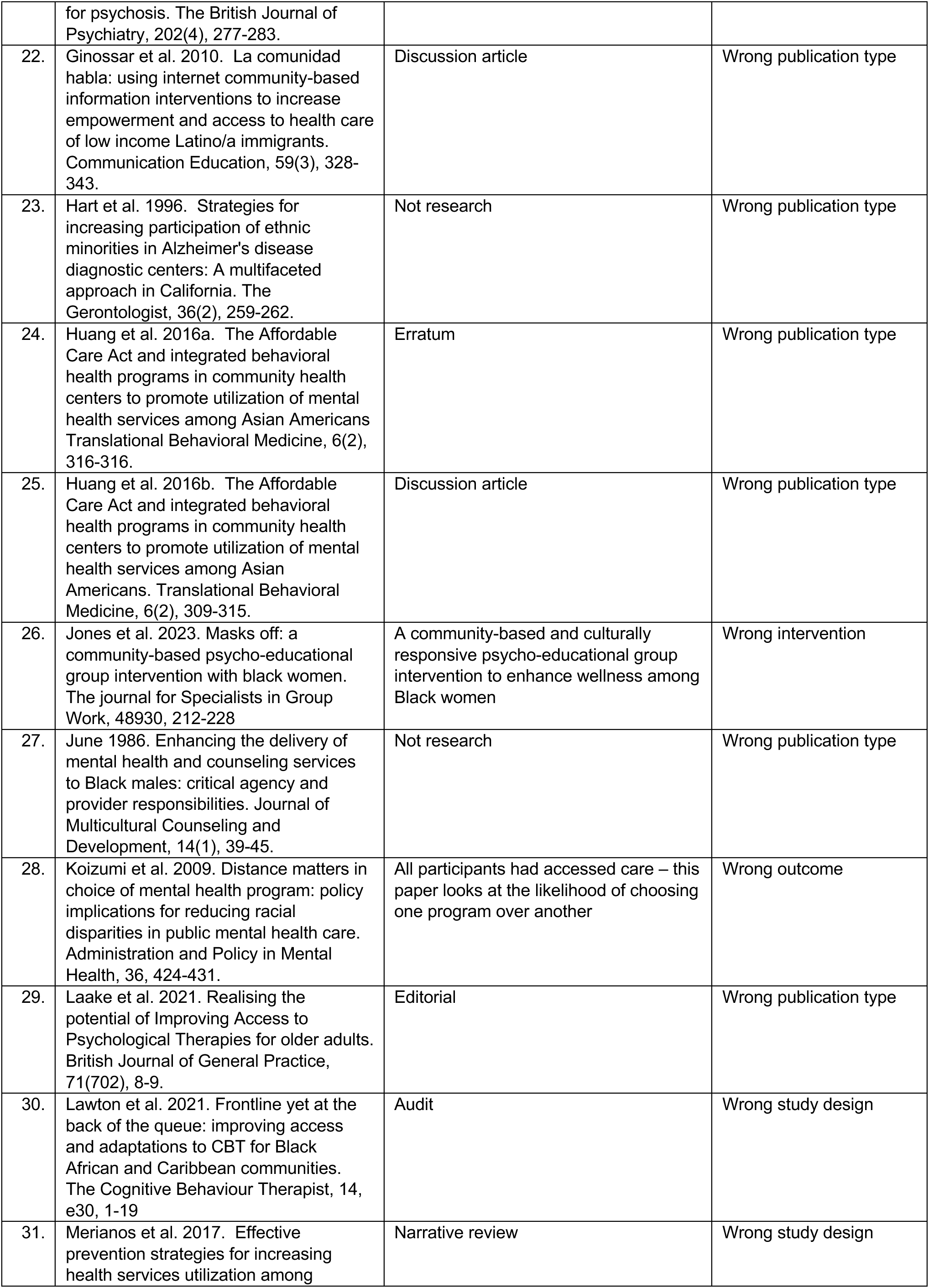
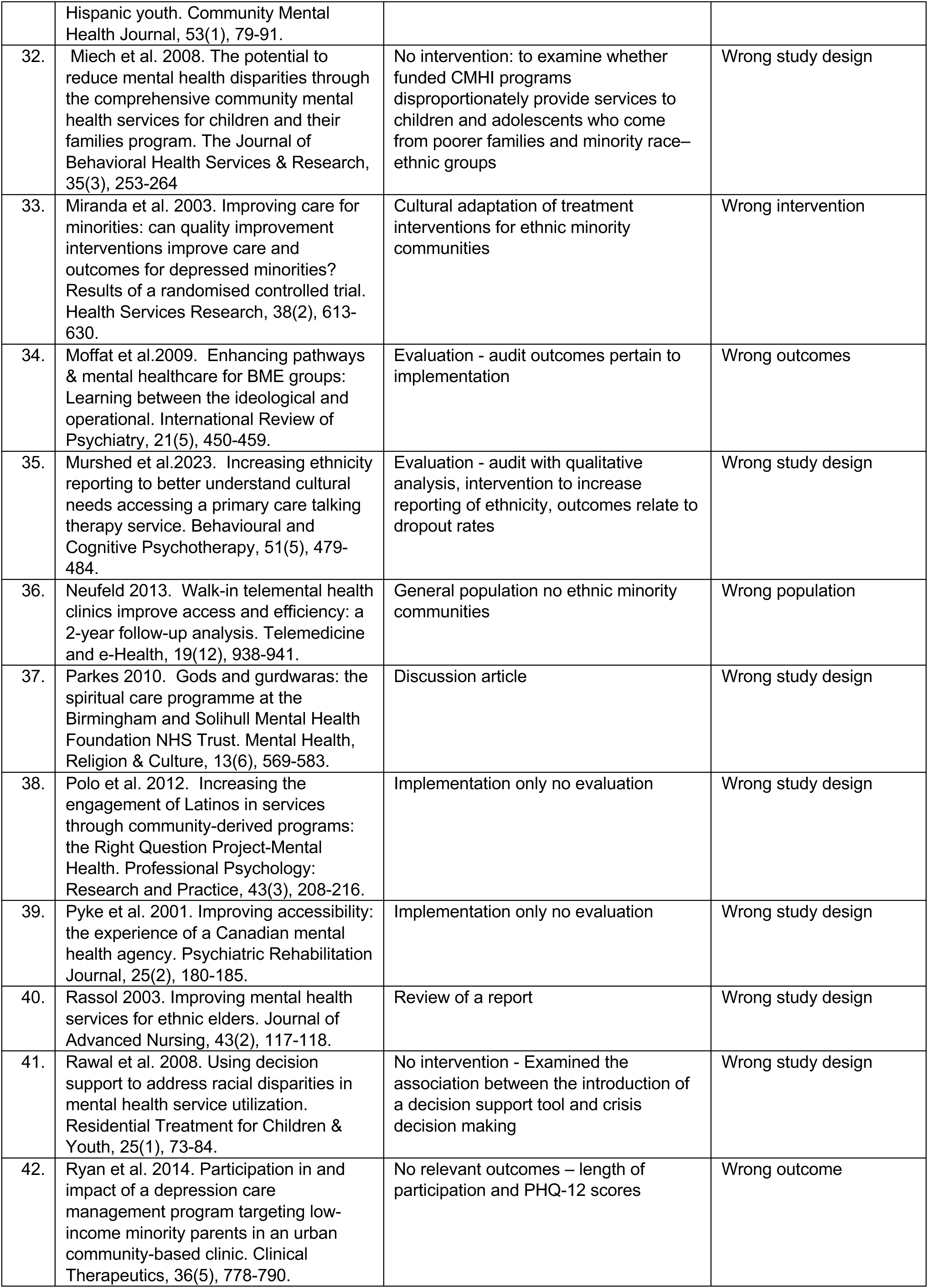
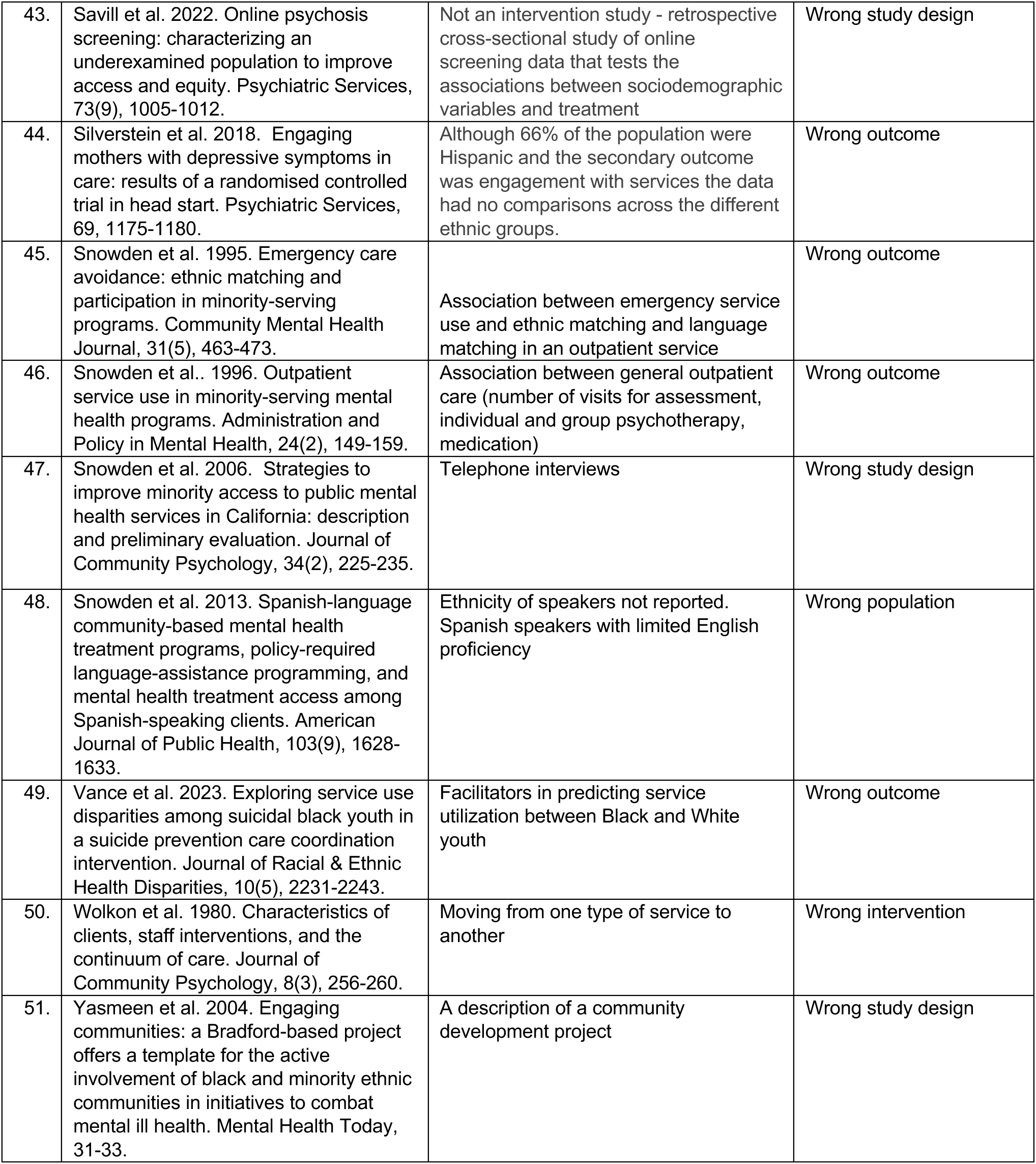

## Footnotes

1 Defined in the Glossary

2 Defined in the Glossary

3 Integrating mental health and substance abuse services in primary care reported across three studies – Arean et al. 2008, Ayalon et al. 2007 and Levkoff et al. 2004 and referred to as Arean et al. 2008 through this report.

4 Educational leaflet and DVD about dementia and getting help for memory symptoms reported across Mukadam et al. 2018 and Mukadam 2017 and referred to as Mukadam 2017.

5 Surrogate outcome for help seeking.

6 Significance level set as p<0.001 by the study authors

7 Significance level set as p<0.001 by the study authors.

8 17 out of 41 people assessed the intervention and of these 10 looked only at the leaflet, three only looked at the DVD and four people looked at both leaflet and DVD.

9 The study authors commented that there may have been multiple barriers such as stigma that preventing this population from consulting mental health experts.

10 Adjusted for baseline levels of distress as measured by the General Health Questionnaire

11 The authors do not discuss why access was decreased for Asian participants.

12 Adjusted for baseline levels of distress as measured by the General Health Questionnaire

13 The authors do not discuss why Asian participants had a significantly greater number of visits in specialty mental health services compared to integrated services into primary care.

14 No statistical analyses reported

15 The authors do not discuss routine depression screening was more likely for Asian participants.

16 This is a positive outcome, indicating success in meeting the needs of Hispanic clients in the community.

17 The authors suggest possible reasons for this: i) culturally relevant care may be more important for Spanish- speaking clients. ii) Hispanics are underrepresented in homeless populations due to strong family networks; those who become homeless may have more severe psychiatric illnesses. Iii) Ethnic matching may offer limited benefits for homeless Hispanics in the program, as their primary needs are met regardless of ethnicity.

18 No statistical analyses reported.

19 The authors identified multiple barriers to referral during implementation, reflecting concerns documented in existing literature which included time constraints, lack of a referral process, difficulty in finding mental health resources, uninsured patients, long wait times at free clinics, and selective referral based on screening scores.

20 No statistical analyses reported.

21 Ethnicity: Hispanic/Latinx (53.8%), Haitian (2.6%), not Hispanic or Haitian (43.6%) Race: American Indian (2.6%), Asian (2.6%), Black or African American (28.2%), White (56.4%), Bi/Multiracial (10.3%)

22 Ethnicity: Hispanic/Latinx (53.8%), Haitian (2.6%), not Hispanic or Haitian (43.6%) Race: American Indian (2.6%), Asian (2.6%), Black or African American (28.2%), White (56.4%), Bi/Multiracial (10.3%),

